# Genomic landscape of *TP53*-mutated myeloid malignancies

**DOI:** 10.1101/2023.01.10.23284322

**Authors:** Haley J. Abel, Karolyn A. Oetjen, Christopher A. Miller, Sai M. Ramakrishnan, Ryan B. Day, Nichole M. Helton, Catrina C. Fronick, Robert S. Fulton, Sharon E. Heath, Stefan P. Tarnawsky, Sridhar Nonavinkere Srivatsan, Eric J. Duncavage, Molly C. Schroeder, Jacqueline E. Payton, David H. Spencer, Matthew J. Walter, Peter Westervelt, John F. DiPersio, Timothy J. Ley, Daniel C. Link

## Abstract

*TP53*-mutated myeloid malignancies are most frequently associated with complex cytogenetics. The presence of complex and extensive structural variants complicates detailed genomic analysis by conventional clinical techniques. We performed whole genome sequencing of 42 AML/MDS cases with paired normal tissue to characterize the genomic landscape of *TP53*-mutated myeloid malignancies. The vast majority of cases had multi-hit involvement at the *TP53* genetic locus (94%), as well as aneuploidy and chromothripsis. Chromosomal patterns of aneuploidy differed significantly from *TP53*-mutated cancers arising in other tissues. Recurrent structural variants affected regions that include *ETV6* on chr12p, *RUNX1* on chr21, and *NF1* on chr17q. Most notably for *ETV6*, transcript expression was low in cases of *TP53*-mutated myeloid malignancies both with and without structural rearrangements involving chromosome 12p. Telomeric content is increased in *TP53*-mutated AML/MDS compared other AML subtypes, and telomeric content was detected adjacent to interstitial regions of chromosomes. The genomic landscape of *TP53*-mutated myeloid malignancies reveals recurrent structural variants affecting key hematopoietic transcription factors and telomeric repeats that are generally not detected by panel sequencing or conventional cytogenetic analyses.

**Key Points:**

- WGS comprehensively determines *TP53* mutation status, resulting in the reclassification of 12% of cases from mono-allelic to multi-hit
- Chromothripsis is more frequent than previously appreciated, with a preference for specific chromosomes
- *ETV6* is deleted in 45% of cases, with evidence for epigenetic suppression in non-deleted cases
- *NF1* is mutated in 48% of cases, with multi-hit mutations in 17% of these cases
- *TP53*-mutated AML/MDS is associated with altered telomere content compared with other AMLs

## Introduction

Myeloid malignancies with complex cytogenetics have adverse prognostic risk[1], and *TP53* mutations are present in 70% of these cases[2]. Extensive structural variation makes it difficult to completely characterize the genomic profile using standard clinical techniques for diagnosis and risk stratification[1,3], such as karyotyping, fluorescence in situ hybridization (FISH), targeted sequencing panels, and immunohistochemistry. The presence of multiple genomic alterations of the *TP53* locus has become a distinguishing characteristic of extremely poor prognosis in myelodysplastic syndromes (MDS)[4] and acute myeloid leukemia (AML)[5]. While multi-hit *TP53* events may be detected by sequencing panels and cytogenetics, they are limited in resolution and generally rely on indirect evidence, such as high variant allele frequency (VAF) in sequencing panels exceeding 50%[1] or 55%[5], and cannot deconvolute the effects of purity, ploidy, and/or clonality. Alternate approaches include DNA microarray, but small copy number changes or complex structural events remain undetected with this method.

Whole genome sequencing (WGS) with adequate genome coverage can detect nearly all copy number changes, structural variants (SV), single nucleotide variants (SNV), and small insertion/deletions (indels) in a tumor. It can supplant clinical characterization by cytogenetics and targeted sequencing[6] in an unbiased fashion (i.e. not limited to specific probes on FISH or sequencing panels). In this study, we performed WGS on 42 samples from patients with *TP53*-mutated AML or MDS. Paired normal tissue from each patient was included for analysis to ensure accurate distinction of somatic and germline variants for SNVs, indels and structural variants. Multiple hits at the *TP53* locus occurred in 41 of the 42 total cases (94%) through a combination of SNV/indels, copy number loss, and/or copy-number neutral loss of heterozygosity (CN-LOH). Furthermore, five of these cases (12%) could be accurately classified only with WGS analysis, rather than conventional clinical testing. Nearly all “multi-hit” events (92%) resulted in biallelic inactivation of *TP53* through SNVs, indels, SVs or CN-LOH, though a small percentage could not be confidently resolved as biallelic due to the limited resolution of short-read sequencing. Although aneuploidy is a well-established feature of *TP53*-mutated malignancies, patterns of recurrent copy number changes, structural variants, chromothripsis, and telomeric repeats differ remarkably between *TP53*-mutated myeloid malignancies and other hematopoietic and solid tumors reported in the PCAWG consortium[7–11]. In this study, we demonstrate that recurrent copy number and structural variants in *TP53*-mutated myeloid malignancies affect key hematopoietic transcription factors and signaling pathways.

## Materials/Subjects and Methods

### Patient selection

Cases were selected from patients with pathogenic mutations or structural rearrangements in *TP53* identified through panel sequencing, whole genome sequencing, or cytogenetics performed for research or clinical purposes at Washington University in St. Louis, including published[6,12,13] and newly identified cases. All patients provided written informed consent for tissue repository and genomic sequencing in accordance with protocol #201011766 approved by the Washington University in St Louis Institutional Review Board. Bone marrow aspirates were used for tumor analysis whenever available, otherwise peripheral blood was used.

### Whole genome sequencing

Whole genome sequencing of tumor samples was performed as previously described[6]. Additional WGS of cryopreserved tumor and skin/buccal extracted DNA was performed using Kapa PCR-free library preparation (Roche), including for previously published samples[12,13]. Sequencing was performed on an Illumina NovaSeq S4 with 150 bp paired end reads, targeting 60x coverage for tumors and 30x for matched normal samples. Sequencing depth and sample purity metrics for each case are summarized in Supplementary Table 1. For inclusion in this study, samples were required to have an estimated tumor purity of >=20% based on somatic variant allele frequencies.

**Table 1.** Clinical and molecular summary of myeloid malignancy cases.

| Cohort |  | TP53 | TP53 | CBF |
| --- | --- | --- | --- | --- |
| Diagnosis |  | AML, n=29 | MDS, n=13 | AML, n=18 |
| Sex | Male | 66% (19) | 31% (4) | 56% (10) |
|  | Female | 34% (10) | 69% (9) | 44% (8) |
| Age, mean (range) |  | 65.7 (26-81) | 61.3 (41-85) | 45.5 (20-76) |
| Ethnicity | Caucasian | 97% (28) | 100% (13) | 100% (18) |
|  | African American | 3% (1) |  |  |
| WBC, mean (10<sup>3</sup> cells/uL) |  | 9.3 | 5.0 | 34.0 |
| BM blast %, mean |  | 54% | 7% | 56% |
| PB blast %, mean |  | 14% | 2% | 39% |
| Subtype | M0 | 14% (4) |  |  |
|  | M1 | 7% (2) |  | 11% (2) |
|  | M2 | 17% (5) |  | 33% (6) |
|  | M4 | 10% (3) |  | 50% (9) |
|  | M5 |  |  | 6% (1) |
|  | M6 | 10% (3) |  |  |
|  | M7; possible M7 | 14% (4) |  |  |
|  | MRC | 10% (3) |  |  |
|  | Secondary; therapy-related | 7% (2) | 38% (5) |  |
|  | Excess blasts |  | 54% (7) |  |
|  | Refractory anemia |  | 3% (1) |  |
|  | Not categorized | 10% (3) |  |  |
| Cytogenetics | complex | 90% (26) | 100% (13) |  |
|  | inv(3) | 3% (1) |  |  |
|  | del(Y) | 3% (1) |  |  |
|  | inv(16) |  |  | 67% (12) |
|  | t(8;21) |  |  | 28% (5) |
|  | incomplete | 3% (1) |  | 6% (1) |
| Prior treatment | None | 100% (29) | 77% (10) | 100% (18) |
|  | Hypomethylating agent |  | 15% (2) |  |
|  | Lenolidomide |  | 8% (1) |  |
| TP53 status | Concordant, multi-hit | 93% (27) | 69% (9) |  |
|  | Discordant,<br>monoallelic->multi-hit | 7% (2) | 23% (3) |  |
|  | ambiguous->monoallelic |  | 8% (1) |  |

WBC: White blood cell count, BM: Bone marrow, PB: Peripheral blood,
MRC: Myelodysplastic-related changes

Sequence data were aligned against reference sequence GRCh38 using BWA-MEM[14]. Aligned reads were sorted, deduplicated, and recalibrated by base quality score. SNVs and small indels were detected using MuTect2[15], VarScan2[16], Strelka2[17], and GATK[18]. Structural variants were called using GRIDSS2[19]. PURPLE was used for copy number calling and detection of CN-LOH[20]. LINX was used for clustering of SV into complex variants[21]. Chromothripsis was assessed with ShatterSeek[7]. Telomeric content was assessed with TelomereHunter[22] and TelSeq[23]. Detection of telomeric insertions into intrachromosomal sequence was performed using an approach similar to previously described[8]. Refer to Supplementary Methods for full details of variant calling and telomere analysis.

### RNA sequencing

Cryopreserved cells from multi-hit *TP53*-mutated MDS/AML cases in this cohort (N=10), *TP53*-mutated MDS/AML with tumor-only WGS (N=4), CBF AML (N=11), and normal karyotype AML (M=52) were used for bulk RNA sequencing. RNA was extracted (Quick-RNA kit, Zymo Research) for preparation of total RNA sequencing libraries (TruSeq Stranded Total RNA kit with Unique Dual Indices, Illumina). Paired-end sequencing was performed at McDonnell Genome Institute on NovaSeq S4 flow cells (Illumina) using 2 × 150 bp read lengths to achieve 15Gb of coverage per sample. Transcript abundance was quantified with kallisto[24] using Ensembl[25] version 95 and scaled to library size and average transcript length. Normalized read counts were used for differential gene expression by edgeR[26].

## Results

### Clinical characteristics of patient cohorts

We performed WGS on paired tumor and normal tissue from 42 patients with *TP53*-mutated myeloid malignancies. The demographics of the cohort, including age, sex, ethnicity, blood counts and blast percentages at clinical presentation are summarized in Table 1. Detailed characteristics are provided in Supplementary Table 2. As a comparator, we analyzed 18 cases of AML with core-binding factor (CBF) translocations, which have defined structural alterations – inv(16) with *CBFB*::*MYH11* or t(8;21) with *RUNX1*::*RUNX1T1* fusions – and lack *TP53* mutations or complex cytogenetics (Table 1). The clinical characteristics of these groups were generally well-matched, though patients with CBF AML were significantly younger (p<0.01, t-test) with higher white blood cell count (p<0.01, t-test) and peripheral blast percentages (p<0.01, t-test) than *TP53*-mutated AML. One patient had a history of Li-Fraumeni syndrome, who developed *TP53*-mutated therapy-related MDS with excess blasts.

### WGS improves the ability to identify mutations in the TP53 gene

Recent evidence suggests that *TP53* mutant allele status is an important prognostic factor, with decreased survival in multi-hit *TP53*-mutated myeloid malignancies[4,5]. Based on standard clinical sequencing and cytogenetics, 93% of AML cases and 69% of MDS were classified as multi-hit *TP53* mutated cases (Table 1); all were confirmed by WGS. Five cases (12%) that initially appeared monoallelic or ambiguous were found to have additional *TP53* mutations via WGS analysis and are best described as multi-hit. Overall, all cases but one contained multi-hit *TP53* mutations, and one case was monoallelic. Multi-hit mutations predominantly occurred as a single SNV/indel combined either with copy loss of the wild-type *TP53* allele (22/42) or CN-LOH (11/42; Figure 1A). In 92% of these cases, there was sufficient supporting evidence to determine that the multiple *TP53* mutations were biallelic.

**Figure 1.**
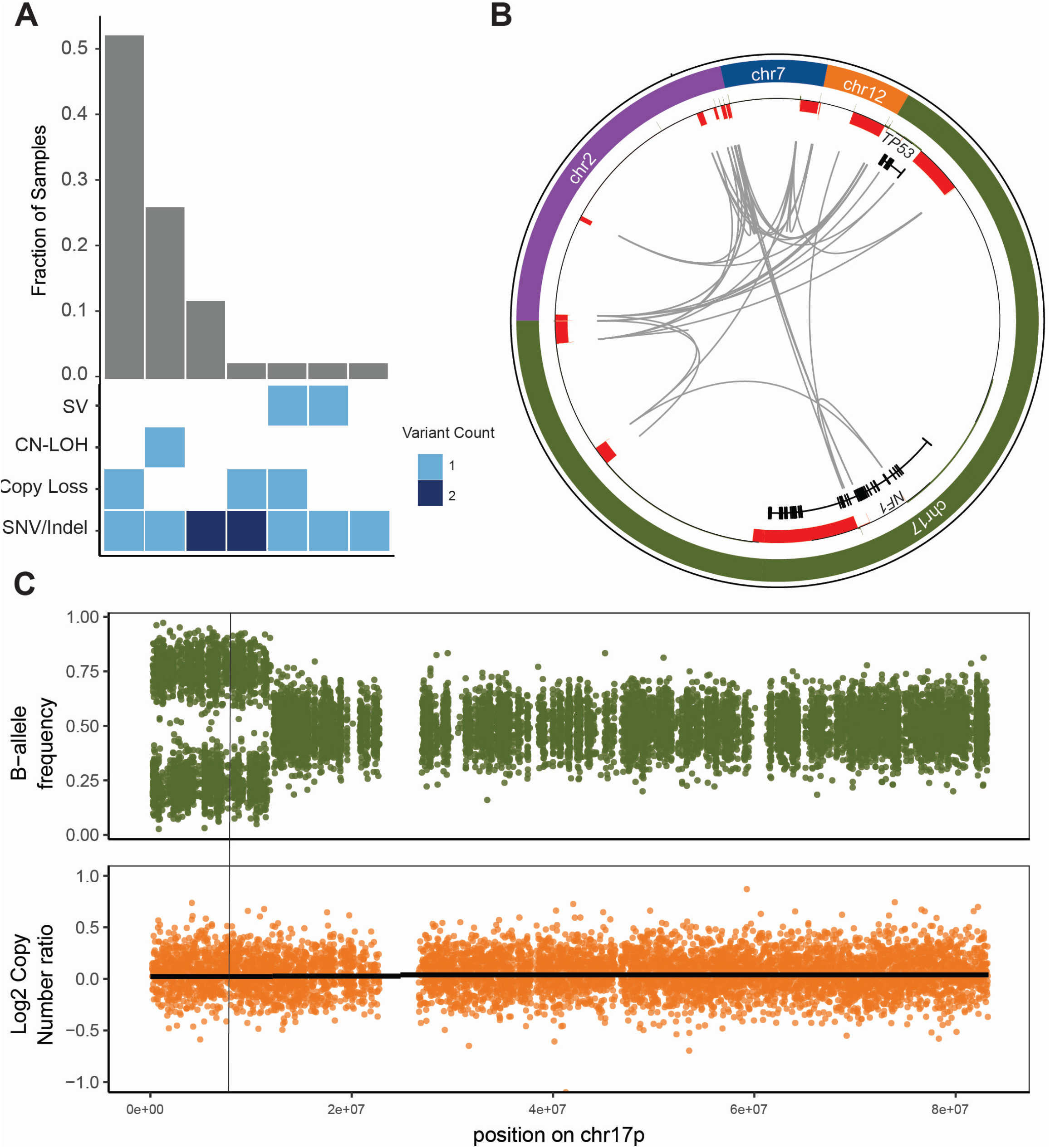
WGS results in the reclassification of some cases of *TP53*-mutated MDS/AML. A. Combinations of mutation types affecting *TP53* in AML/MDS cases, and corresponding proportion in cohort. B. Complex structural variant affecting the *TP53* genetic locus is detected using WGS based on resolution of rearrangements involving ch7, chr12, chr2, and *NF1* on chr17p. Copy number losses are shown in red. Gray arcs indicate regions linked by structural variants. C. Copy neutral loss of heterozygosity affecting chr17p is detected using WGS based on shifts in B-allele frequency (*top*) while log2 copy number ratio remains 0 (*bottom*). Vertical line indicates the position of the *TP53* locus.

Of the 5 cases reclassified from monoallelic to multi-hit, two examples including complex SV and CN-LOH are illustrated. UPN 480109 was previously classified as MDS with monoallelic *TP53* mutation based on panel sequencing; cytogenetics reported add(17)(q11.2) without 17p abnormality detected. By WGS, a complex SV was identified involving *TP53* and *NF1* on chromosome 17 as well as regions on chromosomes 2, 7, and 12 (Figure 1B). Reclassification to multi-hit *TP53* mutation increased the IPSS-M prognostic risk stratification[27] from high to very high. UPN 147444 was previously classified as MDS with monoallelic *TP53* mutation based on a *TP53* mutation VAF of 48.9% and lack of 17p abnormality on cytogenetics. WGS identified clear CN LOH on 17p (Figure 1C). Reclassification to multi-hit *TP53* mutation increased the IPSS-M stratification from moderately high to very high.

### TP53-mutated myeloid malignancies have a distinct pattern of somatic SNVs and Indels

The overall number of somatic point mutations and small indels was similar in *TP53*-mutated myeloid malignancies and CBF AML samples (Figure 2), and neither the mutation spectrum nor signatures suggested recurrent differences in mutational processes (Supplementary Figure 1). The trend towards an increased number of mutations in *TP53*-mutated cases may reflect the older age of this cohort compared to the CBF AML patients. This dataset confirms that cooperating mutations in *TP53*-mutated malignancies are distinct from those in other subtypes of AML, with an absence in this cohort of mutations in common signaling genes like *KIT* and *FLT3*, and also *WT1* [12,28] (Supplementary Table 3). Similarly, mutations in epigenetic modifiers (*DNMT3A, TET2*, or *ASXL1*) or spliceosome genes (*U2AF1, SF3B1, SRSF2*, or *ZRSR2*) are relatively uncommon in *TP53*-mutated myeloid malignancies in this cohort, with a cumulative frequency of 14.3% and 4.8%, respectively. Furthermore, *NRAS* or *KRAS* mutations were present in only two of 29 (7.1%) *TP53*-mutated AMLs versus 8 of 18 (44.4%) CBF AML (p = 0.009). We did observe enrichment for mutations in *NF1* in *TP53*-mutated AML cases. Single nucleotide variants (SNVs), consisting mostly of frameshift or stop gains, were detected in 5 of 29 cases (17.2%) and copy number loss was identified in 14 of 29 cases (48.3%). Of note, all five cases with SNVs also had a loss of the second *NF1* allele, suggesting that there is strong selection for multi-hit inactivation of *NF1* in *TP53*-mutated AML cases. As an exception, the single case with a monoallelic *TP53* mutation – a patient with MDS with excess blasts – had cooperating SNVs not seen in other cases, including *ASXL1, U2AF1* and *NRAS*, perhaps reflecting a distinct mechanism of transformation.

**Figure 2.**
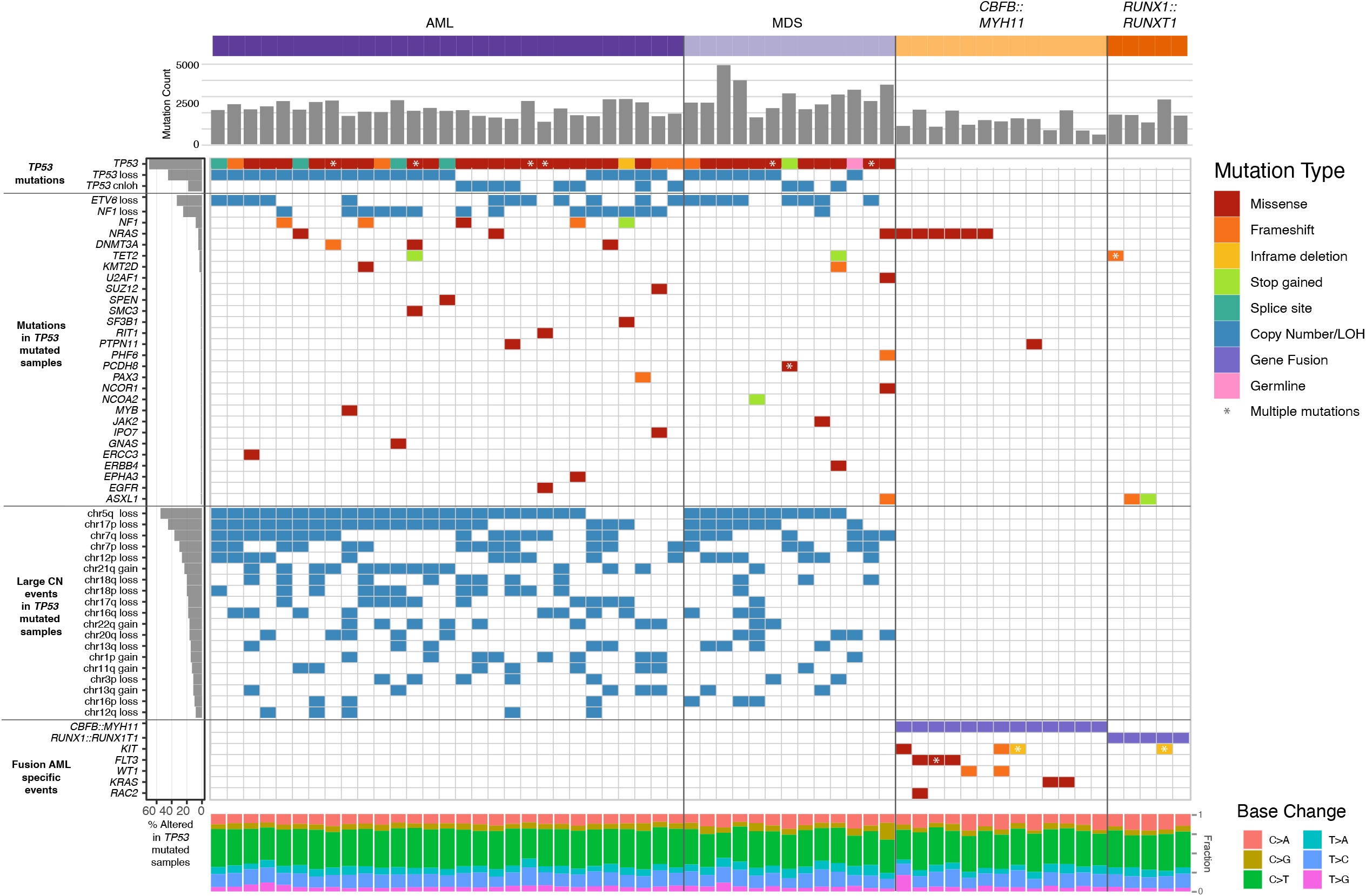
Mutational landscape of *TP53*-mutated myeloid malignancies. *TP53*-mutated AML (left) and MDS (middle), compared with AML driven by the *CBFB*::*MYH11* or *RUNX1*::*RUNX1T1* fusion proteins. Top: SNV/Indel counts for each sample. Left: frequency of selected genomic alterations in *TP53* samples. Middle: Colors represent the types of mutations observed in each sample; asterisks indicate multiple SNV/Indel hits. Bottom: Proportion of each single-nucleotide base change per sample.

### Specific chromosome alterations in TP53-mutated malignancies from different organs

Although aneuploidy is widespread in *TP53*-mutated myeloid malignancies, recurrent copy number changes were evident and consistent with well-established cytogenetic changes, specifically loss of chr5, chr17, chr7, chr 12, chr18, chr16, and chr20q, and gains in chr21, chr22, chr1p and chr8 (Figure 2, Supplementary Figure 2). A comparison to *TP53*-mutated solid tumors in the PCAWG dataset[9] demonstrated striking differences in the patterns of recurrent copy number changes (Figure 3). For example, loss of chr5q is present in >80% of the *TP53*-mutated myeloid malignancies in our study, compared to 58% and 38% of ovarian and esophageal adenocarcinomas, respectively, and fewer than 20% of any other tumor types. Similarly, chr7q loss is present in 56% of *TP53*-mutated myeloid tumors in our dataset, but infrequent in all other *TP53*-mutated PCAWG tumor types. Conversely, some recurrent copy number alterations that are common in other tumor types are only very rarely seen in *TP53*-mutated myeloid malignancies. For example, chromosome 6 loss is observed in <2.5% of *TP53*-mutated myeloid tumors but is often detected in *TP53*-mutated solid tumors including ovarian (83%), pancreatic (61%), and esophageal (29%) adenocarcinomas.

**Figure 3.**
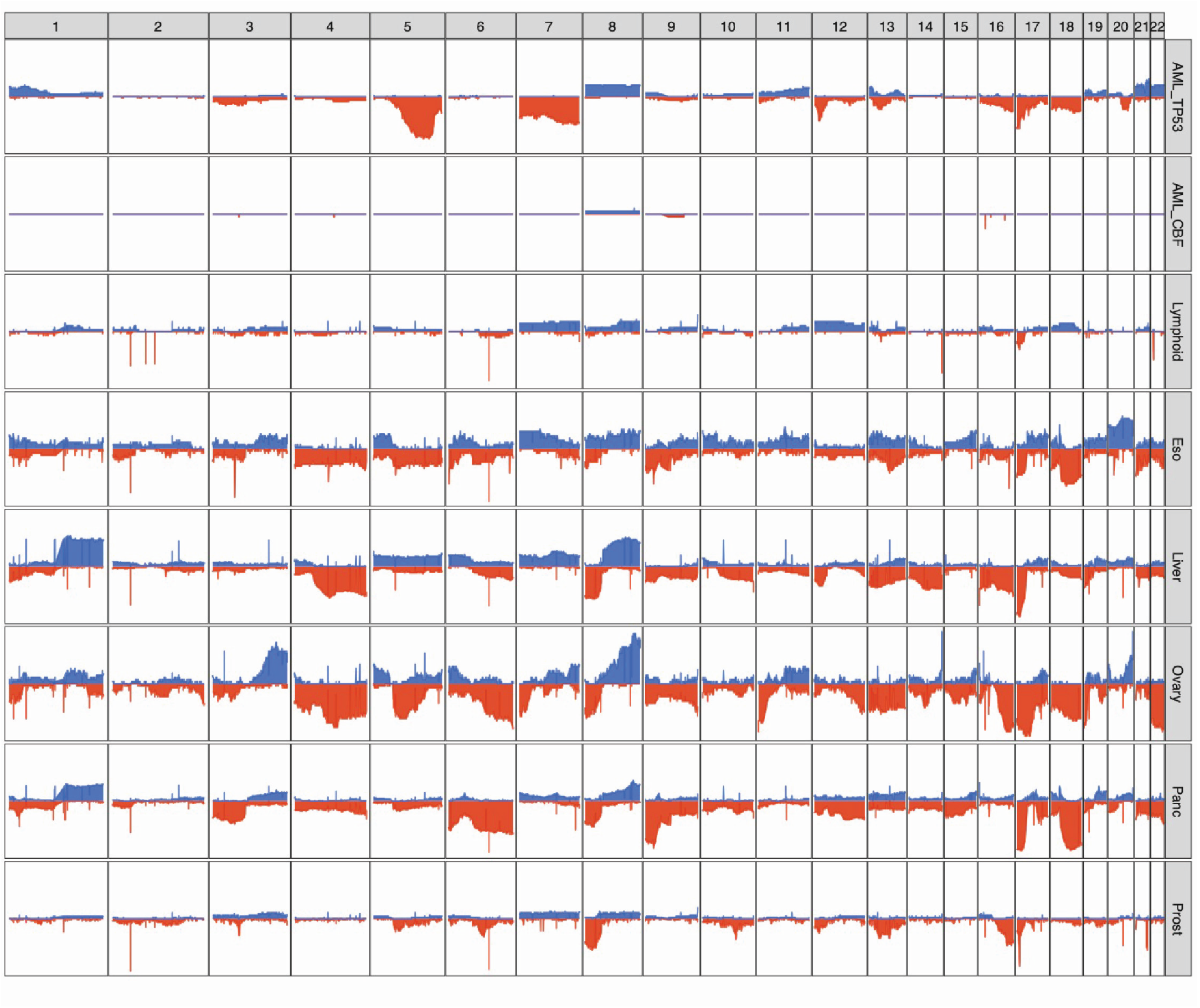
Distinct patterns of copy number alterations are observed in different *TP53*-mutated malignancies. Y-axis indicates the proportion of samples with a copy gain (blue, positive direction) or loss (red, negative direction) overlapping each gene, genome-wide. Only ‘mostly diploid’ samples without evidence of whole-genome doubling and with at least 50% of autosomes copy neutral are included. For the PCAWG data, all tumor types with at least 20 ‘mostly diploid’, *TP53*-mutated samples are included. Datasets included (*top to bottom*): *TP53*-mutated AML/MDS (N=41), *TP53*-wild type core-binding factor AML (N=18), and *TP53*-mutated lymphoid (B-NHL, N=20; CLL, N=6), esophageal (N=24), hepatic (N=56), ovarian (N=23), pancreatic (N=96), and prostate (N=43) malignancies.

Of particular interest are *TP53*-mutated lymphoid malignancies (i.e. non-Hodgkin lymphoma and chronic lymphocytic leukemia), which lack the recurrent copy number losses observed in *TP53*-mutated myeloid malignancies. These groups share only a high frequency of loss of the p-arm of chromosome 17, which encompasses the *TP53* locus itself.

### Structural variants are frequent and often complex in TP53-mutated malignancies

In *TP53*-mutated AML/MDS samples, we observed a median of 71 somatic SVs per sample (range: 2-576, Figure 4A, Supplementary Table 4), which were classified as follows: deletion (DEL), duplication (DUP), insertion (INS), inversion (INV), and break-end (BND). This represents a significantly higher burden than observed in the CBF AML samples (median=6, range=3-14, p=5.6 × 10^−8^, 2-sided Wilcoxon ranked-sums test). In the *TP53*-mutated samples, the majority of structural variants represented not simple events, but highly complex rearrangements (Figure 4B). Of 3,948 structural variants in *TP53*-mutated samples, 2,673 (68%) were part of a complex SV involving 20 or more individual SV calls. In the 42 *TP53*-mutated samples, we detected one or more complex events comprising >20 SVs in 26 samples (62%), and one or more complex events comprising >100 SVs in 8 samples (19%). Genome-wide, an analysis of SV breakpoints occurring in 100 kilobase windows showed clear enrichment of breakpoints at the inv(16) and t(8;21) loci for core-binding factor AML samples (Figure 4C). In contrast, breakpoints were identified throughout the genome in *TP53*-mutated AML/MDS cases, with enrichment on 17p (containing *TP53*) and on 21 (with one hotspot occurring just upstream of *RUNX1*).

**Figure 4.**
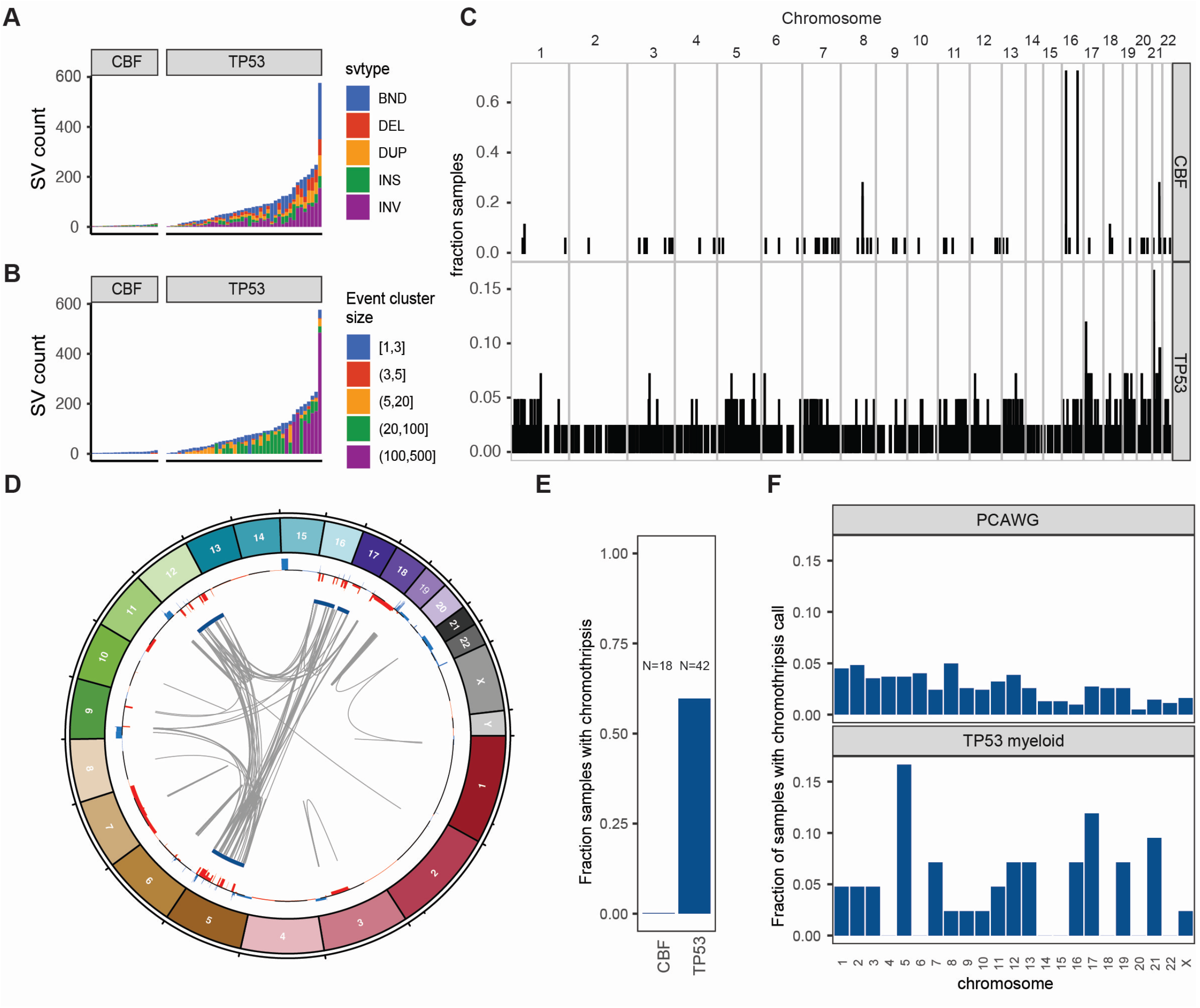
Structural variants and chromothripsis involve very large, complex events in *TP53*-mutated myeloid malignancies. A. Somatic structural variant call counts for each case. Counts colored according to SV classification: breakends (BND), deletion (DEL), duplication (DUP), insertion (INS), and inversion (INV). B. Distribution of somatic structural variant cluster sizes for each case. Counts colored according to the number of individual structural variant events occurring within the corresponding complex SV cluster. C. Frequency of somatic structural variant breakpoints in 100Kb windows genome-wide. Bar height indicates the fraction of samples with at least one SV breakpoint within each window. (*top*) Fraction of CBF AML with somatic SV breakpoints in each window. (*bottom*) Fraction of *TP53*-mutant AML with somatic SV in each window. In core-binding factor AML, enrichment of breakpoints is clearly present at inv(16) and t(8;21) positions. Breakpoints are present throughout the genome in *TP53*-mutated AML/MDS cases, with enrichment on chr17 and chr21. D. Large regions of chr5, chr12, chr16, and chr17 are involved in complex rearrangements affecting copy number, including the *TP53* genomic locus in a case of *TP53*-mutated AML (UPN 983349). *(outer track*) Copy gains (blue) and losses (red). (*inner track*) Regions involved in high-confidence chromothripsis events (dark blue). Gray arcs indicate novel adjacencies created by structural rearrangements. E. Chromothripsis is detectable in 60% of *TP53*-mutated myeloid malignancies and is not present in any core-binding factor AML cases. F. Genome-wide distribution of high-confidence chromothripsis calls. Proportion of all *TP53*-mutated AML/MDS (red; N=42) and PCAWG (blue; N=623) samples with a chromothripsis call on each chromosome.

Chromothripsis is common in myeloid malignancies with a complex karyotype, with a reported frequency of 35% based on SNP arrays[29]. Chromothripsis arises from the attempted repair of extensive double-strand break fragmentation across regions of one or more chromosomes. The definition of chromothripsis includes the following: (1) clustering of breakpoints with large regions of interleaving normal sequence; (2) oscillations between two or three copy number states; and (3) rearrangements of multiple fragments in random orientation and order[30]. Using ShatterSeek to define high confidence regions of chromothripsis[7], 25 (60%) of 42 *TP53*-mutated myeloid malignancies exhibited chromothripsis (Figure 4D, Supplementary Table 5). In contrast, chromothripsis was not observed in any CBF AML samples. Of the 25 *TP53*-mutated AML/MDS samples with at least one high-confidence chromothripsis region, nearly half (12/25; 48%) had high-confidence regions detected on more than one chromosome (See Figure 4E for an example). In *TP53*-mutated AML/MDS, chromothripsis events occurred most frequently on chr5 (7/43 chromothripsis events), chr17 (5/43 chromothripsis events), and chr21 (4/43 chromothripsis events), a pattern distinct from that observed across 623 *TP53*-mutated samples from the PCAWG consortium[7], in which 408 high confidence chromothripsis regions were reported in 246 samples (38.2%; Figure 4F) We note that the seven samples with chromothripsis within chr5 were a subset of the 33 samples with 5q loss, and do not represent biallelic loss of 5q.

### Haploinsufficiency for ETV6 due to genetic loss or epigenetic silencing is frequent in TP53-mutated myeloid malignancy

Copy number losses on the p-arm of chromosome 12 occur frequently in *TP53*-mutated myeloid malignancies, affecting 19 (45%) of 42 samples in this cohort (Supplementary Tables 6 and 7). The minimally-deleted region of 12p spans 2.75Mb, and includes the gene bodies of 18 protein-coding genes, including the tumor suppressors *ETV6* and *CDKN1B* (Figure 5A). Of the 18 protein-coding genes intersecting the minimally deleted regions, mRNA expression was analyzed (Figure 5B). Only one, *ETV6*, exhibited high expression in CBF and NK AMLs and a significant difference in expression in *TP53*-mutated AML with 12p loss (adjusted p-value=1.0×10^−4^, Figure 5C, Supplementary Table 8). The other gene notable for significantly decreased expression in the region was *DDX47* (adjusted p-value=4e^-8^), but with less expression in CBF and NK AMLs, as well as unclear function in hematopoietic cells. *ETV6* also was frequently deleted in *TP53*-mutated samples in the Beat AML cohort[28] with loss of one copy of *ETV6* in 6/19 (32%) cases, compared to 4/238 (1.7%) of non-*TP53* mutated primary AML samples (OR=26, 95% CI=(5.4, 141.9), p=1.3×10^−5^, 2-sided Fisher’s exact test, Supplementary Table 9). Of note, somatic coding SNVs or indels in *ETV6* were not detected in any *TP53*-mutated myeloid malignancies in this study, or the BEAT AML cohort.

**Figure 5.**
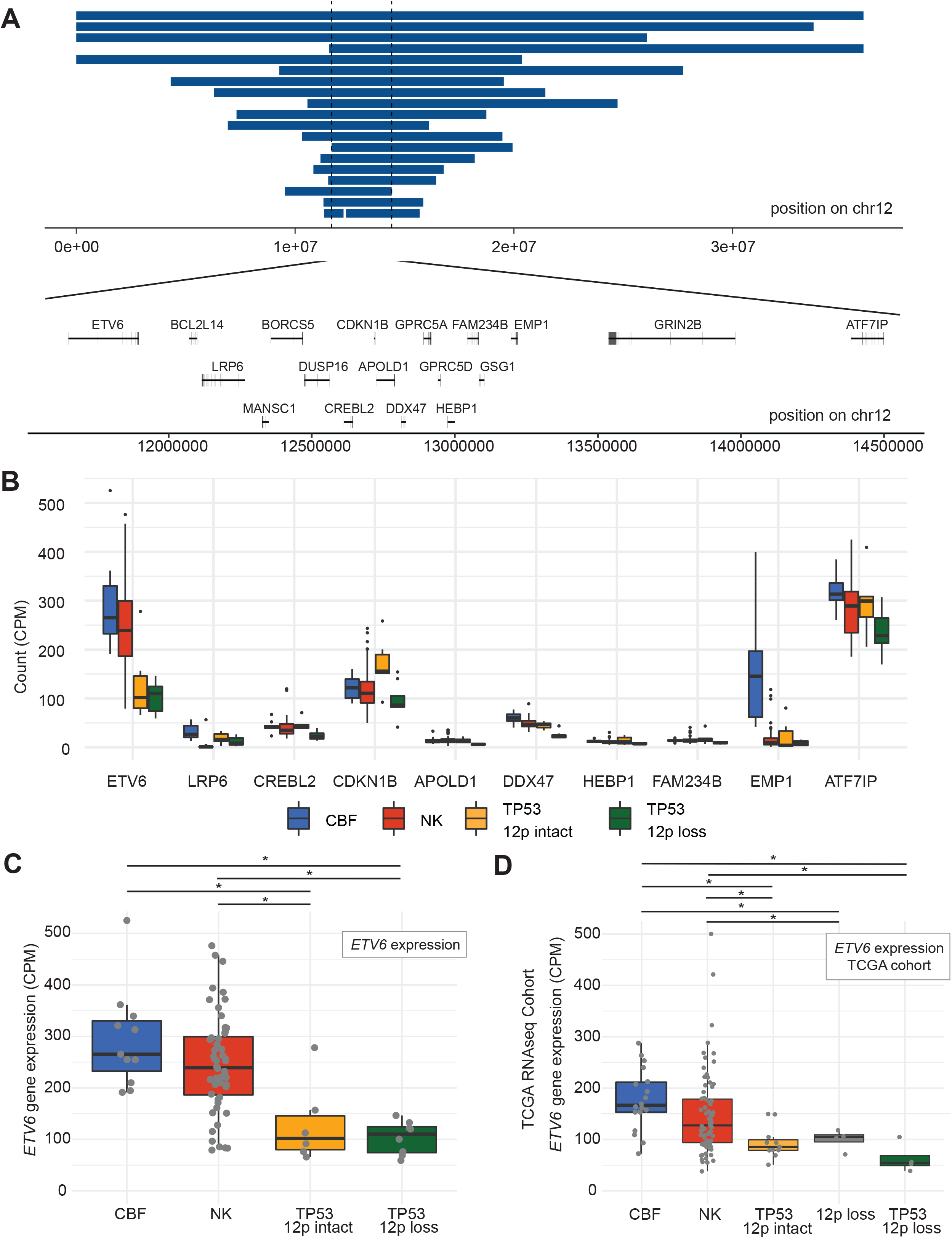
*ETV6* deletion and decreased expression is common in *TP53*-mutated myeloid malignancies. A. Genomic locations of copy number losses intersecting the minimally deleted region on chr12p (N=19). Locations of all protein-encoding genes intersecting this region are shown. B. mRNA expression for genes within the minimally deleted region of chr12p, displayed for genes with a minimum mean expression of 10 CPM for at least one group. Boxplot indicates median and first and third quartiles; whiskers indicate the range of all data points falling within 1.5*IQR (interquartile range). CBF, core-binding factor AML (N=11); NK, normal karyotype AML (N=52); *TP53*-mutated with 12p intact (N=6); *TP53*-mutated with 12p loss (N=8). C. mRNA expression of *ETV6* in *TP53*-mutated samples with and without 12p loss, compared to NK and CBF AML cases (N=8, 6, 52, and 11, respectively). * Adjusted p-value < 0.05, edgeR exactTestDoubleTail with adjustment for multiple comparisons. D. Extension cohort of additional cases from the previously published TCGA AML data set. mRNA expression of *ETV6* in *TP53*-mutated samples with 12p loss and with 12p intact (N=4 and 11, respectively), NK (N=79), CBF (N=18), and 12p loss (*TP53* wildtype, N=4) samples. * Adjusted p-value < 0.05, t-test with multiple testing correction.

Surprisingly, decreased *ETV6* expression also was observed in *TP53*-mutated myeloid malignancies *without* 12p structural rearrangements (adjusted p-value=0.021, Figure 5C), and was confirmed in an extension cohort of mRNA data from the TCGA[12] (p-value < 0.02 and 0.002 compared to CBF and NK AML cases, respectively; Figure 5D), suggesting an epigenetic mechanism that reduces expression of this gene. No regulatory mutations in the *ETV6* promoter or nearby enhancers were detected, nor was *ETV6* promoter methylation altered in a cohort of 29 previously published samples with whole-genome bisulfite sequencing[13]. Further studies will be needed to understand the mechanisms underlying these observations.

### Telomere content is increased in TP53-mutated myeloid malignancies

Telomere length is decreased in most cases of AML or MDS, presumably due to increased replication of leukemic blasts[31–34]. We estimated telomere content using two independent approaches (TelomereHunter[22] and Telseq[23]), which were highly correlated (R^2^=0.88, Supplementary Figure 3). Here, we report results from TelomereHunter. As expected, telomere content was reduced in CBF AML bone marrow compared with matched normal tissue (skin or buccal swab), with a mean shortening of 226 TRPM (telomeric reads per GC content-matched million reads; p=8.3×10^−7^; Figure 6A; Supplementary Table 10). In contrast, telomere content in *TP53*-mutated myeloid tumors was similar between leukemic and matched normal tissues. Compared to CBF AML, telomere content in *TP53*-mutated AML/MDS was significantly increased (p=0.0068), despite the increased average age of the *TP53*-mutated AML/MDS patients. Indeed, after normalizing the tumor telomeric content for each sample to its paired normal as an internal control, we observed significantly less telomere shortening in *TP53*-mutant AML/MDS compared to CBF AMLs (p=1.7×10^−5^; Figure 6B). An analysis of telomere variant repeats (TVR) in singleton context (i.e., variant hexamers flanked by at least 3 t-type hexamers to each side) showed that telomeric repeats containing TTTGGG hexamers were significantly increased in *TP53*-mutated AML/MDS compared to the CBF subset (p=1.4×10^−7^; Figure 6C).

**Figure 6.**
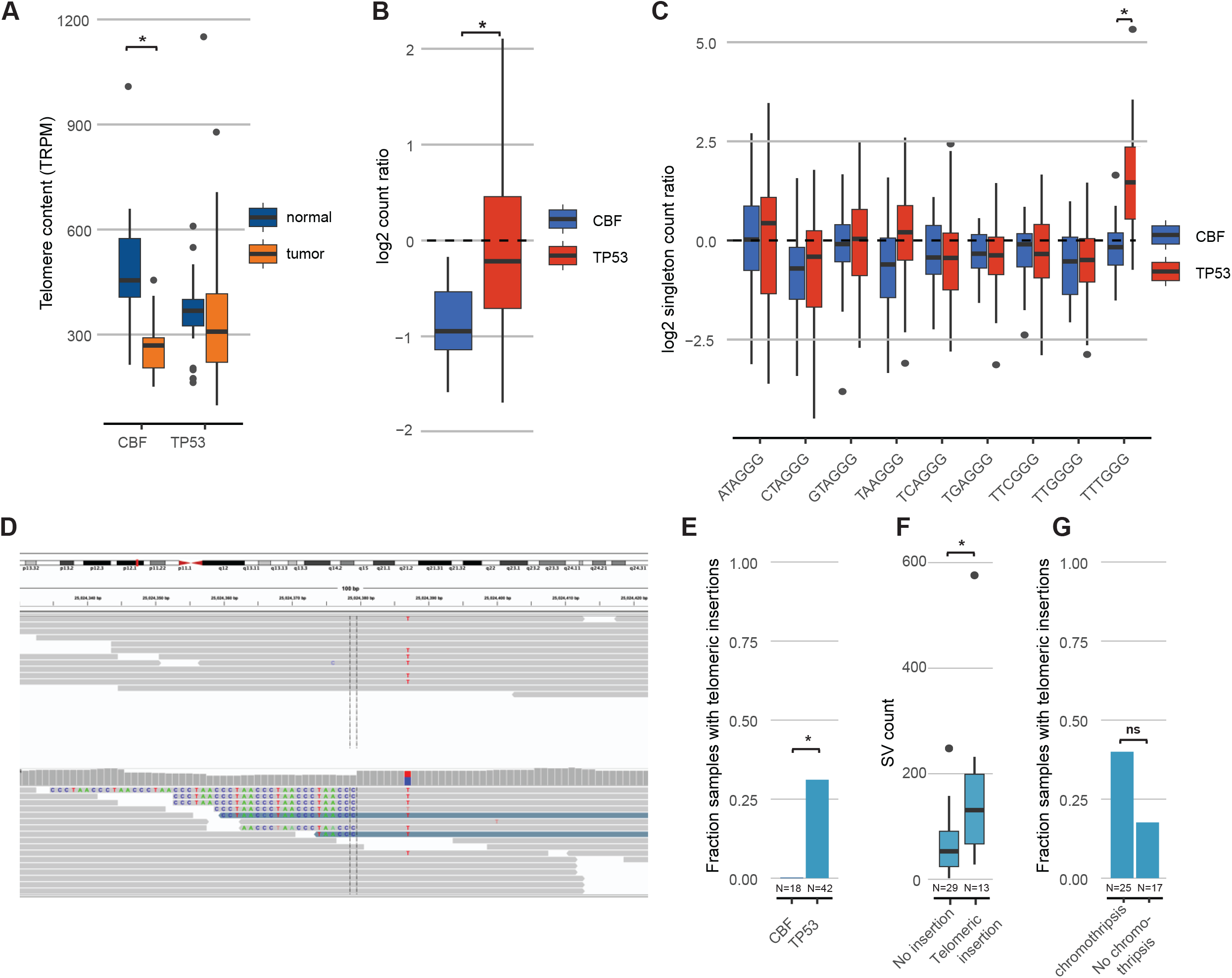
Telomere content is increased in *TP53*-mutated myeloid malignancies A. Telomere content in units of telomeric reads per GC content-matched million reads (TRPM) was quantified in WGS from tumor specimens (*orange*) and paired germline control tissues (skin or buccal DNA, *blue*) for core-binding factor AML (CBF) and *TP53*-mutated myeloid malignancies. Significant telomere shortening in the CBF sample subset, with a mean shortening of 226 TRPM (telomeric reads per GC content-matched million reads; p=8.3×10^−7^; 2-sided, paired t-test) but not in the *TP53*-mutant subset (mean difference of -2.5 TRPM between tumor and normal) B. Ratio of telomere content for tumor compared to normal tissue. *TP53*-mutated myeloid malignancies have significantly higher telomere content compared to CBF AML (p=1.7×10^−5^, 2-sided student’s t-test). C. Relative abundance of TVR repeats in singleton context. Log2 tumor/normal ratio of singleton TVR repeats in *TP53*-mutant and CBF subtypes. The relative abundance of singleton TTTGGG repeats is significantly higher in *TP53*-mutated myeloid malignancies compared to CBF AML (p=1.4×10^−7^, 2-sided student’s t-test). D. Example of intrachromosomal insertion of t-type telomeric hexamer sequences seen in the tumor but not the paired normal sequence data from a *TP53*-mutated case (UPN 387082). E. Fractions of *TP53*-mutated myeloid malignancies (N=42) and CBF AML patients (N=18) with detectable interstitial (non-telomere) telomeric repeat variants (p=0.006, Fisher’s exact test) *F. TP53*-mutated cases with detectable interstitial insertions of telomeric variant repeats have higher structural variant counts (p=0.0033, Wilcoxon ranked-sums test). G. Fraction of *TP53*-mutated cases with detectable interstitial insertions of telomeric variant repeats in sample with (N=25) and without (N=17) chromothripsis (p-value not significant, odds ratio=3.03, 95% CI=(0.61, 20.7), Fisher’s exact test).

In addition to the differences in relative telomere content, insertions of telomeric repeats into intrachromosomal regions (example shown in Figure 6D; Supplementary Table 11) were detected in 13/42 (31%) *TP53*-mutated AML/MDS tumor samples but none of the CBF tumor samples (p=0.0061, Fisher’s exact test; Figure 6E). Nine of these 13 samples contained a single telomeric insertion; the remaining four contained two or more insertion sites. Within the subset of *TP53*-mutated samples, these telomeric insertions were associated with higher overall SV counts (p=0.0033, Wilcoxon ranked-sums test, Figure 6F) and showed a trend towards the presence of chromothripsis that was not statistically significant (Figure 6G). The telomeric insertions tended to occur in close proximity to structural variations: 5/19 (26%) occurred within 10 kb of a high-confidence chromothripsis region, and 13/19 (68%) occurred within 10 kb of an SV breakpoint.

Notably, we did not observe any somatic SNV, indels, or SV affecting either the *TERT* gene body and promoter region, or the genes *ATRX* or *DAXX*, which have been implicated in the ALT pathway of telomere maintenance. We did observe copy number events affecting the *TERT* locus on chr5p, resulting in amplification in 2 samples and a single copy loss in 2 samples (Supplementary Table 6). However, no significant difference in *TERT* mRNA expression was observed in TP53-mutated AML/MDS samples; similarly, *ATRX* and *DAXX* did not significantly differ in expression (Supplementary Figure 4).

## Discussion

This study represents the largest WGS characterization of *TP53*-mutated myeloid malignancies described to date. The complexity and frequency of structural variants and copy number abnormalities makes genomic characterization of these cases difficult when using targeted sequencing, karyotyping, FISH panels and/or array CGH. In this study, complex structural variants were resolved using matched normal control DNA to facilitate the definitive identification of acquired structural variants [35]. As a result, high confidence structural variant calls could be used to assess the frequency, size, and complexity of rearrangements in these very disordered genomes.

Multi-hit involvement of the *TP53* locus was frequent (94% in this cohort), and 92% of multi-hit cases could be confirmed as biallelic. WGS analysis identified multi-hit *TP53* mutations in 5 cases (12% of the cohort) that were previously ambiguous, or that appeared to be monoallelic, based on cytogenetics and panel sequencing. The importance of multi-hit *TP53* mutations has been recently emphasized[4,5] and included in the current International Consensus Classification of Myeloid Neoplasms and Acute Leukemias[3], but the requirement for multifaceted approaches to accurately identify multi-hit mutations at the *TP53* locus has been an ongoing dilemma. Multi-hit mutations in *TP53* are reported to occur in 76% (N=174/230) of cases with any *TP53* mutation in AML/MDS-EB[5] and 63% (N=253/402) in MDS[4], and supplemental approaches were necessary to clarify multi-hit status by LOH mapping in both studies. For example, copy neutral loss of heterozygosity is not detectable by conventional cytogenetics, and requires SNP analysis using arrays or comprehensive sequencing panel testing[36]. Reliance on the variant allele frequency of *TP53* mutations above a threshold of 50%[3] or 55%[5] missed 12% of AML/MDS with multi-hit *TP53* in this study, and up to 25% of MDS cases with CN-LOH in the previous study[4]. Furthermore, these VAF cutoffs may not be transferable to peripheral blood samples or to bone marrow aspirate specimens that are hemodilute. The results of this study emphasize the utility of WGS to accurately determine the allele status of *TP53*-mutated myeloid malignancies.

Chromosomal patterns of both aneuploidy and chromothripsis differed remarkably in *TP53*-mutated myeloid cancers versus lymphoid malignancies or solid tumors. In *TP53*-mutated myeloid malignancies, recurrent copy number loss, structural variants, and chromothripsis often involve large regions on chr5 and chr7 that are well established through conventional cytogenetics and other studies[4]. Indeed, chromothripsis in *TP53*-mutated AML with complex karyotype has previously been noted (using SNP array profiling) to affect the entire genome, most prevalently on chromosomes 3, 5, 7, 12, 16, and 17[29]. Interestingly, distinct patterns of chromosome loss or gain were observed in different *TP53*-mutated cancer types. Notably, *TP53*-mutated lymphoid malignancies have a distinct pattern of aneuploidy compared to *TP53*-mutant myeloid malignancies. These data suggest a model in which genomic instability induced by *TP53* mutations selects for chromosomal alterations that are tolerated and potentially also increase cell fitness, a process that may be highly dependent on the cell of origin. Consistent with this model, prior studies have suggested that tissue-specific mutation profiles may arise due to the sensitivity of the cell of origin to disruptions in particular transcription factors and signaling networks [37,38] or due to chromosome architecture and transcriptional activity[39].

One of the most frequently mutated genes in *TP53*-mutated myeloid malignancies is *NF1*. Somatic SNVs, indels, and/or copy number loss of *NF1* were detected in 48% of *TP53*-mutated AML cases in our cohort, with multi-hit mutations in 17% of cases. Prior studies have shown that *NF1* mutations are present in 7% of all AML cases[40] and further enriched in complex karyotype AML, ranging from 9-19% using gene panel sequencing[41,42] to 22.7% using a SNP-array based method, with approximately one-third of these cases carrying multi-hit deletions[43]. NF1 is a negative regulator of Ras signaling. Although activating mutations of *NRAS* and *KRAS* are frequently observed in other types of AML, they are relatively uncommon in *TP53*-mutated myeloid malignancies. A reason for the preference for NF1 inactivation rather than activating Ras mutations in *TP53*-mutated AMLs is not clear and will require additional studies to define the mechanism(s) involved.

Deletion of the p-arm of chromosome 12 was identified in 45% of our cases with *TP53*-mutated myeloid malignancy. Analysis of the minimally deleted region and gene expression identified *ETV6* as the most likely target gene in this region. This is consistent with a prior study that reported 12p deletions in 39 of 79 (49%) patients with complex karyotype AML, and identified *ETV6* to lie within the minimally deleted region[44]. In contrast, non-complex karyotype AML/MDS has a much lower incidence of somatic mutation of *ETV6*. Structural variants in *ETV6* as identified by cytogenetics and interphase FISH have been reported in 1.1% of 3,798 AML cases and 0.2% of 3,375 MDS cases[45]; similarly, somatic SNVs/indels were reported in 1.4% of AMLs and 3.0% of MDS cases among a cohort of 970 patients[46]. Surprisingly, decreased *ETV6* mRNA expression was observed in *TP53*-mutated myeloid malignancies – with or without 12p loss. Although the number of cases analyzed is small, these observations suggest that *ETV6* haploinsufficiency, either through gene deletion, epigenetic silencing, or another mechanism, is relevant for the pathogenesis of *TP53*-mutated myeloid malignancies.

Telomeres are ribonucleotide structures that cap the end of chromosomes, playing an essential role in genomic stability. Telomeres shorten with each mitotic cell division[47], and multiple prior studies have demonstrated shortened telomeres in AML and MDS cells[31–34]. Consistent with these reports, telomere content (as measured by WGS) is decreased in CBF AMLs relative to matched normal tissue. In contrast, no significant decrease in telomere content was observed in *TP53*-mutated myeloid malignancies, and a similar observation has been reported in a preliminary study [48]. Telomeres are maintained by two major pathways: 1) a telomerase-dependent pathway, and 2) ALT, a recombination-dependent pathway to extend telomeres. We observed an increase in the TTTGGG telomere variant repeat in *TP53*-mutated myeloid malignancies, which has been associated with a telomerase-dependent mechanism of telomere maintenance[8]. However, copy number amplifications involving *TERT* were observed in only 2 samples, and no mutations in *TERT* or its promoter were detected; this finding is in stark contrast to solid tumors in the PCAWG consortium that showed extensive structural variants and copy number amplifications affecting *TERT* expression[10]. Moreover, no consistent increase in *TERT* mRNA expression was observed in *TP53*-mutated AML versus CBF AML. Somatic insertions of telomere sequences into non-telomeric DNA were identified in approximately one-third of our *TP53*-mutated myeloid malignancies cases; this likely is an underestimate, since the short-read WGS approach used in our study is not well suited to identify these alterations. A prior study reported that Interstitial insertion of telomere sequences is associated with ALT telomere maintenance[8]. However, no truncating mutations of *ATRX* and *DAXX*, which are associated with ALT tumors, or loss of expression of these genes was observed in our cohort of *TP53*-mutated myeloid malignancies. Thus, the mechanisms by which telomeres are maintained in *TP53*-mutated myeloid malignancies are unclear.

In summary, WGS of AML/MDS with paired normal tissue provides the most accurate characterization of the genomic landscape of *TP53*-mutated myeloid malignancies. These data emphasize the unique features of these cancers, including the high frequency of chromothripsis and structural variants, and the involvement of unique signaling genes, including *NF1* and *ETV6*. Moreover, we provide evidence that telomere maintenance is altered in *TP53*-mutated myeloid malignancies. These pathways may represent potential new targets for therapeutic intervention.

## Data Availability

The datasets generated during and analyzed during the current study are deposited in the dbGap repository, in study phs000159. The supplementary tables for this study are deposited in Zenodo repository 
doi: 10.5281/zenodo.7523559.

https://doi.org/10.5281/zenodo.7523558

## Acknowledgements

Research support includes the Genomics of Acute Myeloid Leukemia Program Project grant NCI P01 CA101937 (D.C.L and T.J.L.), Specialized Program of Research Excellence in Leukemia NCI P50 CA171963 (D.C.L.), NCI K12 CA167540 (K.A.O. and R.B.D.), NCI R50 CA211782 (C.A.M), and American Society of Hematology Scholar Award (K.A.O.). Core services were provided by the Alvin J. Siteman Cancer Center Tissue Procurement Core and Biostatistics Shared Resource Core through an NCI Cancer Center grant (P30CA091842) and by the Cytogenomics and Molecular Pathology Laboratory, Genome Technology Access Center, and the McDonnell Genome Institute at the Washington University School of Medicine. The content is solely the responsibility of the authors and does not necessarily represent the official views of the NIH.

## Author Contributions

HJA, KAO, CAM, DCL, TJL, MJW, and JFD designed the study. Tissue repository sample identification and clinical annotations were performed by SEH, PW, RBD, SPT, and JEP. Data acquisition was performed by RSF, CCF, NMH, DHS, EJD, and MCS. Data analysis was performed by HJA, CAM, KAO, SMR, SNS, DHS, EJD, and MCS. HJA, KAO, CAM, DCL, and TJL wrote the manuscript draft, and all authors reviewed and contributed to the final manuscript.

## Data Availability

The datasets generated during and analyzed during the current study are available in the dbGap repository, in study phs000159.

## Supplementary Materials

The supplementary tables for this study are deposited in Zenodo repository doi: 10.5281/zenodo.7523559

## Supplementary Methods. Detailed methods and references for data analysis

**Supplementary Table 1.** WGS coverage depth metrics and estimated tumor purity (xlsx)

**Supplementary Table 2.** Clinical and molecular details (xlsx)

**Supplementary Table 3.** Somatic SNV and indel calls (tsv)

**Supplementary Table 4.** Somatic SV calls (tsv)

**Supplementary Table 5.** Chromothripsis events (xlsx)

**Supplementary Table 6.** Gene-based somatic copy-number variant calls (tsv)

**Supplementary Table 7.** Somatic copy-number variant calls (tsv)

**Supplementary Table 8.** RNA expression data with differential gene expression calculated for *TP53*-mutated cases compared to combined NK and CBF cases. (tsv)

**Supplementary Table 9.** CNV analysis of *TP53*-mutated cases in Beat AML cohort (tsv)

**Supplementary Table 10.** Telomere length estimates (xlsx)

**Supplementary Table 11.** Telomeric insertion calls (xlsx)

**Supplementary Figure 1.** Mutation signatures

**Supplementary Figure 2.** Circos plots

**Supplementary Figure 3.** Telomere content agreement between methods. Telomere content by patient age.

**Supplementary Figure 4.** mRNA expression of *TERT, ATRX*, and *DAXX*

