## Supplementary Methods for "Genomic landscape of *TP53*-mutated myeloid malignancies"

### SNV calling

SNVs and small indels were detected using MuTect2[1], VarScan2[2], Strelka2[3], and GATK[4]. Variants with a population frequency in gnomAD of greater than 0.1% were removed[5], as were those in regions of low-quality mapping (< 10% of reads with MQ0), low coverage (< 20X), and those called by Varscan only. Variant annotation was performed with the Variant Effect Predictor[6], version 95. Manual review was performed to remove further artifacts and to recover low variant allele frequency (VAF) variants in known AML driver genes with at least three supporting reads of evidence. Additional artifacts were excluded by removing calls that occurred at a homopolymer tract and had at least two different indels called at that same location. The entire somatic pipeline is available as a CWL workflow at https://github.com/genome/analysis-workflows (commit URL: https://github.com/genome/analysis-workflows/tree/0952c3f53a5eceaa32a2b8c1e974da79e01b76de).

### Structural variant and CNV calling

Structural variant and CNV calling were performed using the GRIDSS-Purple-Linx pipeline as described (https://github.com/hartwigmedical/gridss-purple-linx/blob/master/gridss-purple-linx.sh) using as input the aligned tumor and normal bam pairs, the filtered, high-confidence somatic SNV/indel calls and the following tool versions: GRIDSS_VERSION=2.9.4, GRIPSS_VERSION=2.1, AMBER_VERSION=3.9, COBALT_VERSION=1.13, PURPLE_VERSION=3.4.1, LINX_VERSION=1.19.

Following an initial run of the pipeline, purity estimates from Purple, followed by manual review (Supp Table 1), were used as input parameters to the GRIPSS filtering and downstream tools. SV were filtered for FILTER status=PASS and length>50bp.  Linx was used for clustering complex variants.  Fractional absolute copy number calls from Purple were rounded to the nearest integer value for classification as ‘gain’ or ‘loss’. Copy-neutral loss-of-heterozygosity was called using the MinorAlleleCopyNumber estimate provided by Purple.  All mutations affecting *TP53* were manually reviewed.

### Chromothripsis detection

Chromothripsis detection was performed using Shatterseek (https://github.com/parklab/ShatterSeek, commit 4b8b41011ecfe6d1496e906e5d9ec7d65467d476), using the filtered SV (DEL, DUP, INV, and BND types only) and copy number outputs from Purple as input, and default parameters for Shatterseek. Filtering for high-confidence chromothripsis regions was performed as recommended in the Shatterseek documentation, and was followed by manual review to exclude false-positive calls.

### Comparison of recurrent cell-type specific recurrent CNV

Comparison to the PCAWG per-gene copy number calls were performed using copy number estimates downloaded from https://dcc.icgc.org/api/v1/download?fn=/PCAWG/consensus_cnv/gene_level_calls/all_samples.co nsensus_CN.by_gene.170214.txt.gz. From the set of ICGC public dataset, we selected the 623 samples identified as having a *TP53* driver mutation (https://dcc.icgc.org/api/v1/download?fn=/PCAWG/driver_mutations/TableS3_panorama_driver_mutations_ICGC_samples.public.tsv.gz) irrespective of allelic status or mutation type. Samples reported to have undergone whole genome duplication were excluded (based on https://dcc.icgc.org/api/v1/download?fn=/PCAWG/consensus_cnv/consensus.20170217.purity.ploidy.txt.gz), and samples for which fewer than 50% of autosomal genes were estimated copy neutral were also excluded. We combined the PCAWG tumor type classifications Lymph-BNHL and Lymph-CLL to form a single 'Lymphoid' malignancy tumor type, and then restricted our analysis to tumor types with at least 20 samples meeting the above criteria.

**Estimation of telomere content**

Telomere content was estimated using TelomereHunter (v1.1.0)[7] in tumor-normal mode, using default parameters, and Telseq (v0.0.1)[8] separately for tumor and paired normal samples. Estimates of telomere content from the two methods were highly correlated ($R^2$=0.88), so we focused on just the results of TelomereHunter. Analyses of TVR (telomere variant repeats) in singleton context (i.e., flanked by at least 3 t-type telomeric hexamers to either side) were based on the per-sample tumor/normal ratio of normalized singleton read counts, as provided by TelomereHunter.

**Identification of intrachromosomal telomeric insertions**

Identification of intrachromosomal insertions of telomeric repeats was performed following the approach previously described[9]. Using the telomeric reads identified by TelomereHunter (i.e., with at least six t-type, c-type, g-type or j-type hexameric repeats), we identified reads such that only one member of the pair was classified as telomeric. We then identified candidate insertion regions as 1Kb windows containing 3 or more of these 'orphaned' telomeric reads in the tumor and none in the paired normal sample, excluding assembly gaps and the terminal cytoband of each chromosome. Within these candidate regions, we identified soft-clipped reads with mapping quality>30, excluding duplicates, secondary, and supplementary reads, where at least one t-type, c-type, g-type or j-type hexamer was present in the soft-clipped region. We identified all positions at the site of clipping in 4 or more reads from the tumor, followed by filtering of sites within segmental duplications or simple repeats, sites with the presence of soft-clipped telomeric repeats in the paired normal, and sites identified in more than 2 samples. Finally, all candidate insertions were manually reviewed to exclude false positives.

**Copy Number Analysis of BeatAML cohort**

Copy number analysis in the BeatAML cohort[10] was performed using all primary AML cases and all available normal controls. In instances where more than one sample per primary AML case was provided, we chose one sample at random, which preference for bone-marrow samples when available. Copy number analysis was performed using cnvkit (v0.9.8)[11] and with masking of assembly gaps and centromeric regions, using the full set of normal samples as a panel of normal, and according to the cnvkit authors' recommended workflow. In order to account for observed systematic noise due to, e.g., differences in GC-content between adjacent genomic bins, we re-centered the log2 copy number ratio in each bin by subtracting the median log2r ratio across all tumor samples, and then performed a second round of copy number segmentation.
