## Supplementary figures and images for "Genomic landscape of *TP53*-mutated myeloid malignancies"

### Supp_Fig_2_circos_Page_01.jpg

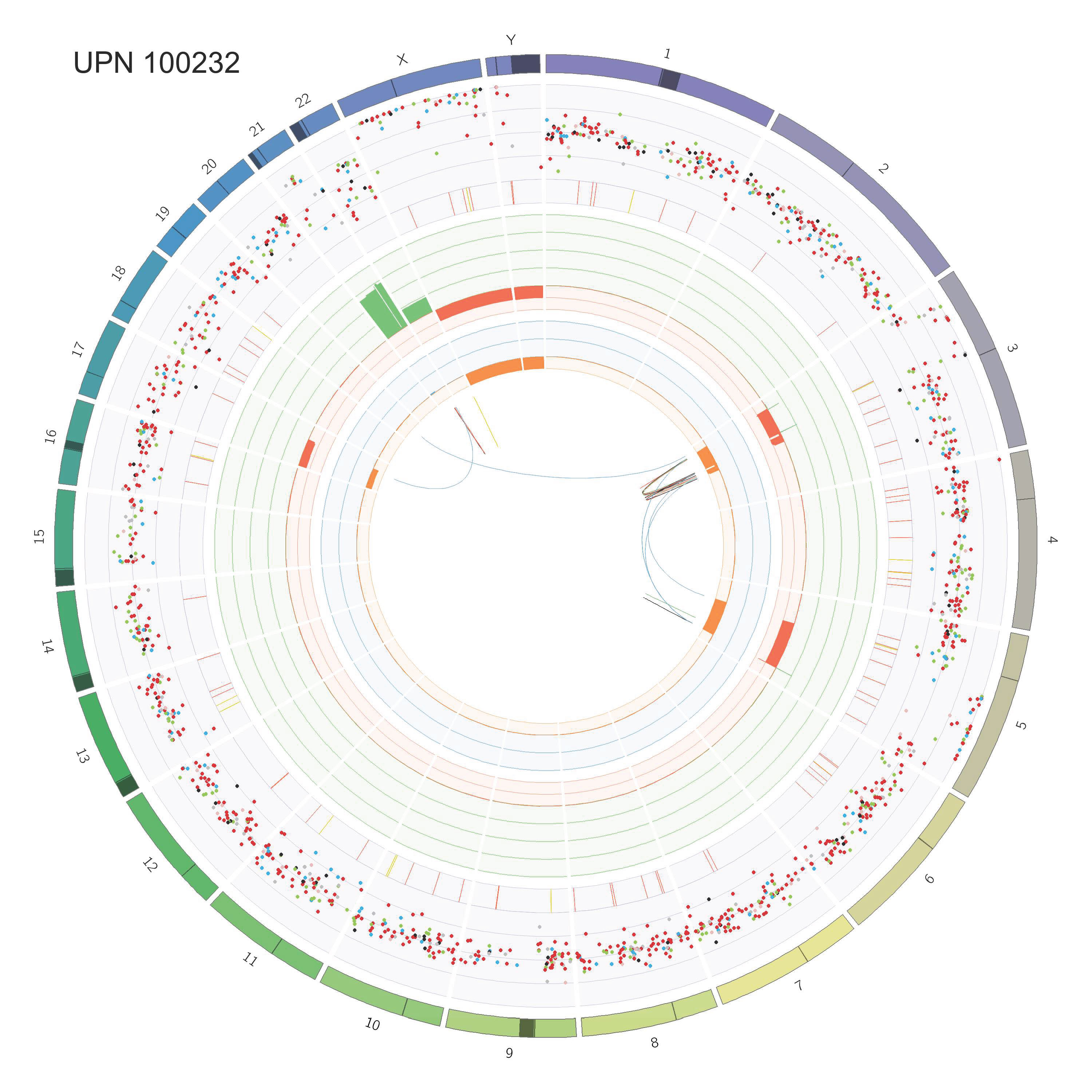

### Supp_Fig_2_circos_Page_02.jpg

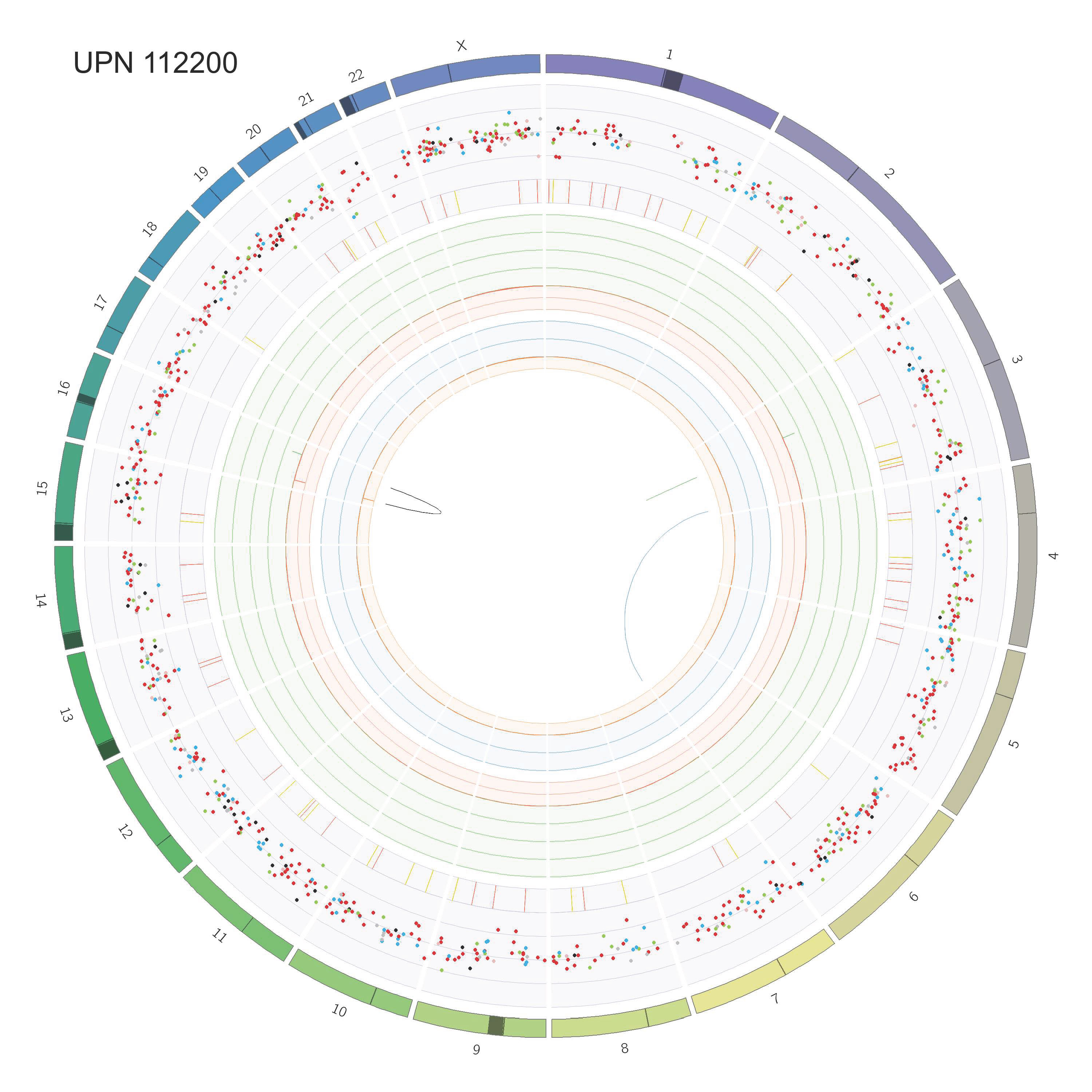

### Supp_Fig_2_circos_Page_03.jpg

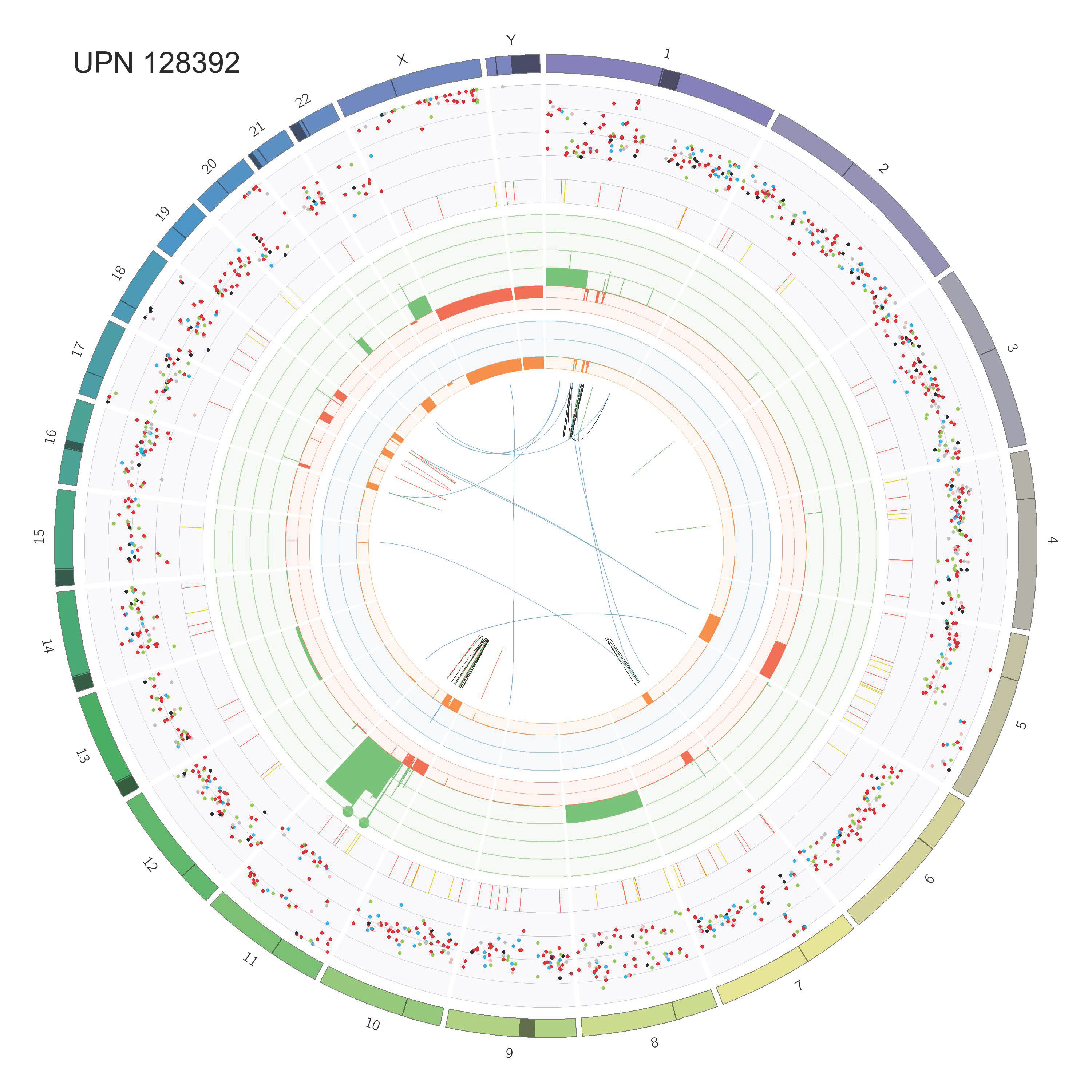

### Supp_Fig_2_circos_Page_04.jpg

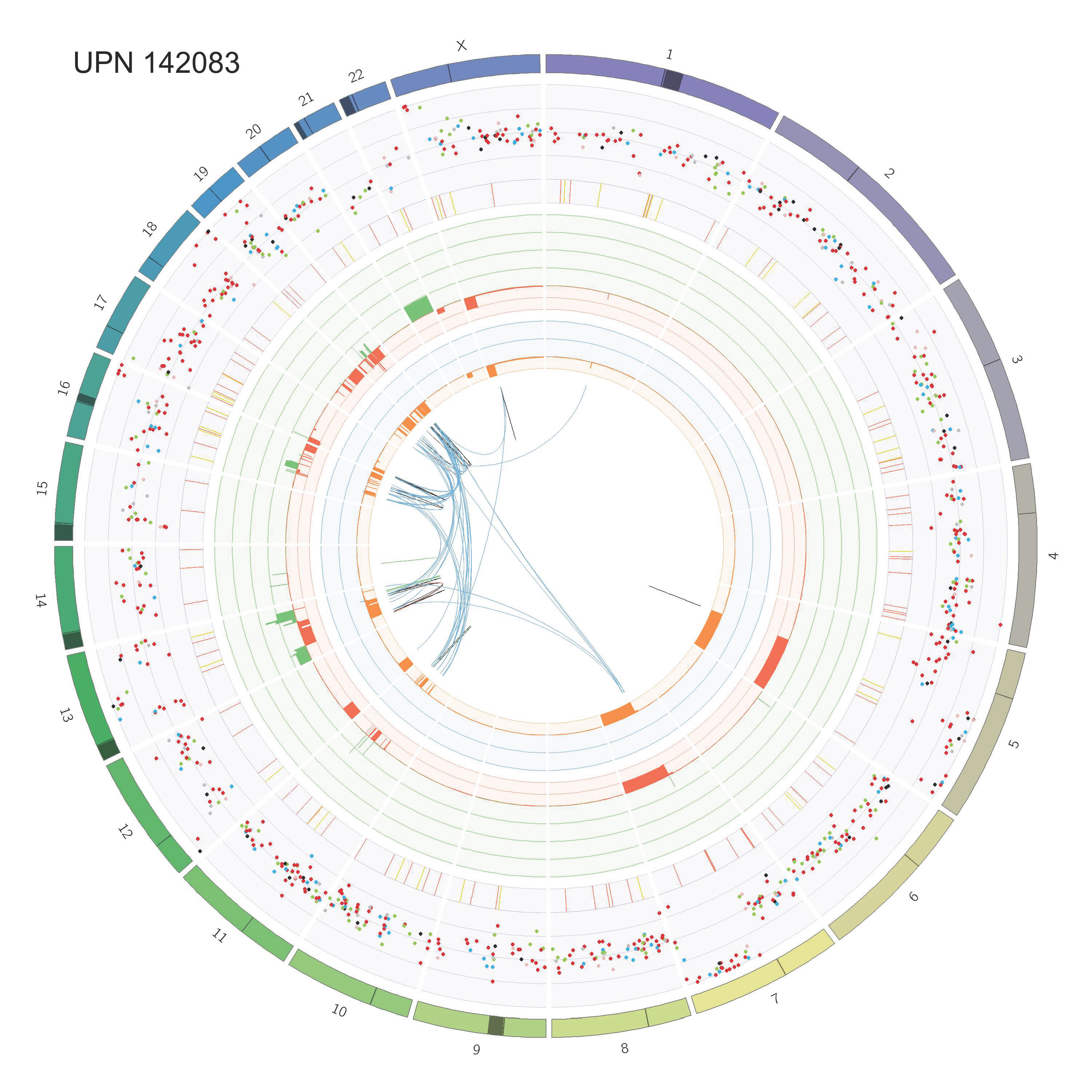

### Supp_Fig_2_circos_Page_05.jpg

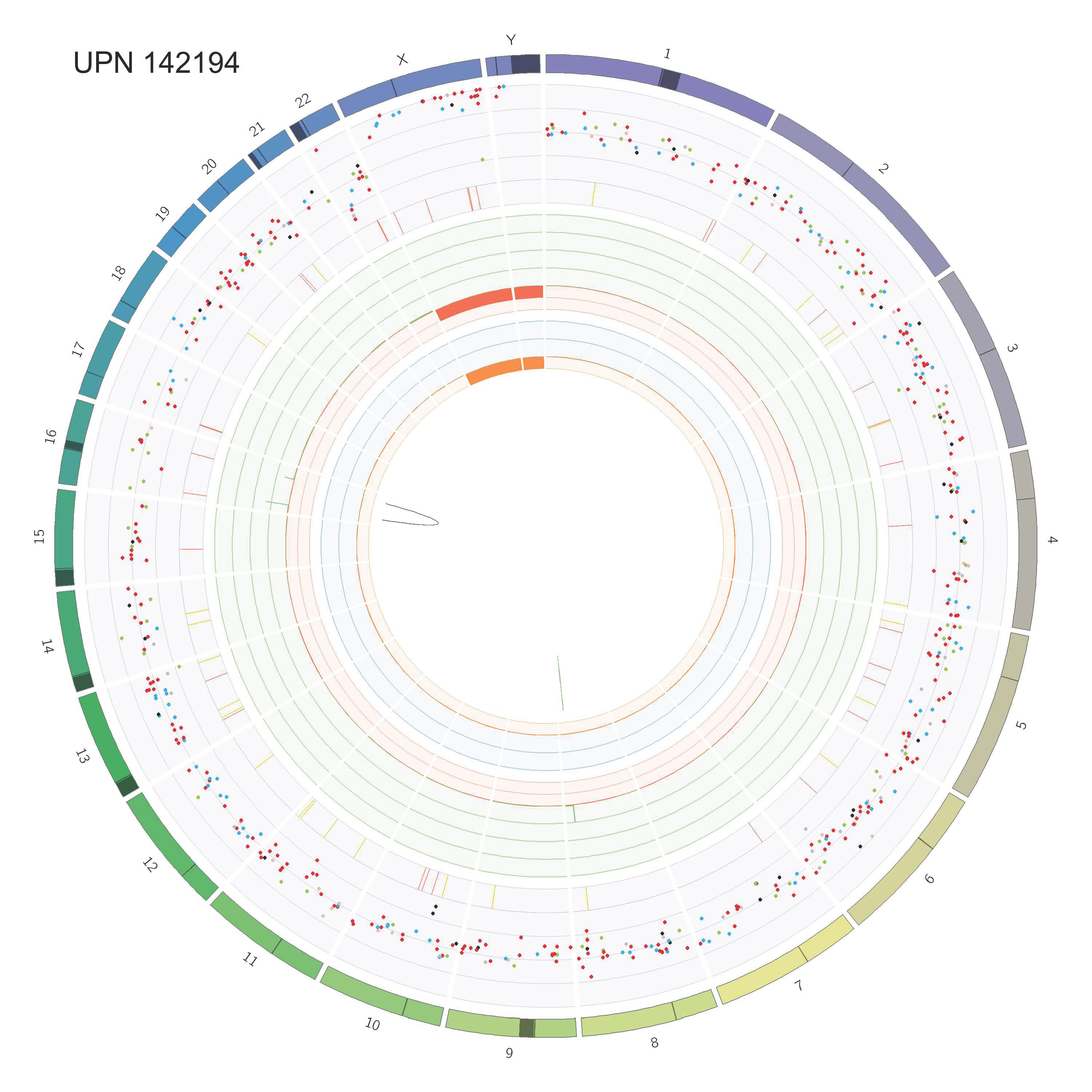

### Supp_Fig_2_circos_Page_06.jpg

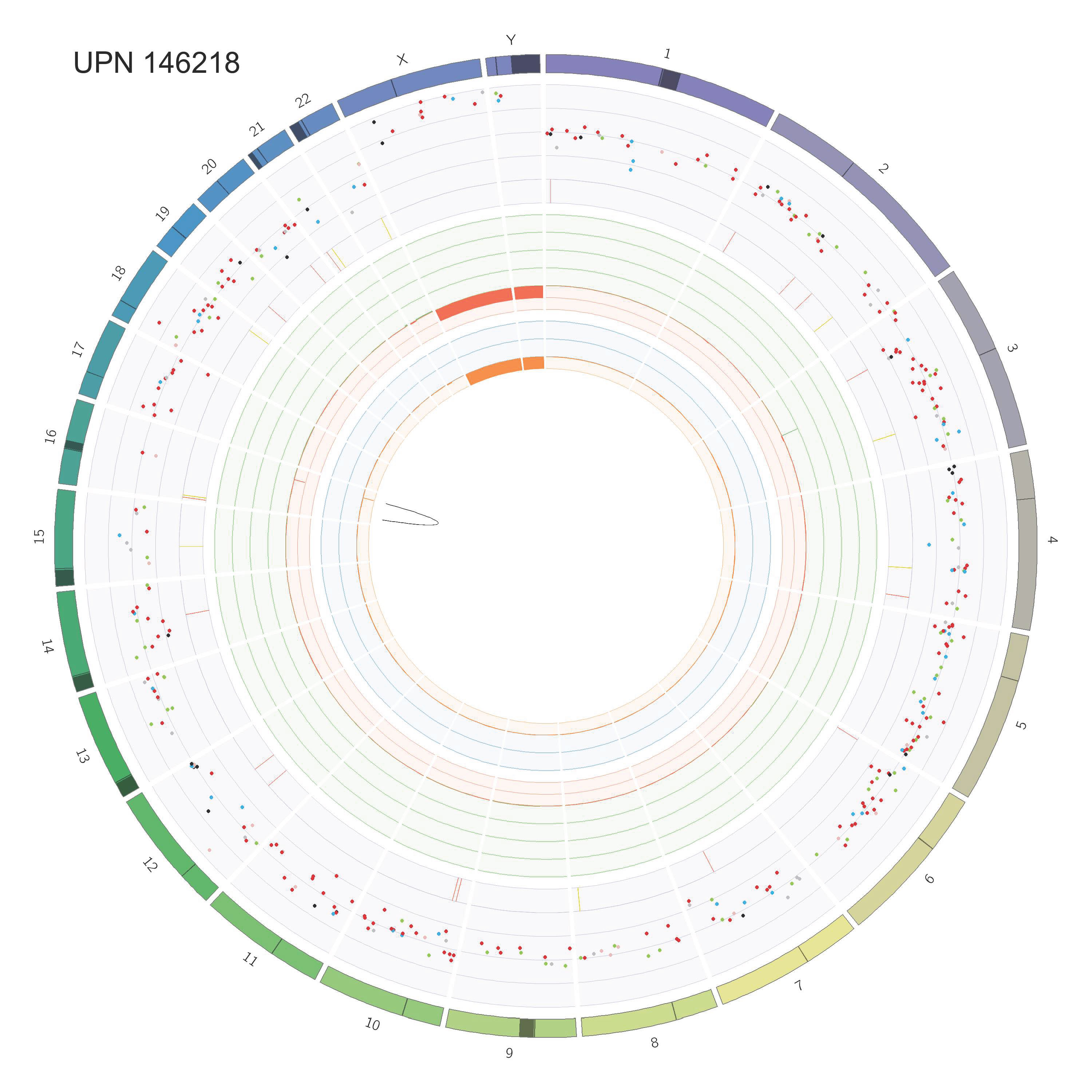

### Supp_Fig_2_circos_Page_07.jpg

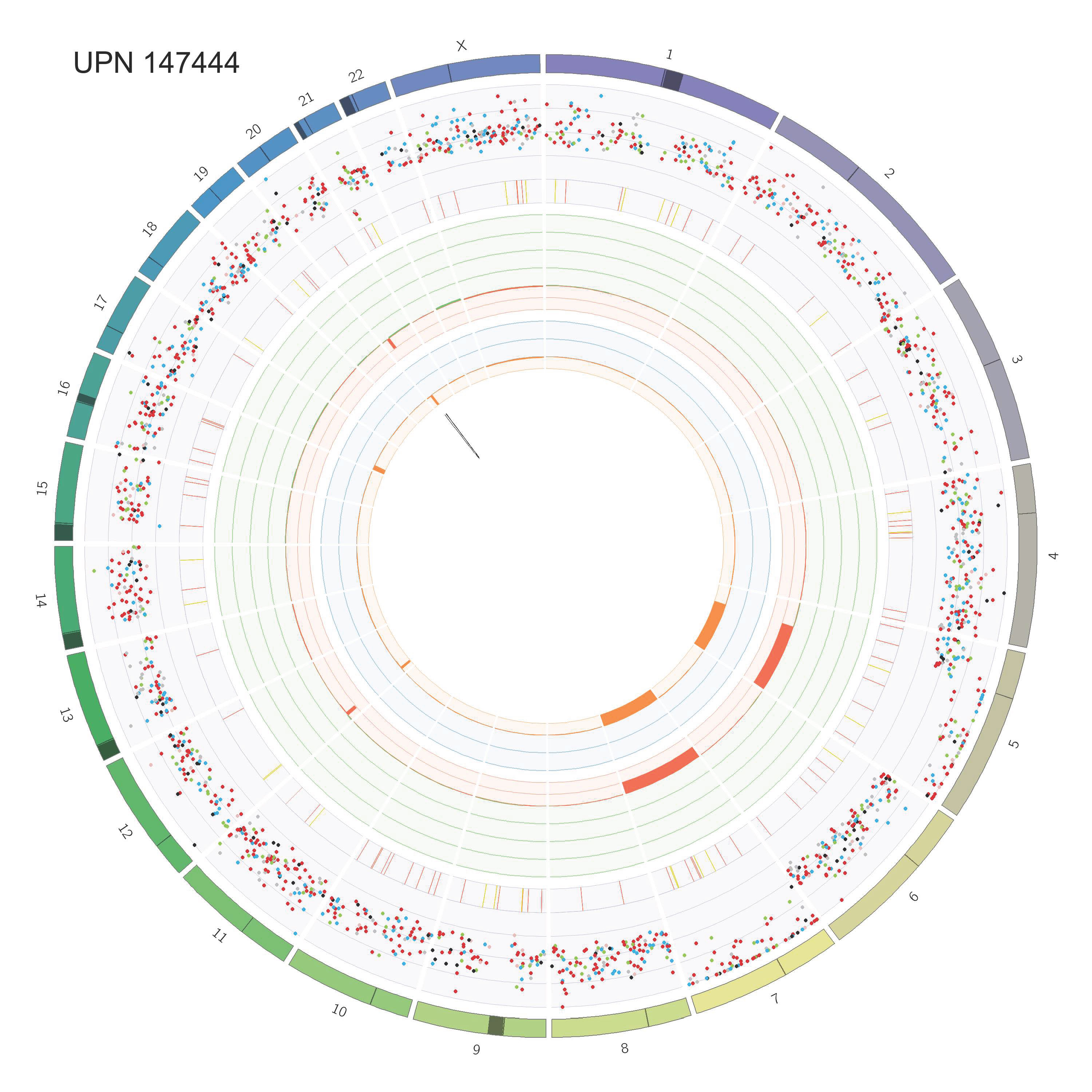

### Supp_Fig_2_circos_Page_08.jpg

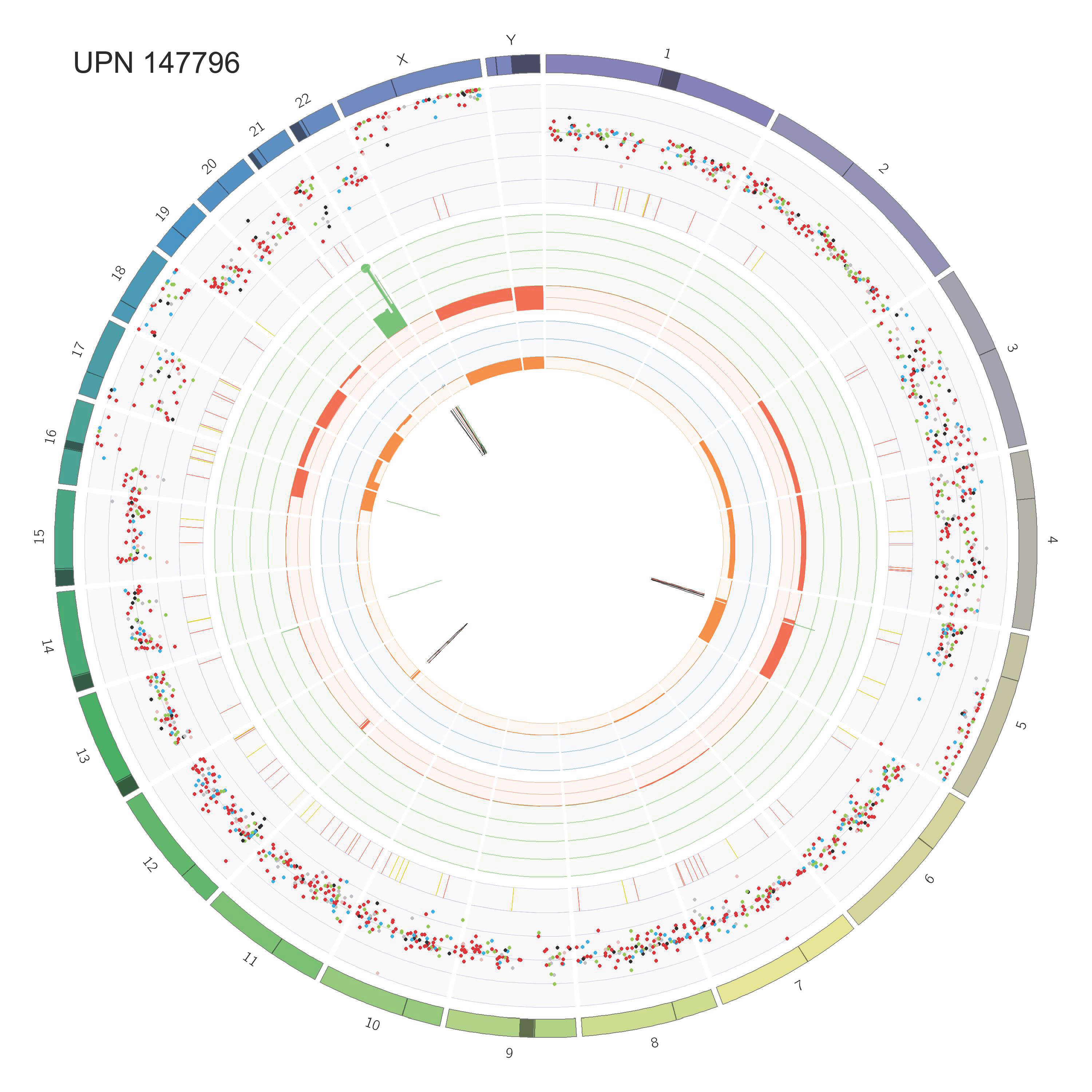

### Supp_Fig_2_circos_Page_09.jpg

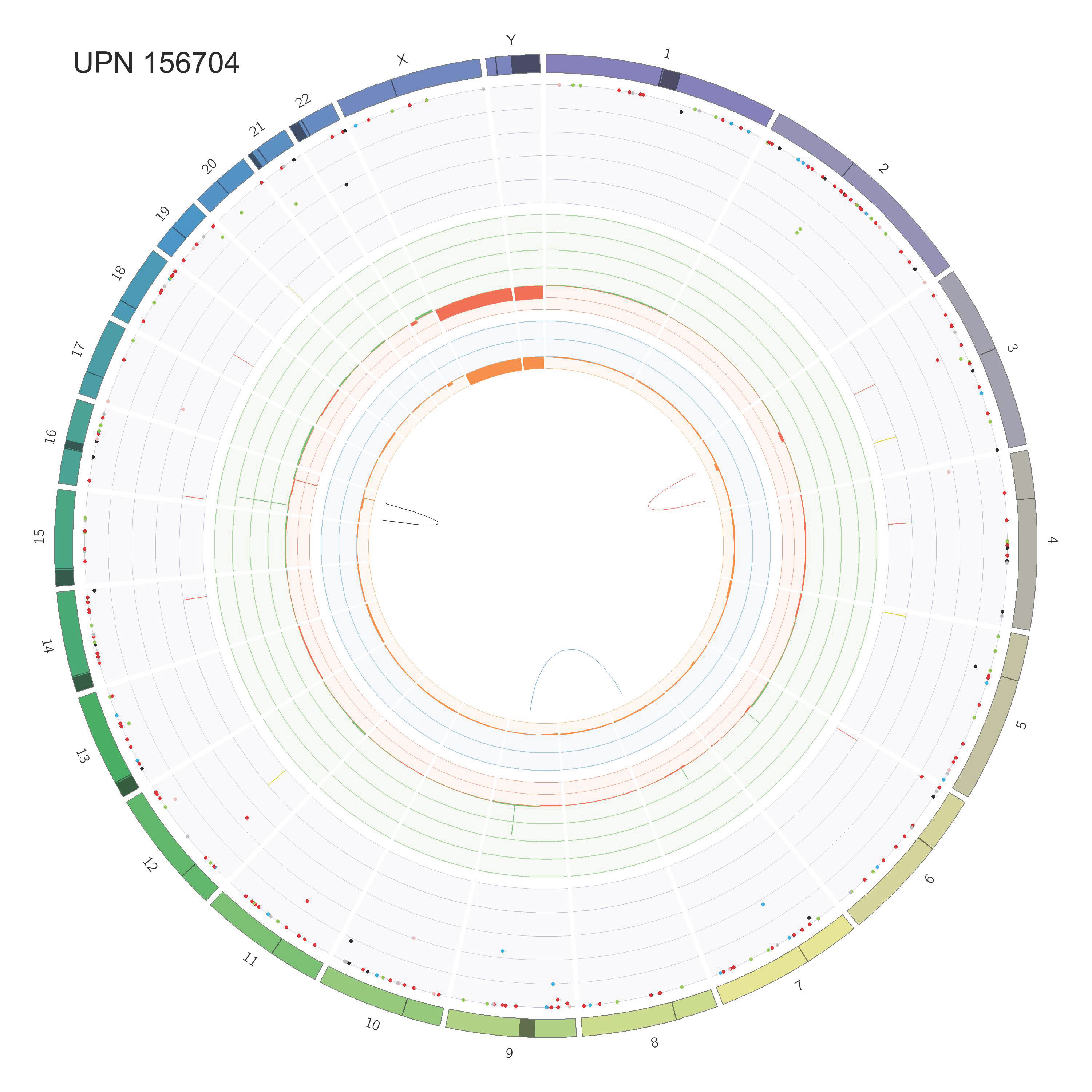

### Supp_Fig_2_circos_Page_10.jpg

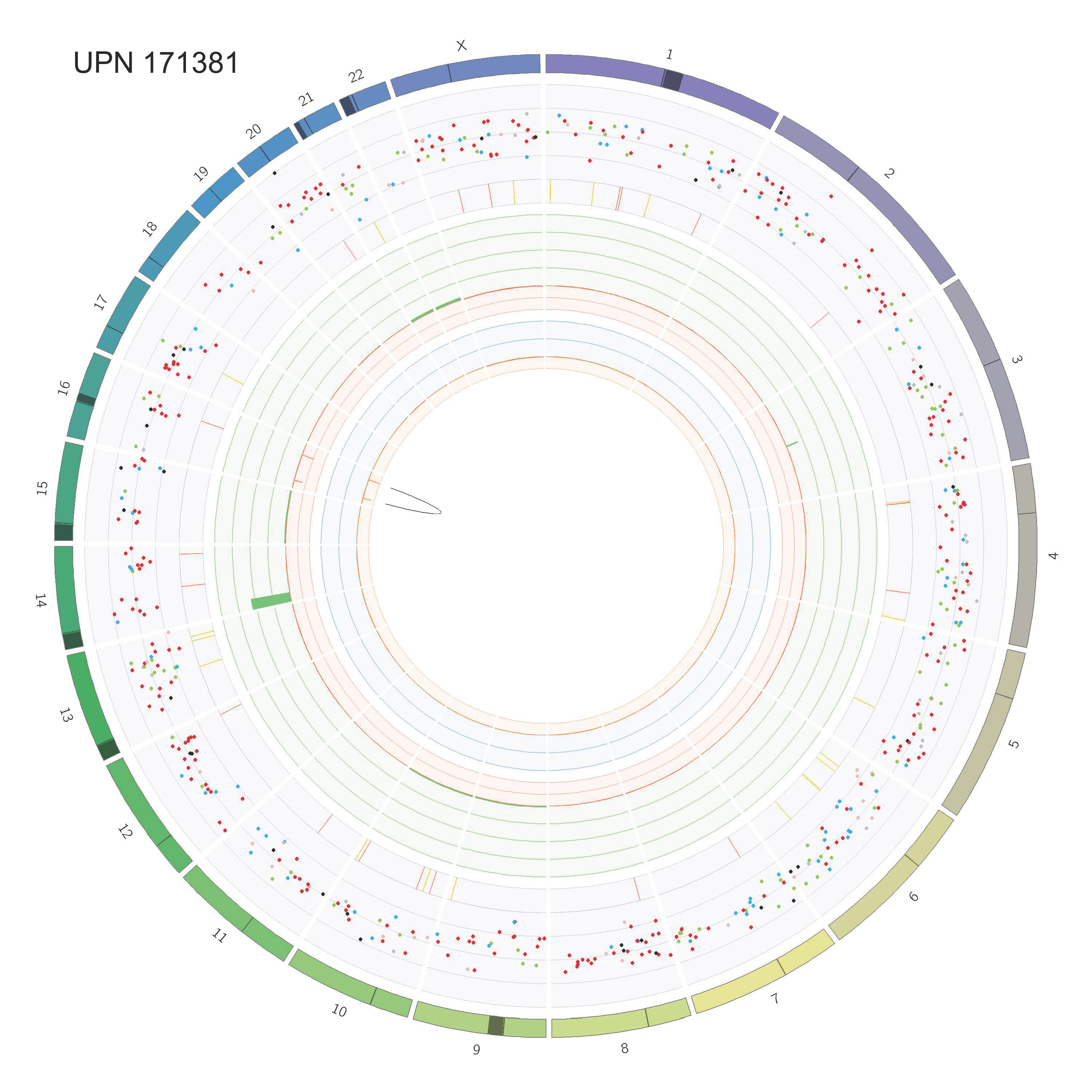

### Supp_Fig_2_circos_Page_11.jpg

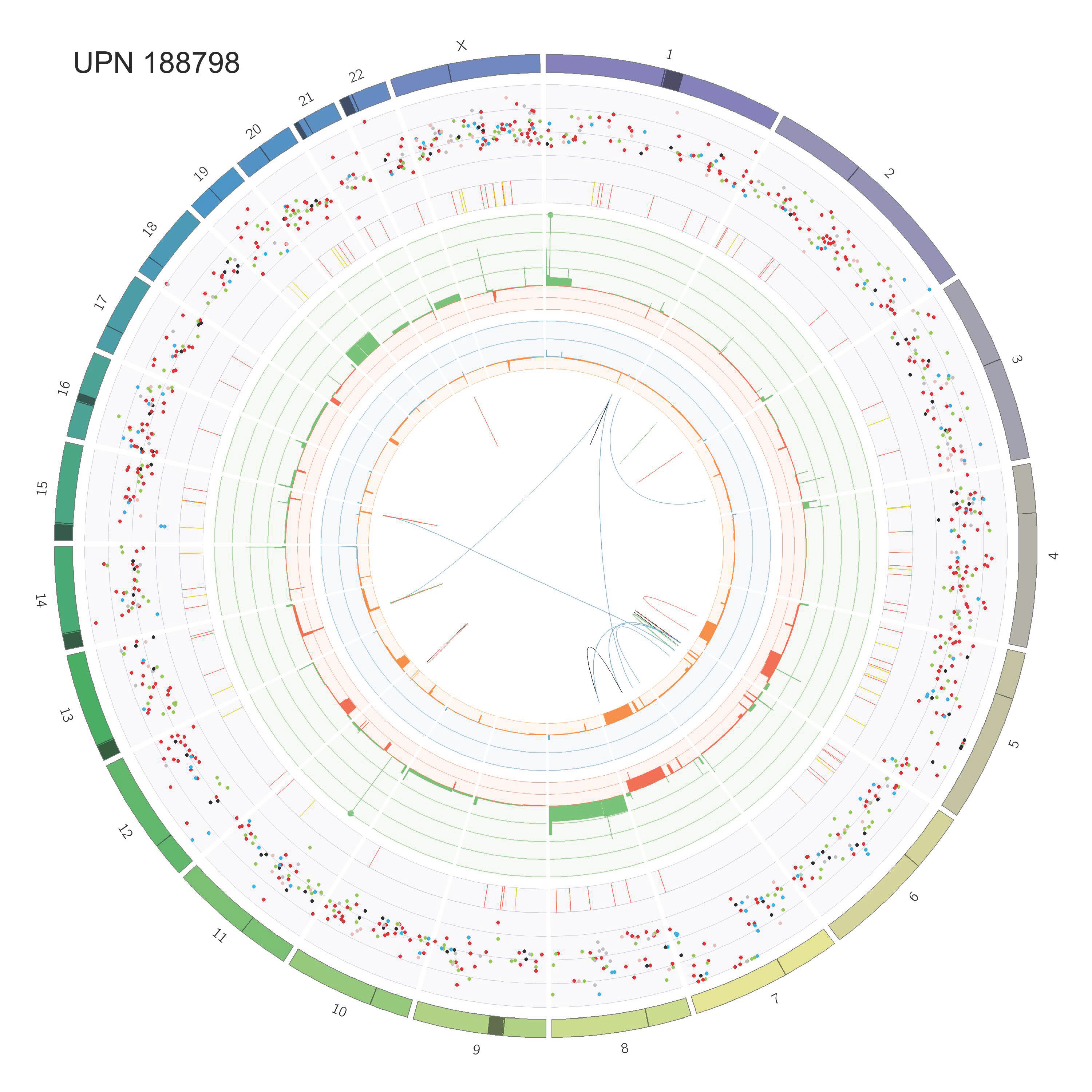

### Supp_Fig_2_circos_Page_12.jpg

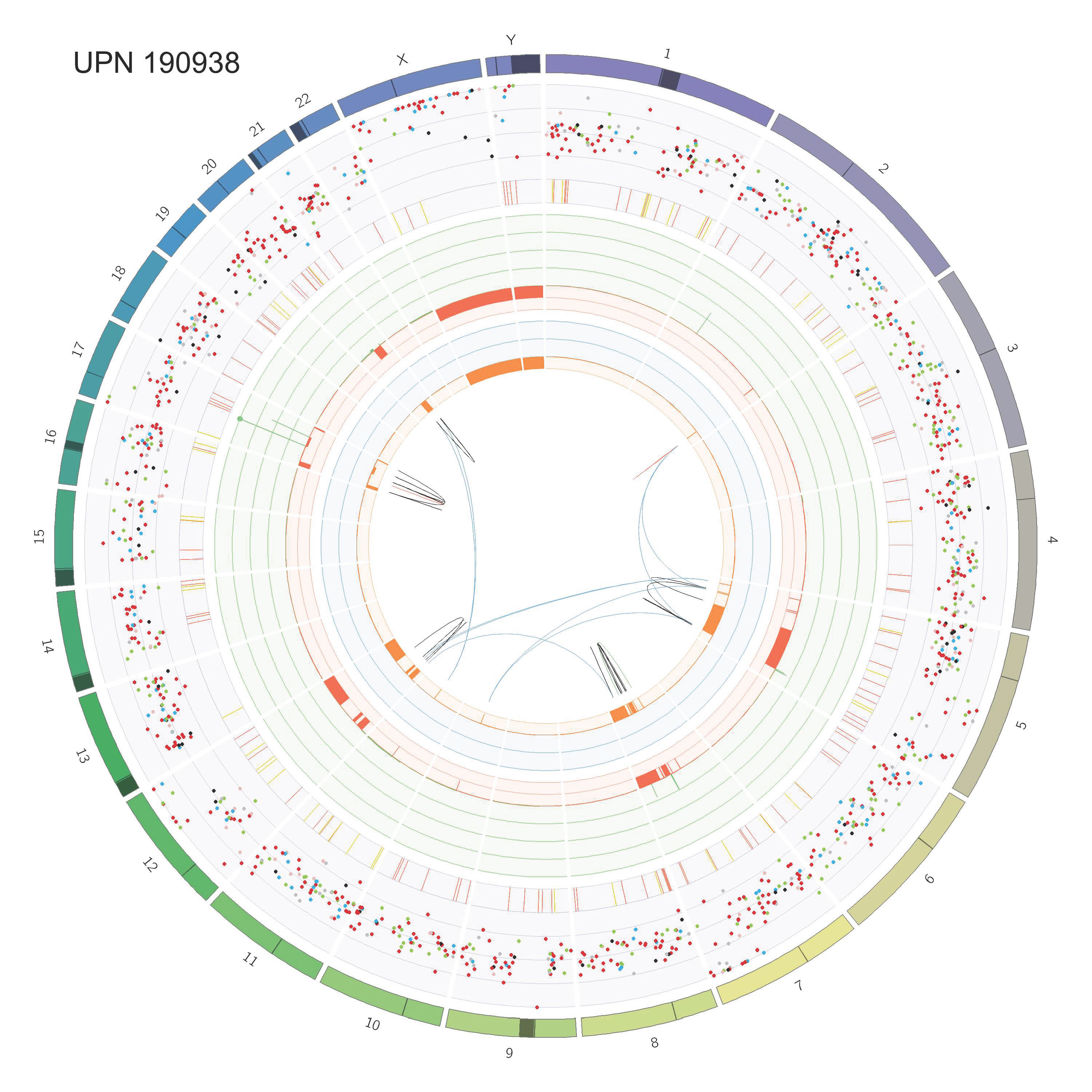

### Supp_Fig_2_circos_Page_13.jpg

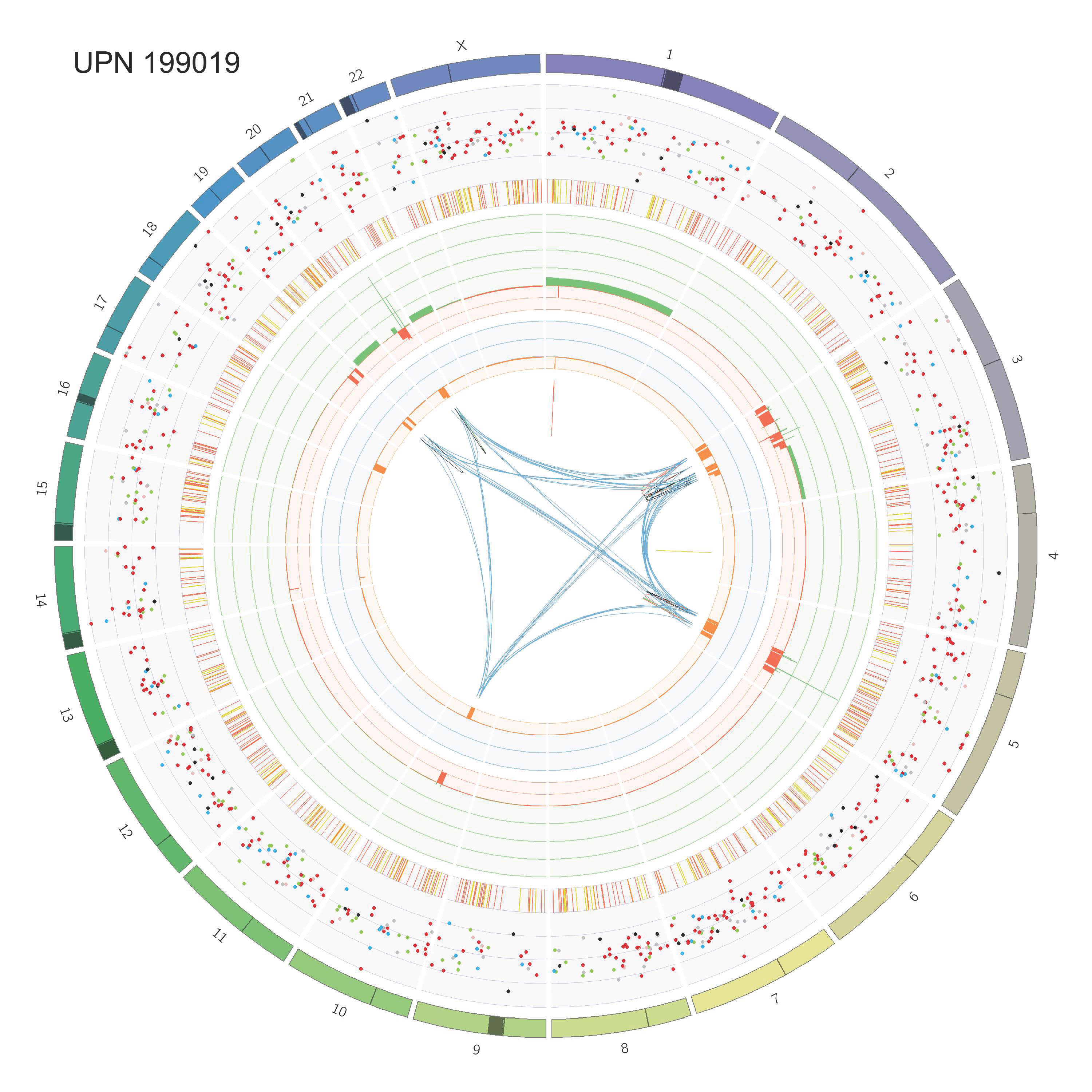

### Supp_Fig_2_circos_Page_14.jpg

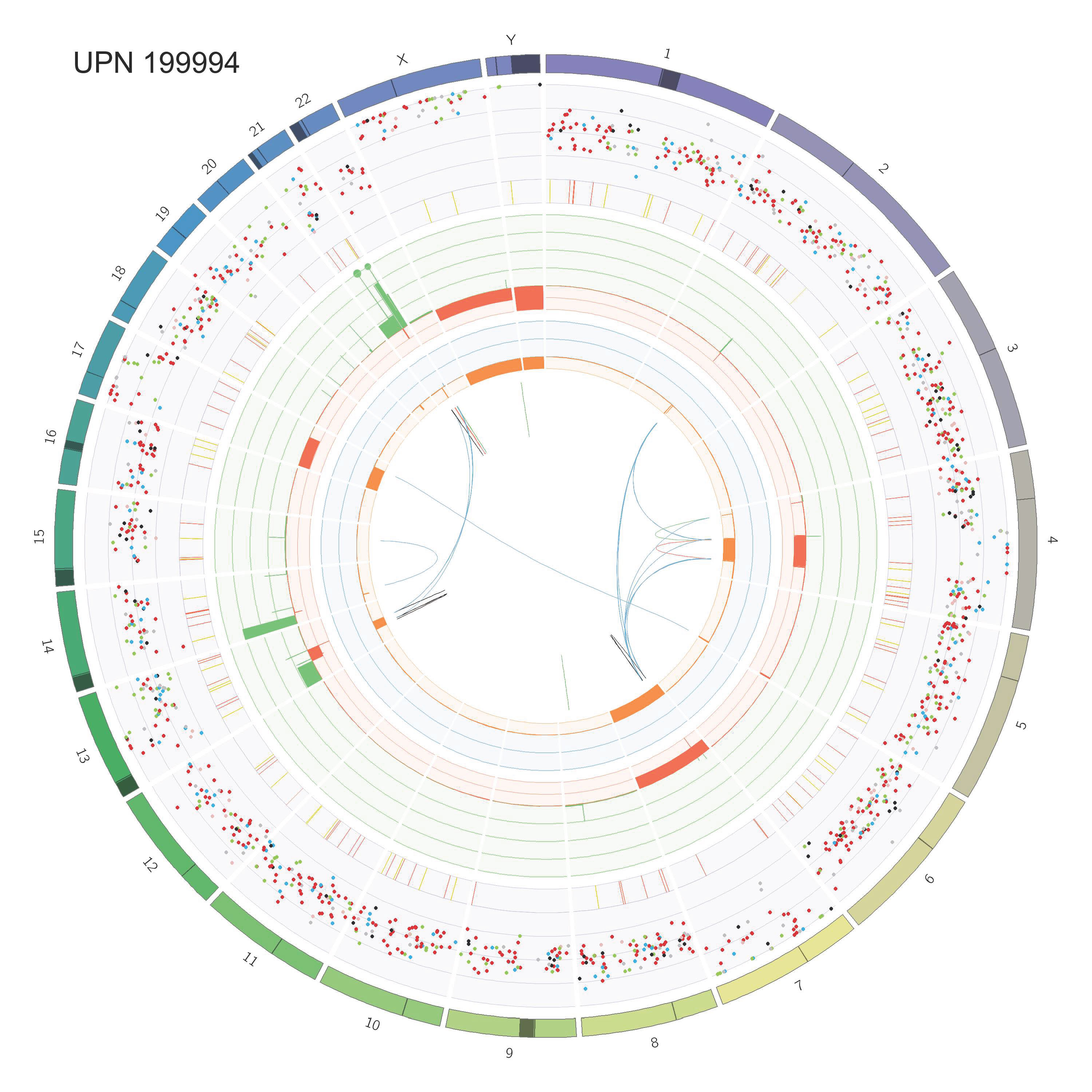

### Supp_Fig_2_circos_Page_15.jpg

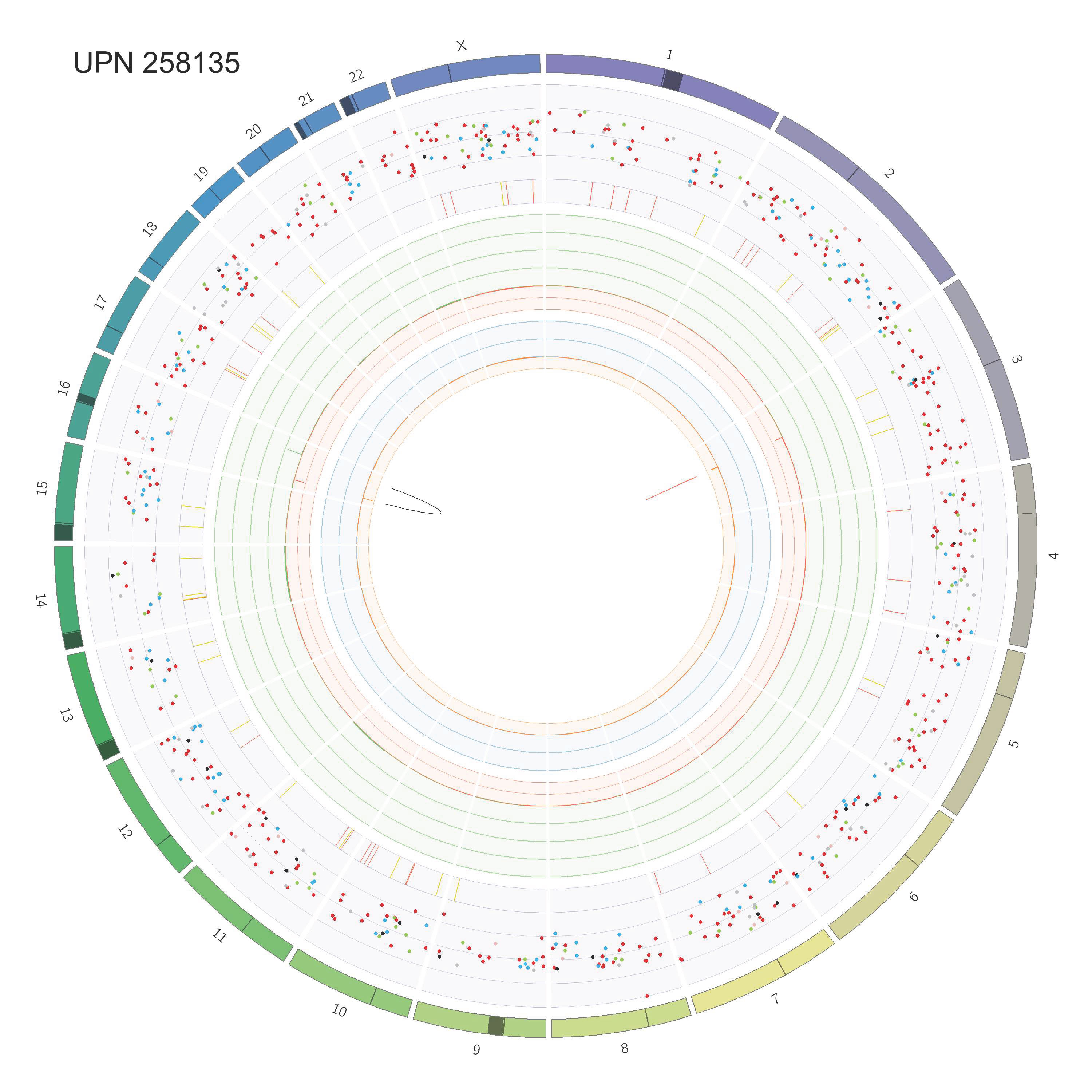

### Supp_Fig_2_circos_Page_16.jpg

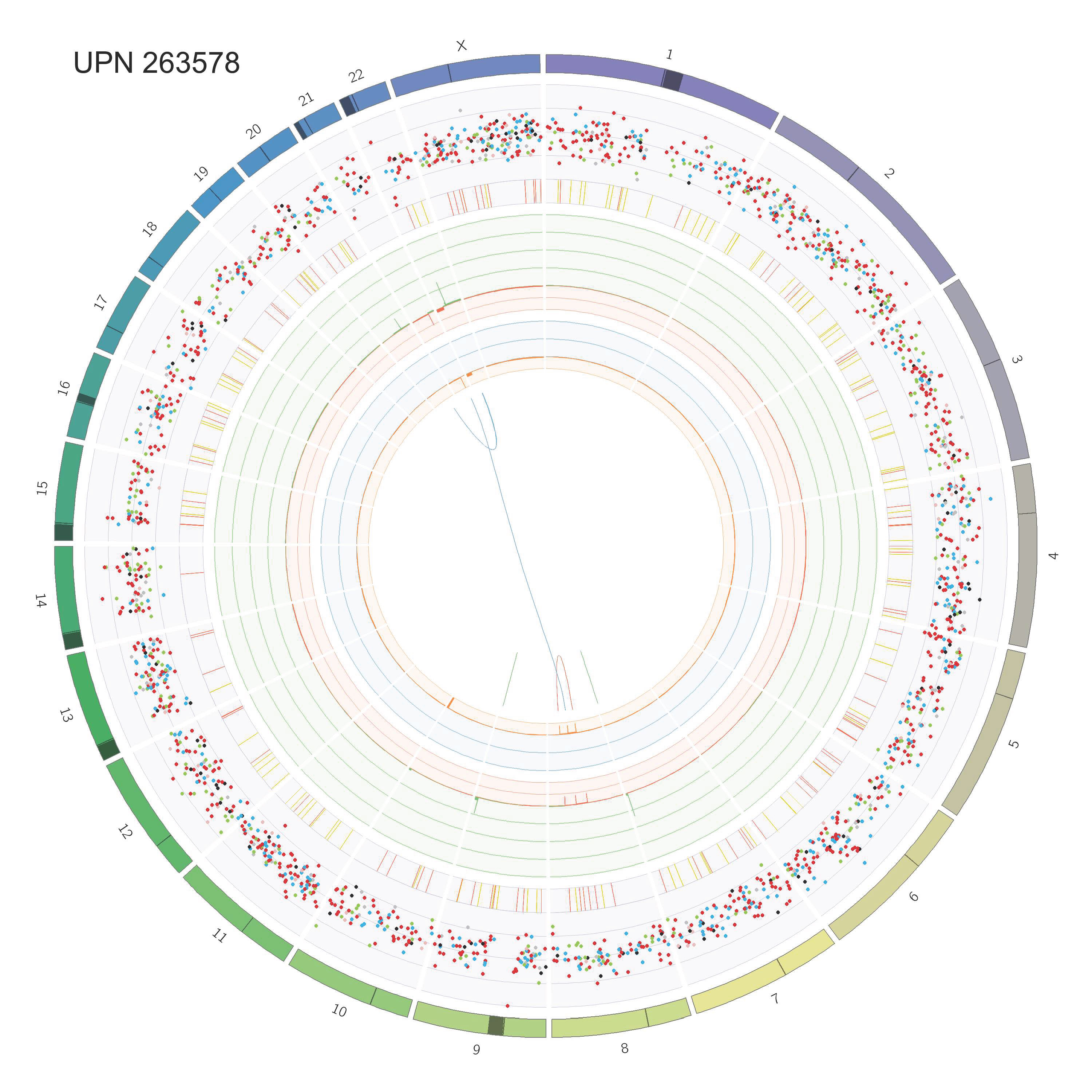

### Supp_Fig_2_circos_Page_17.jpg

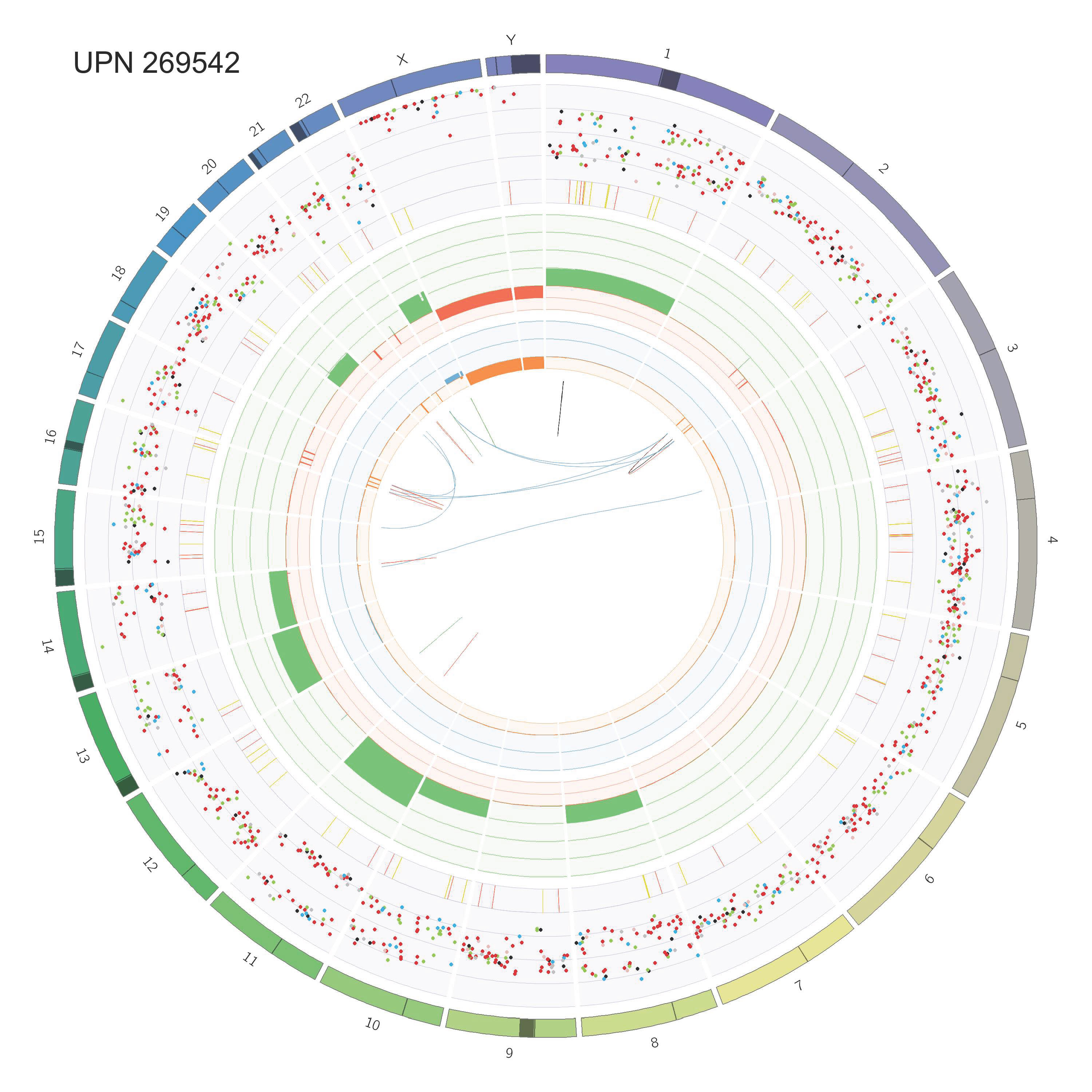

### Supp_Fig_2_circos_Page_18.jpg

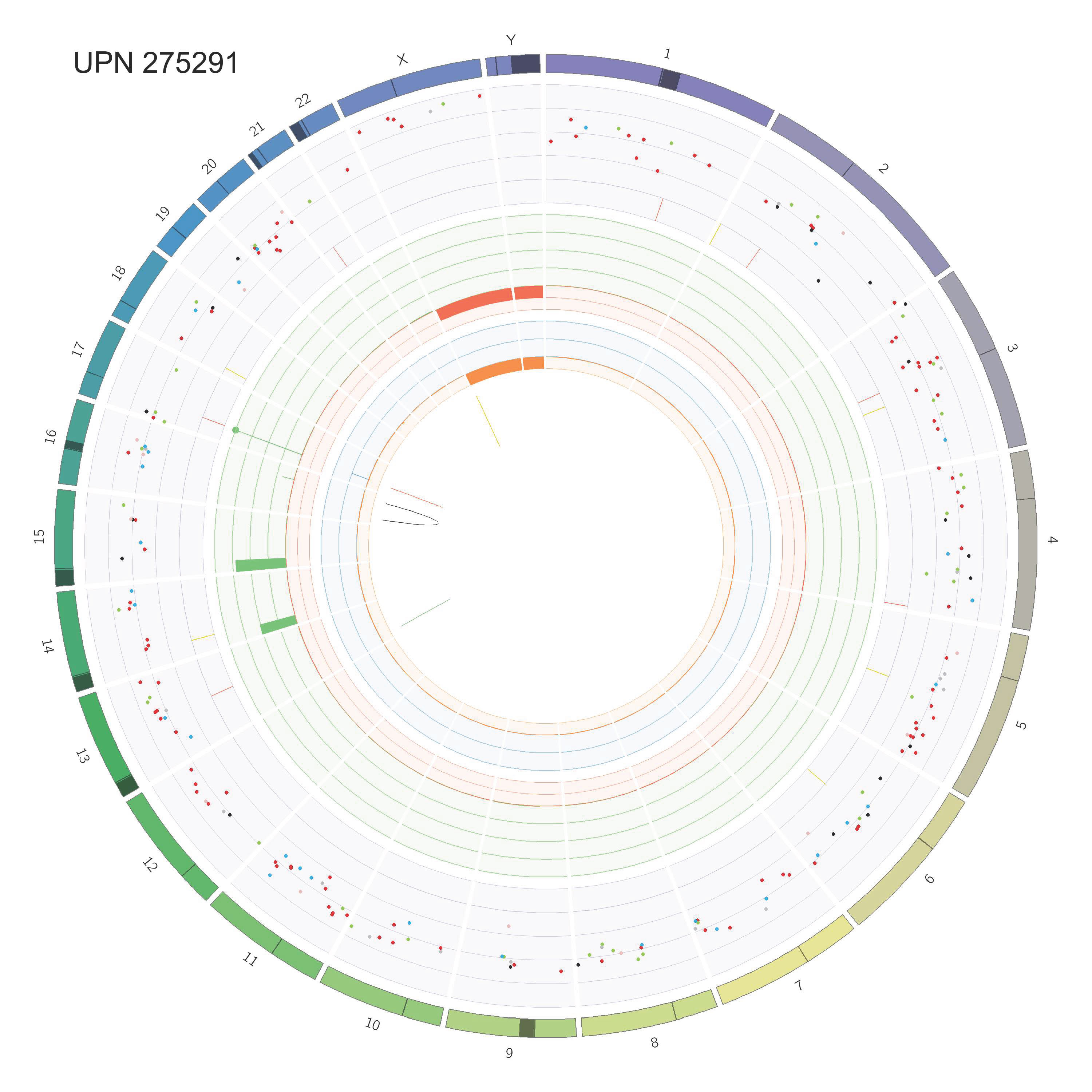

### Supp_Fig_2_circos_Page_19.jpg

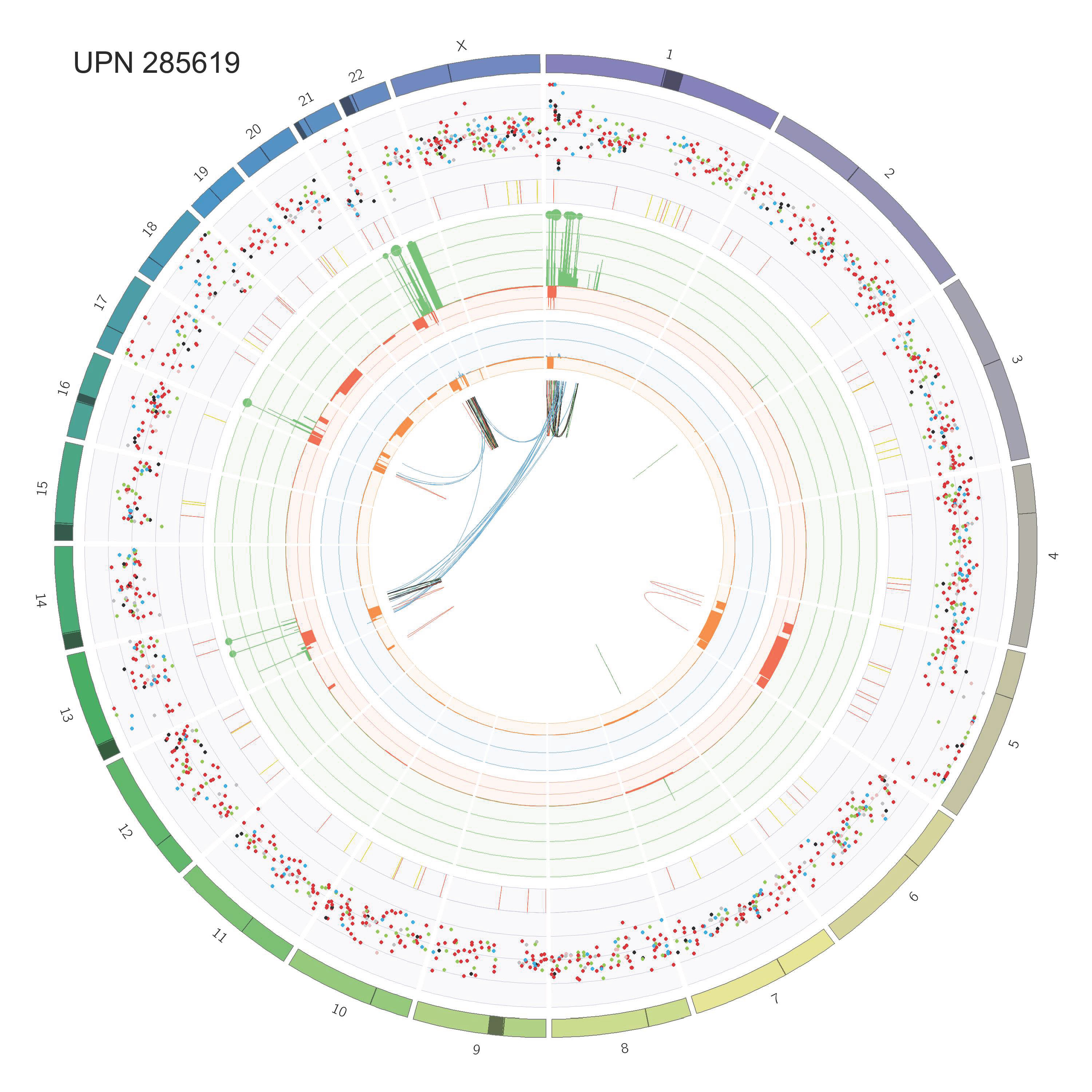

### Supp_Fig_2_circos_Page_20.jpg

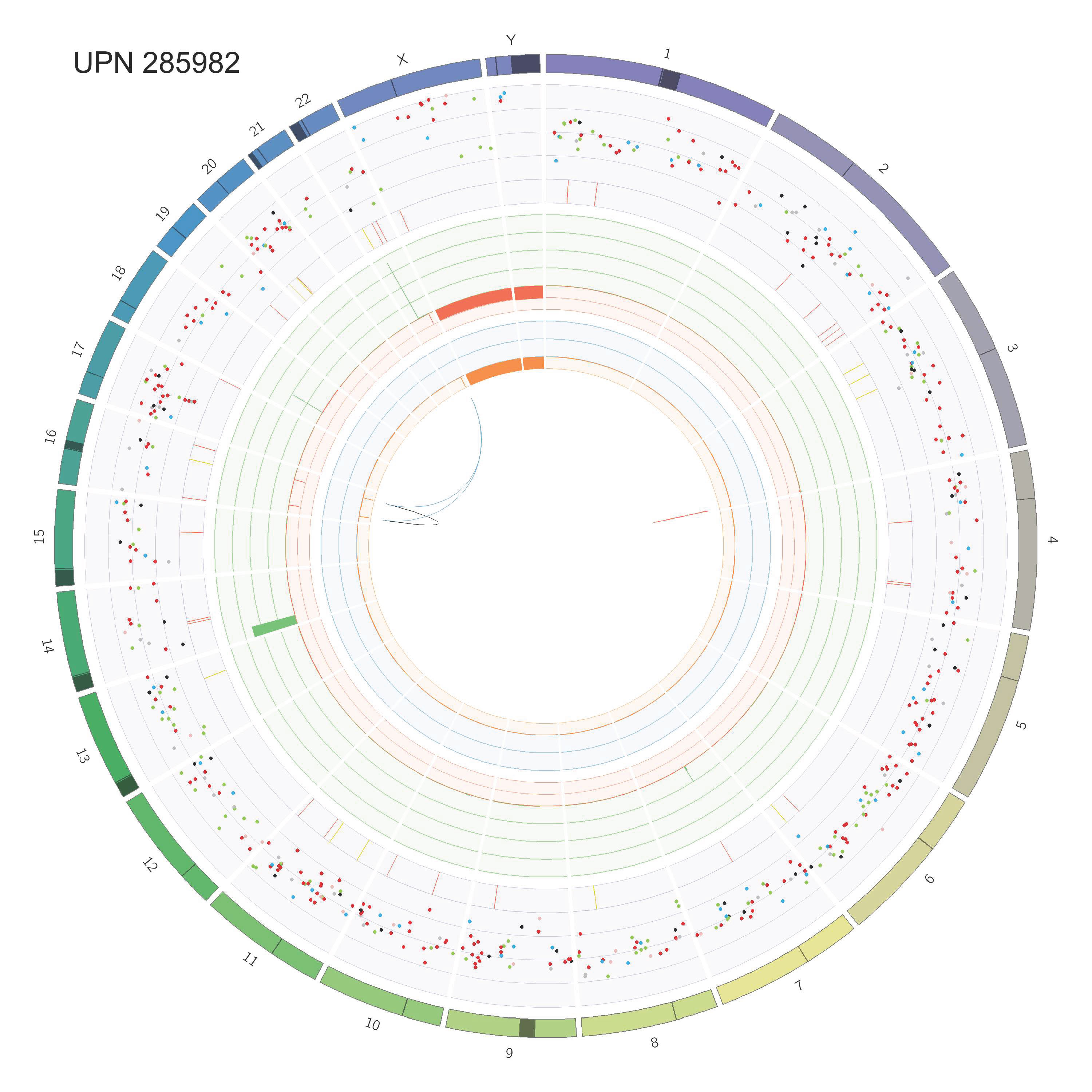

### Supp_Fig_2_circos_Page_21.jpg

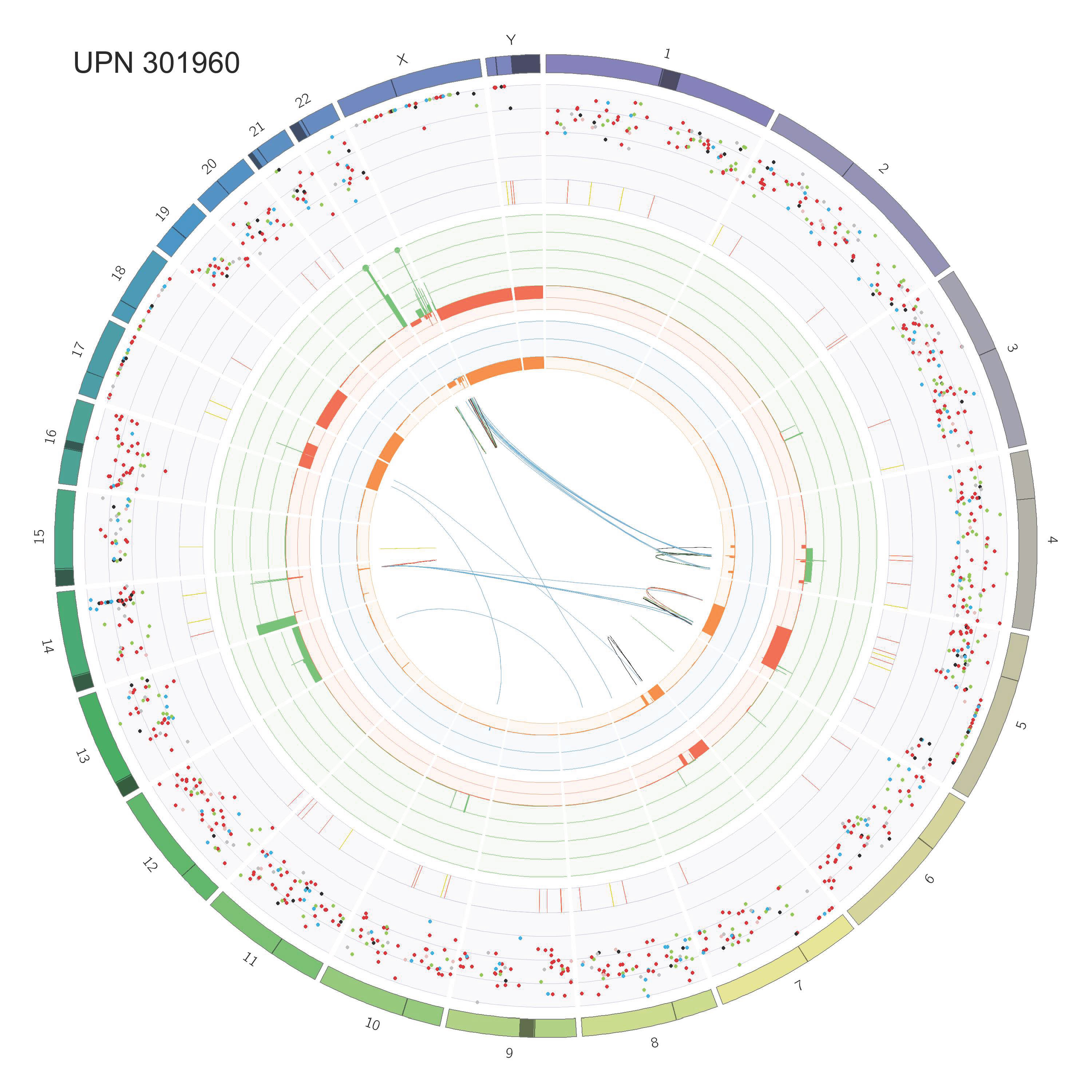

### Supp_Fig_2_circos_Page_22.jpg

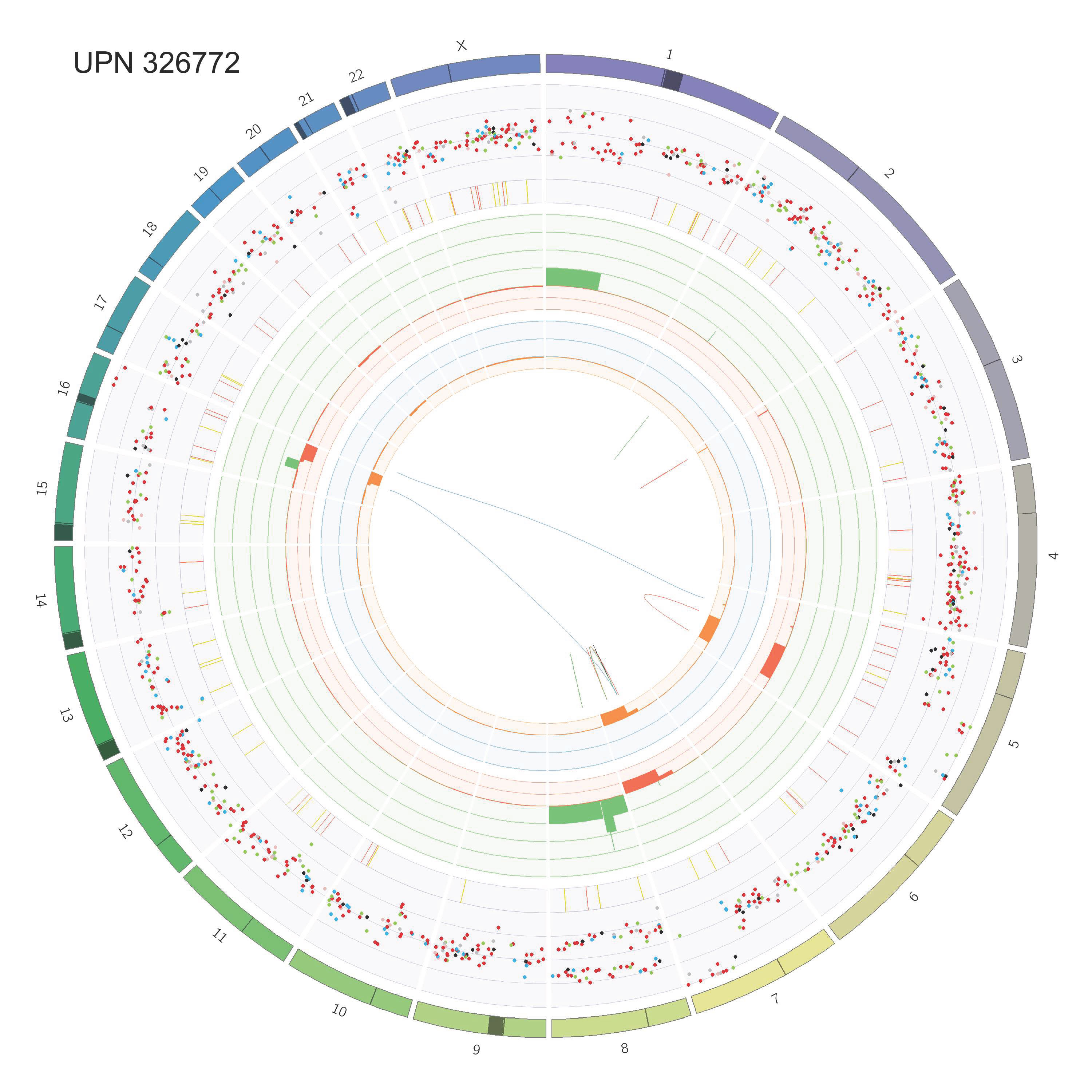

### Supp_Fig_2_circos_Page_23.jpg

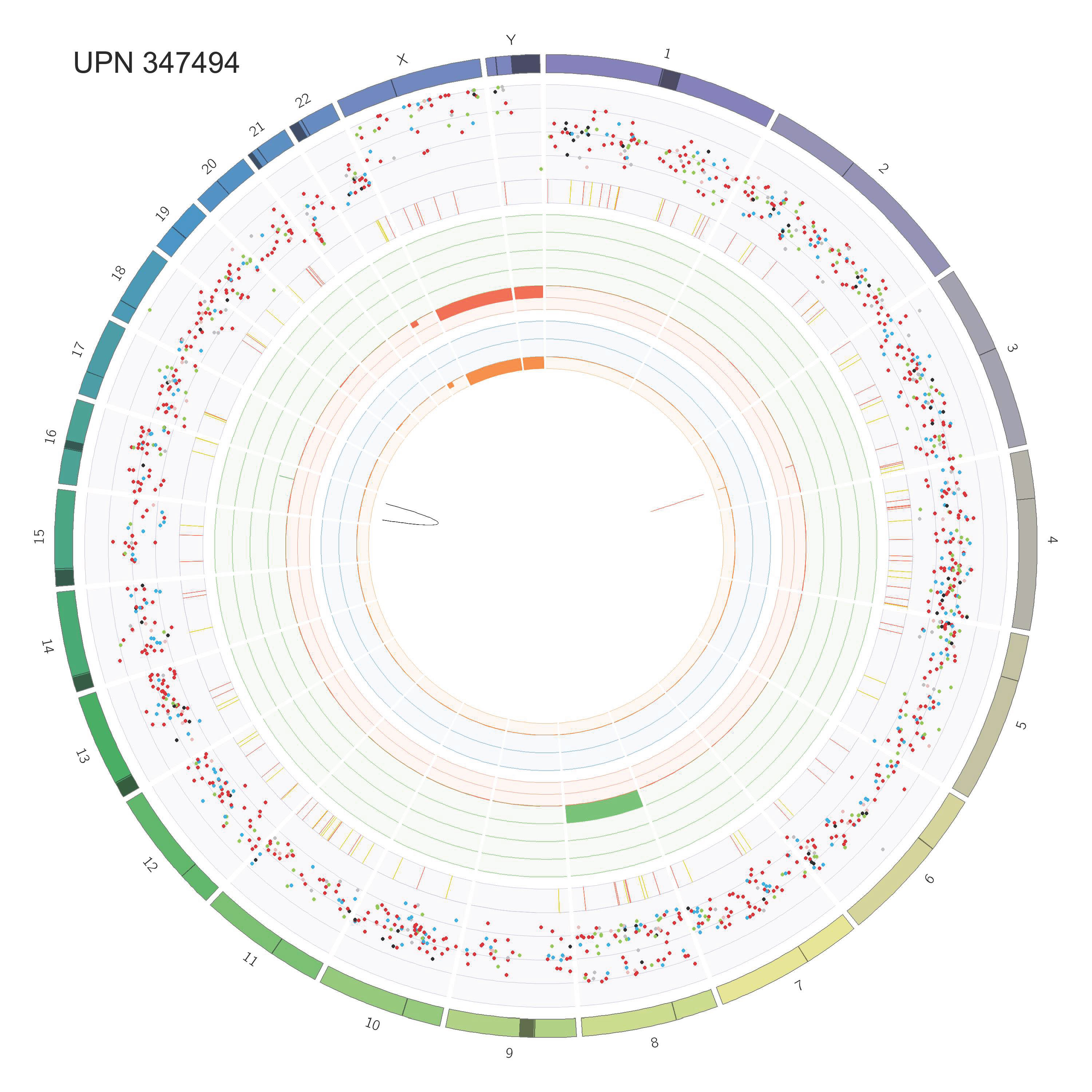

### Supp_Fig_2_circos_Page_24.jpg

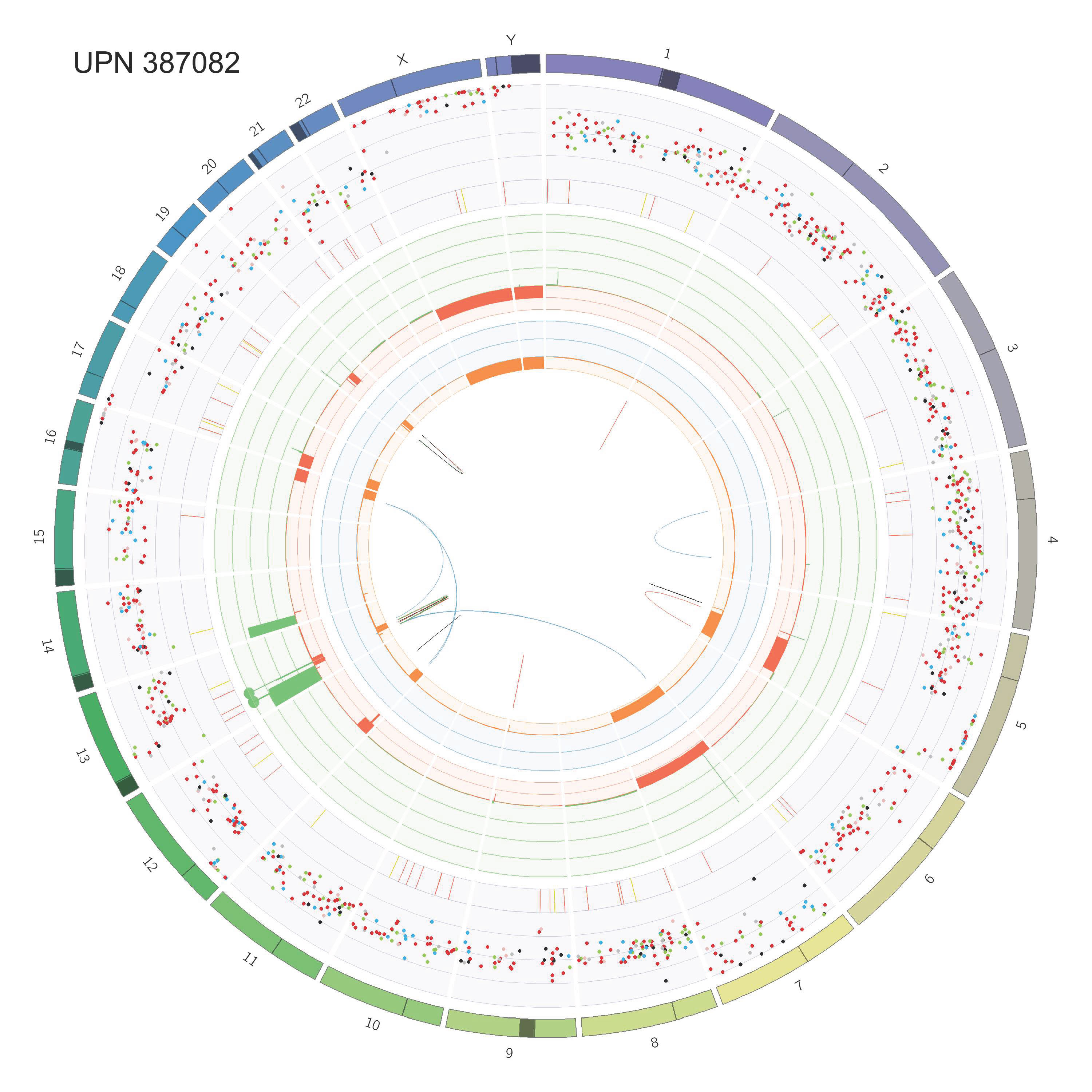

### Supp_Fig_2_circos_Page_25.jpg

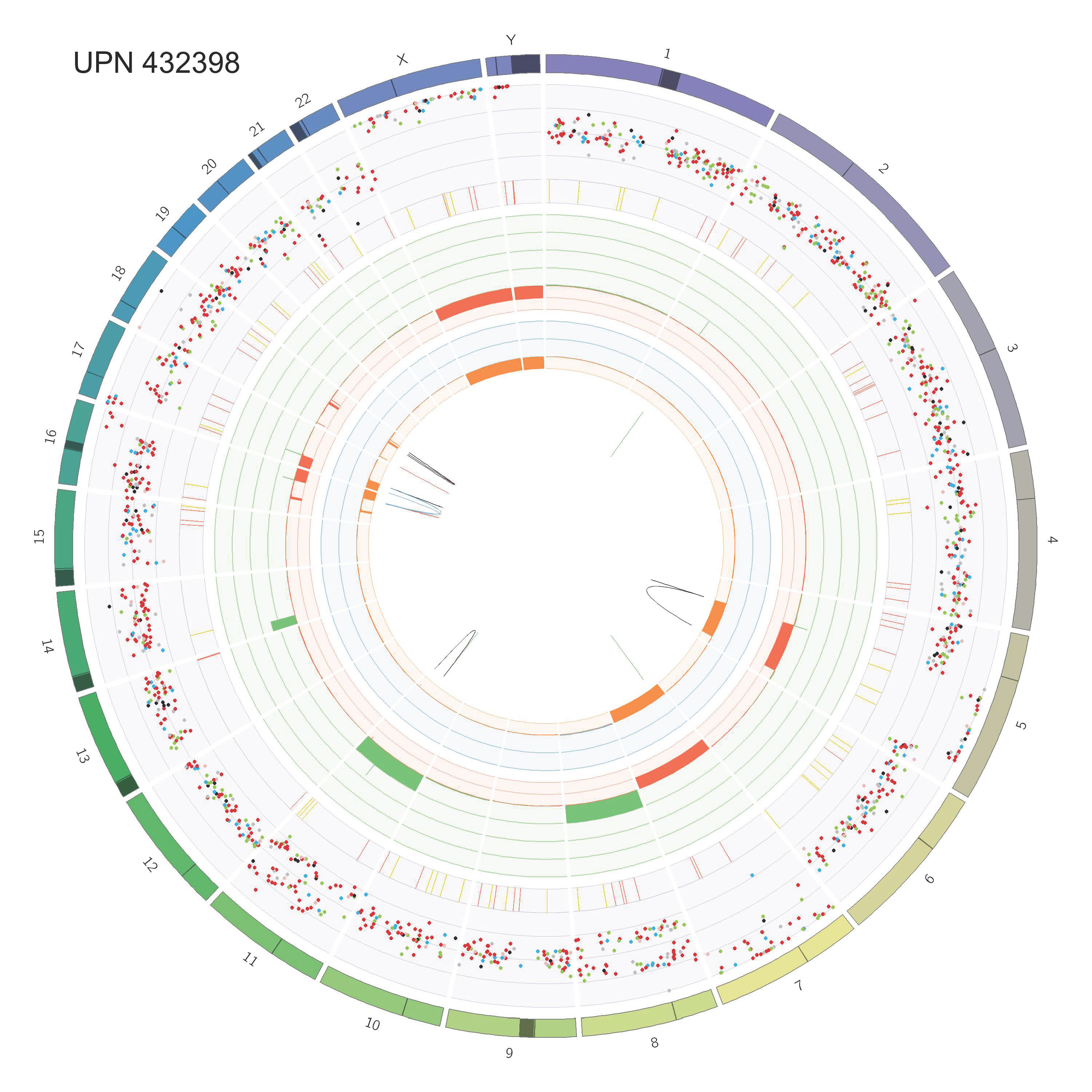

### Supp_Fig_2_circos_Page_26.jpg

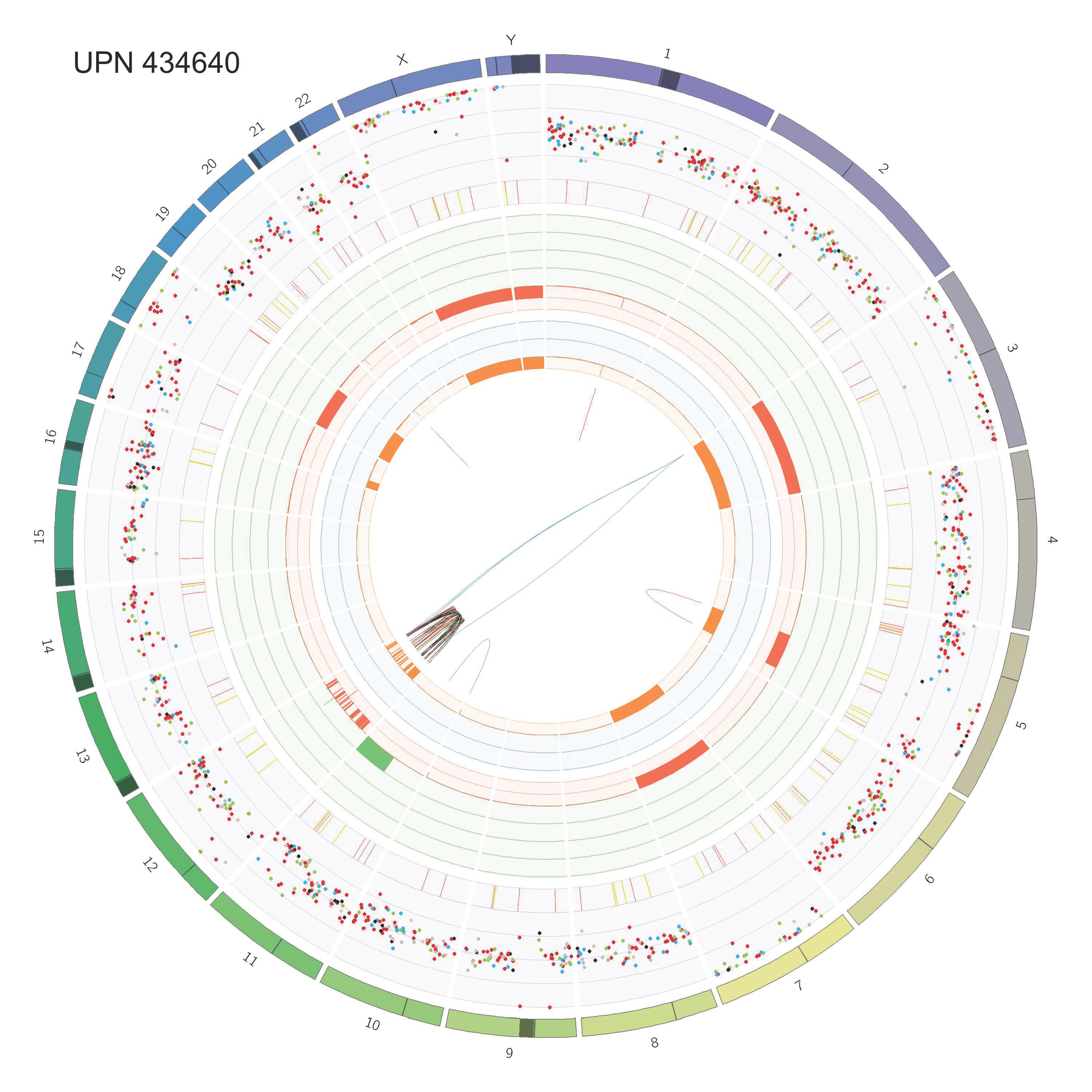

### Supp_Fig_2_circos_Page_27.jpg

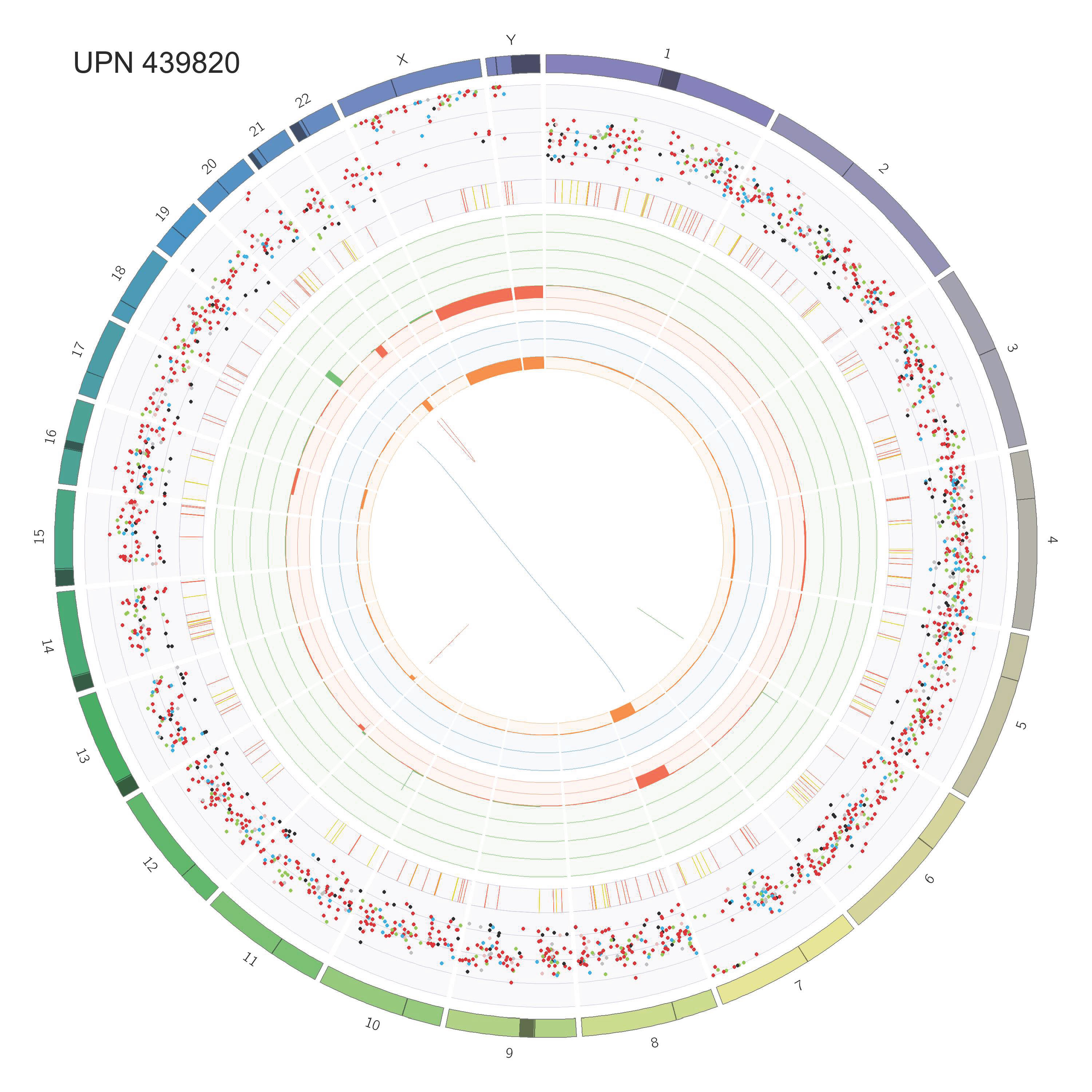

### Supp_Fig_2_circos_Page_28.jpg

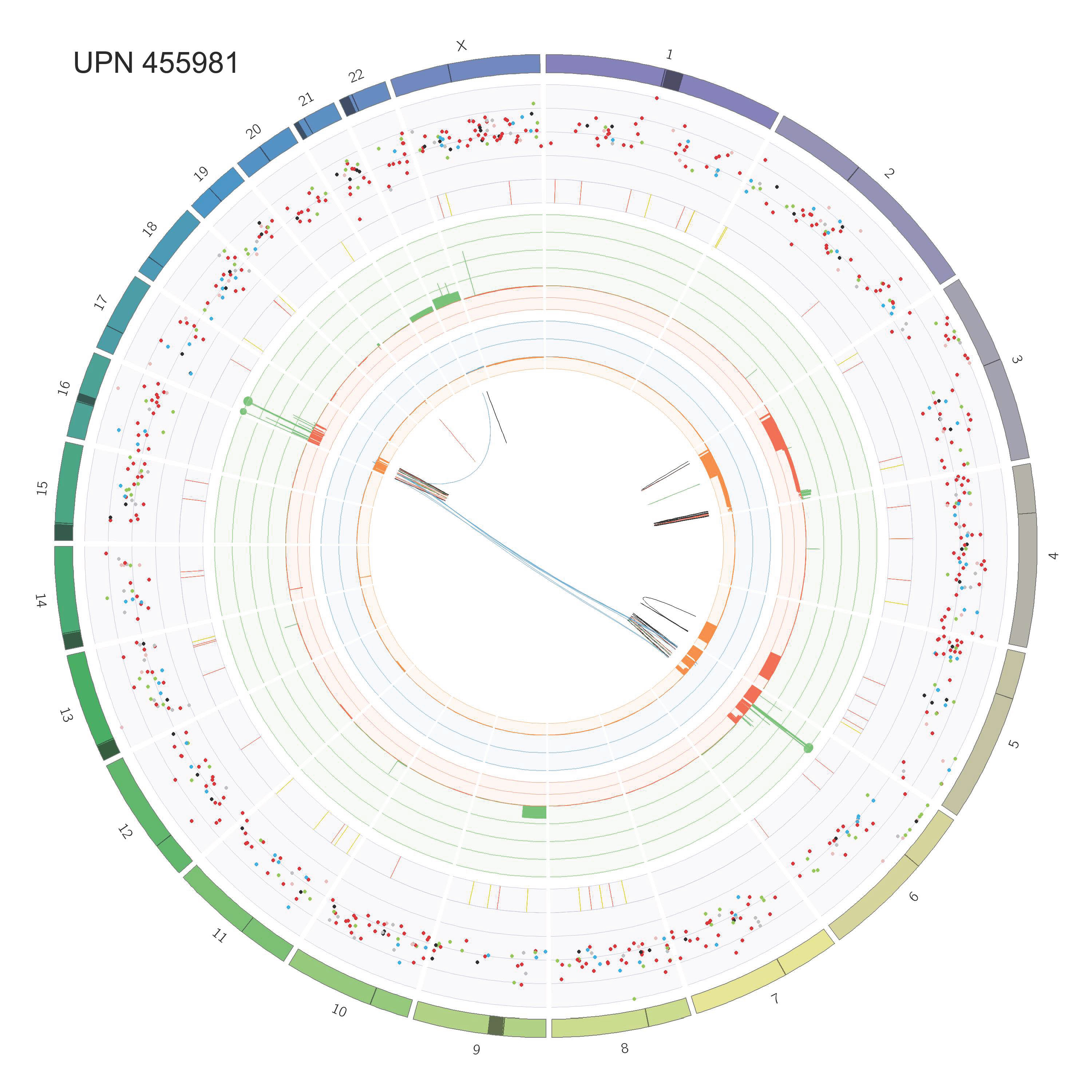

### Supp_Fig_2_circos_Page_29.jpg

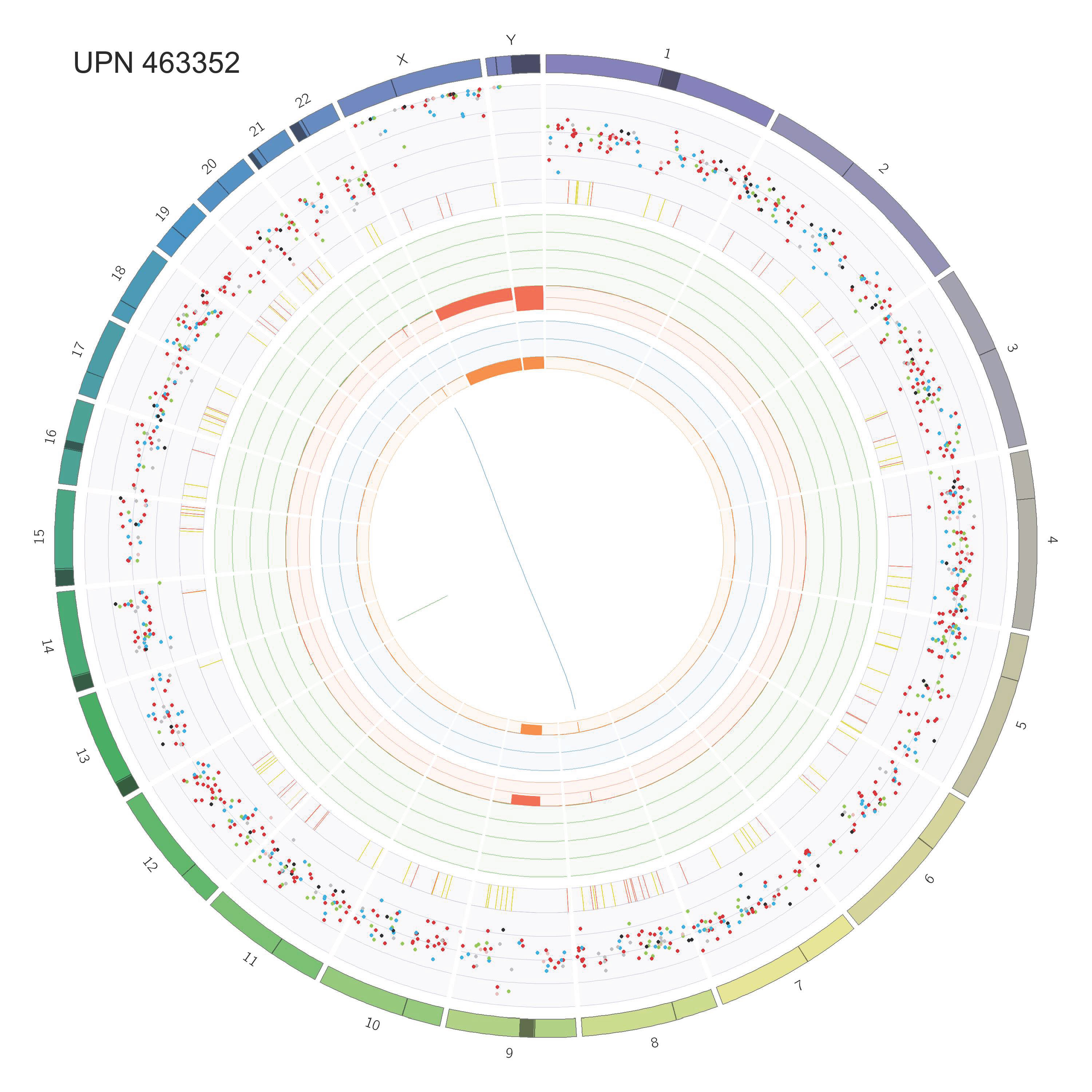

### Supp_Fig_2_circos_Page_30.jpg

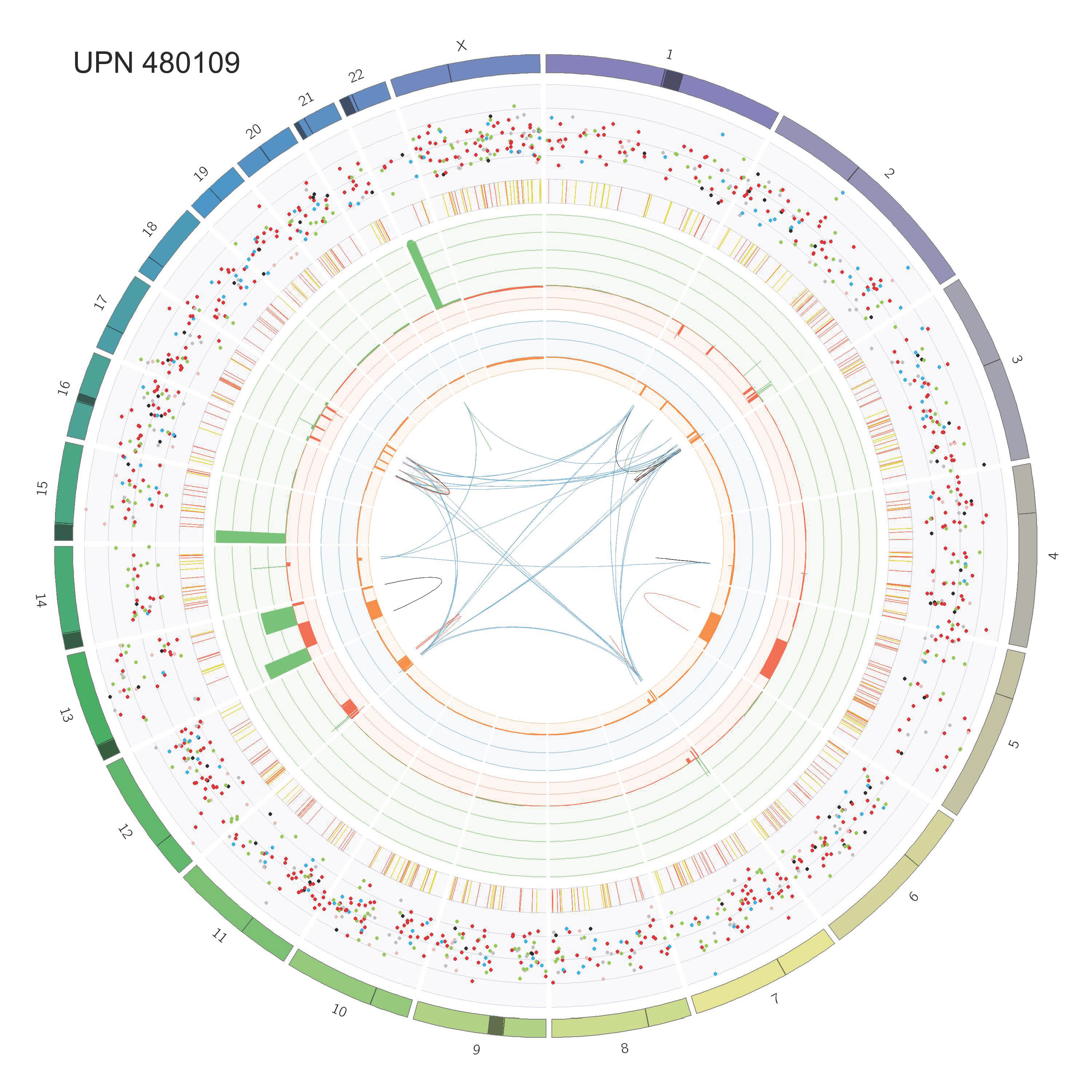

### Supp_Fig_2_circos_Page_31.jpg

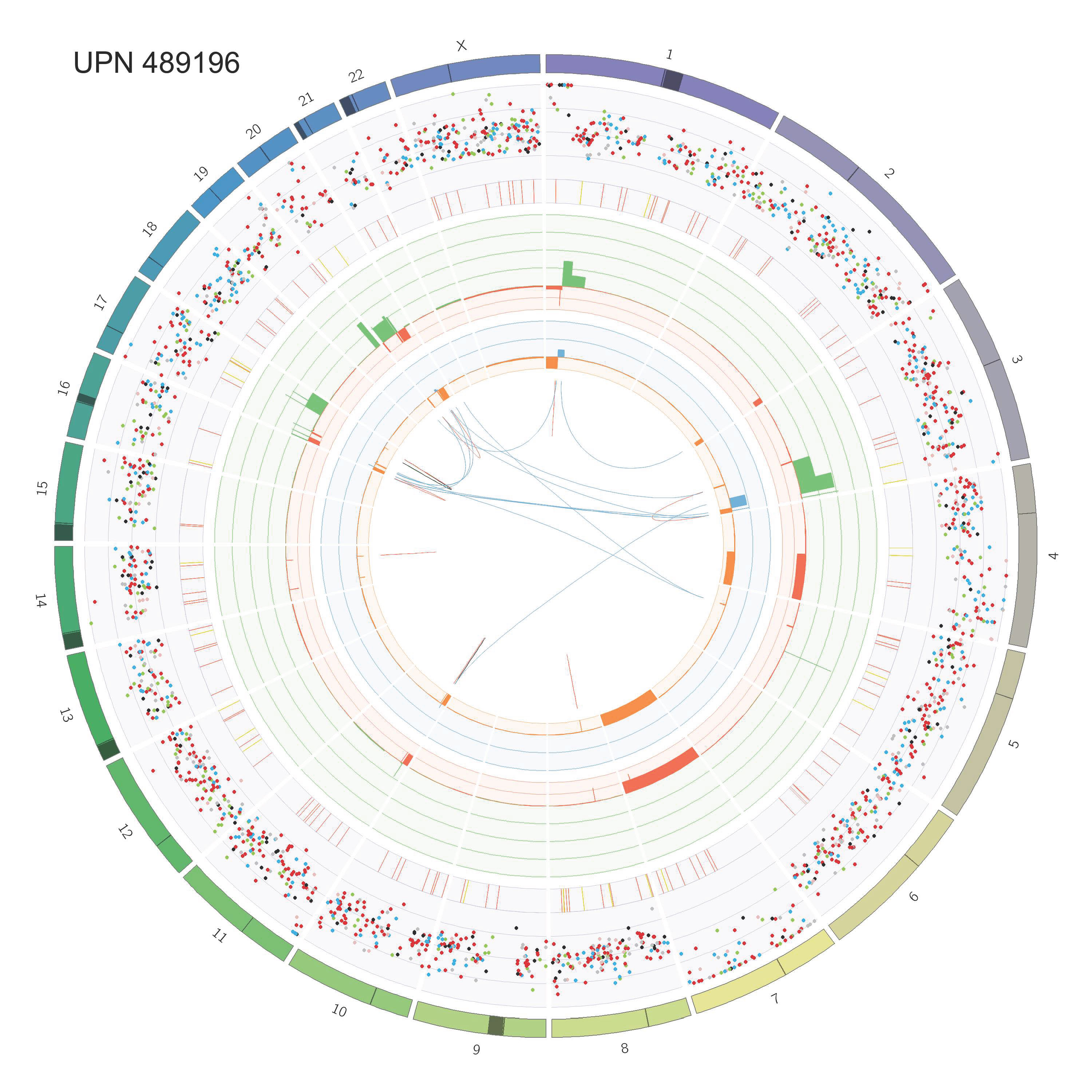

### Supp_Fig_2_circos_Page_32.jpg

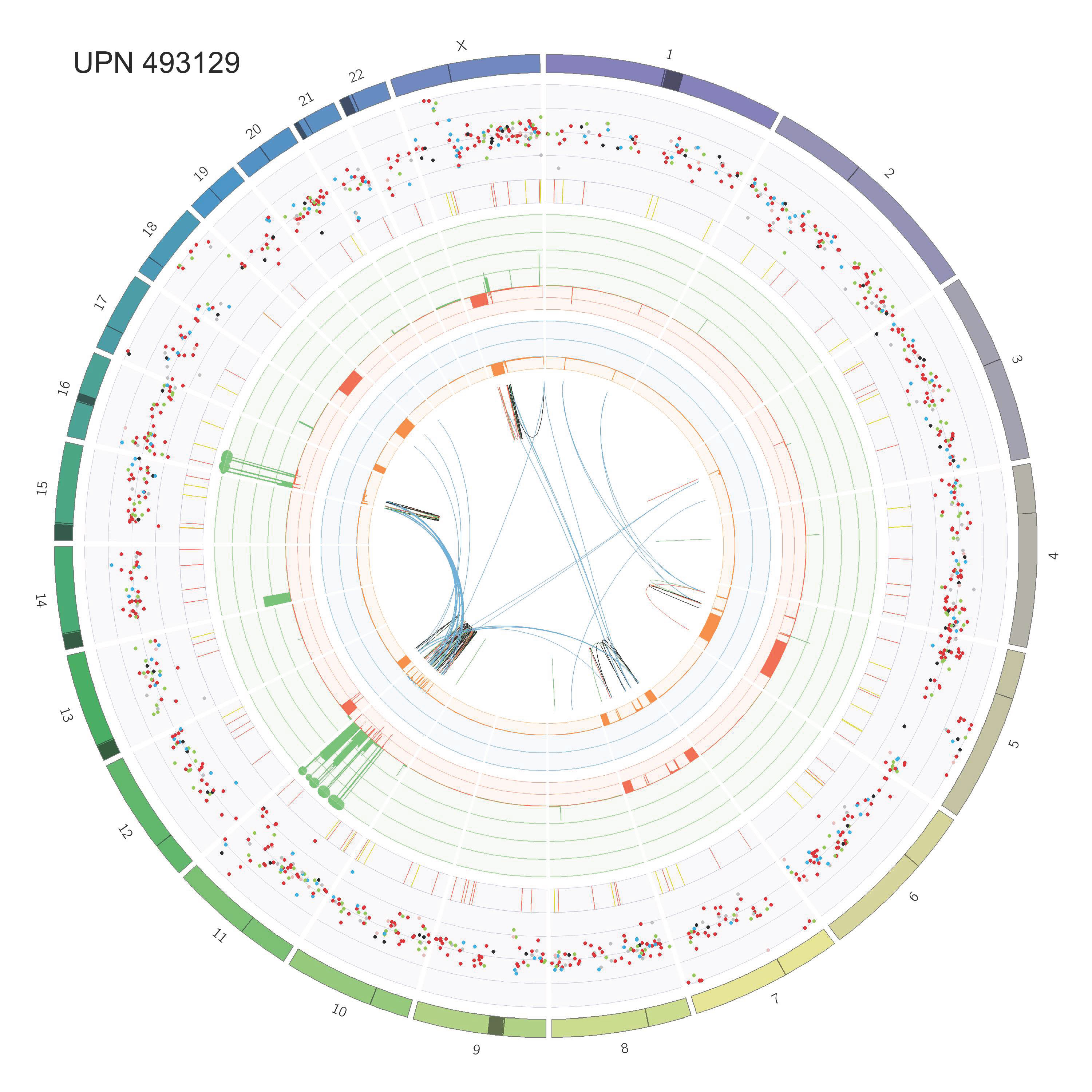

### Supp_Fig_2_circos_Page_33.jpg

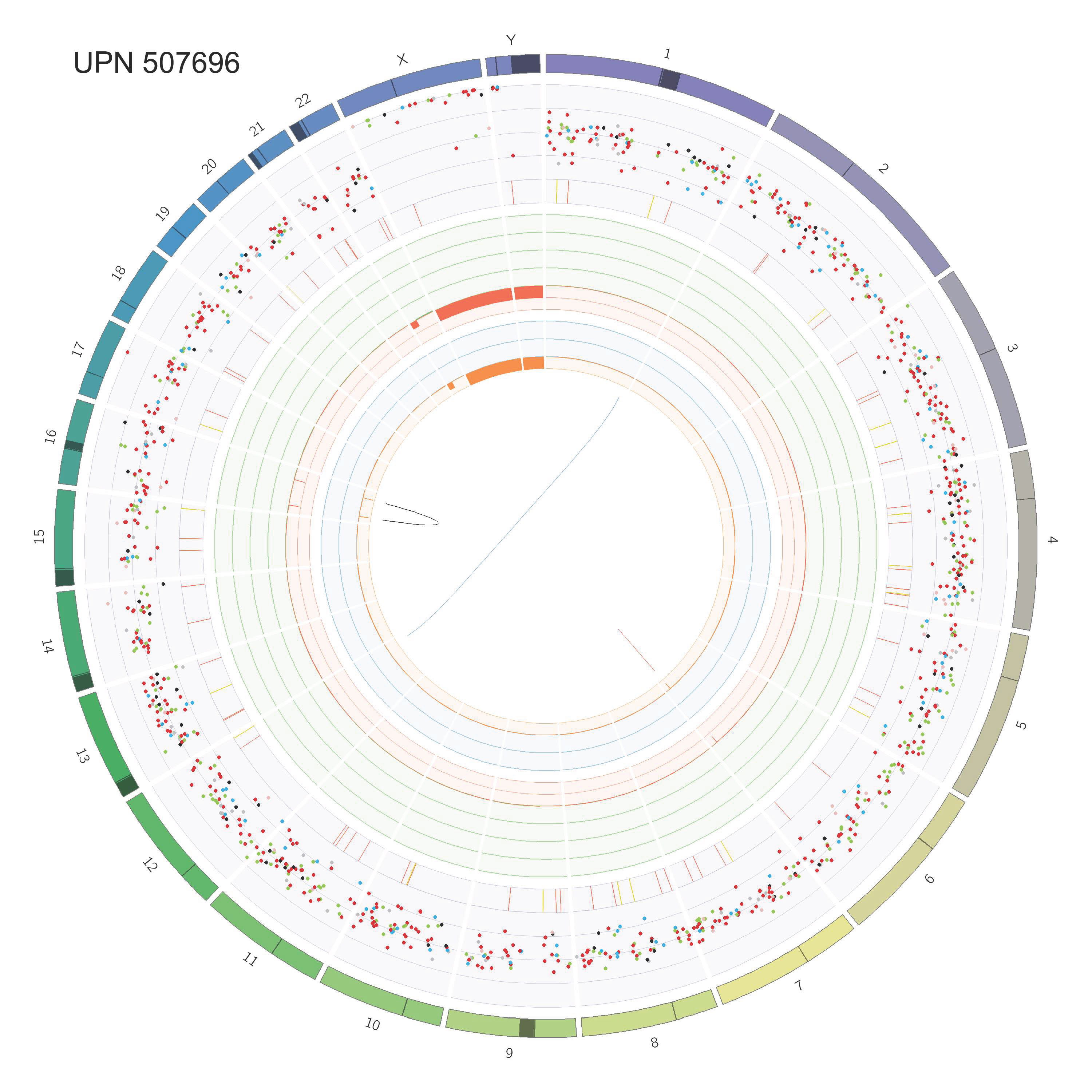

### Supp_Fig_2_circos_Page_34.jpg

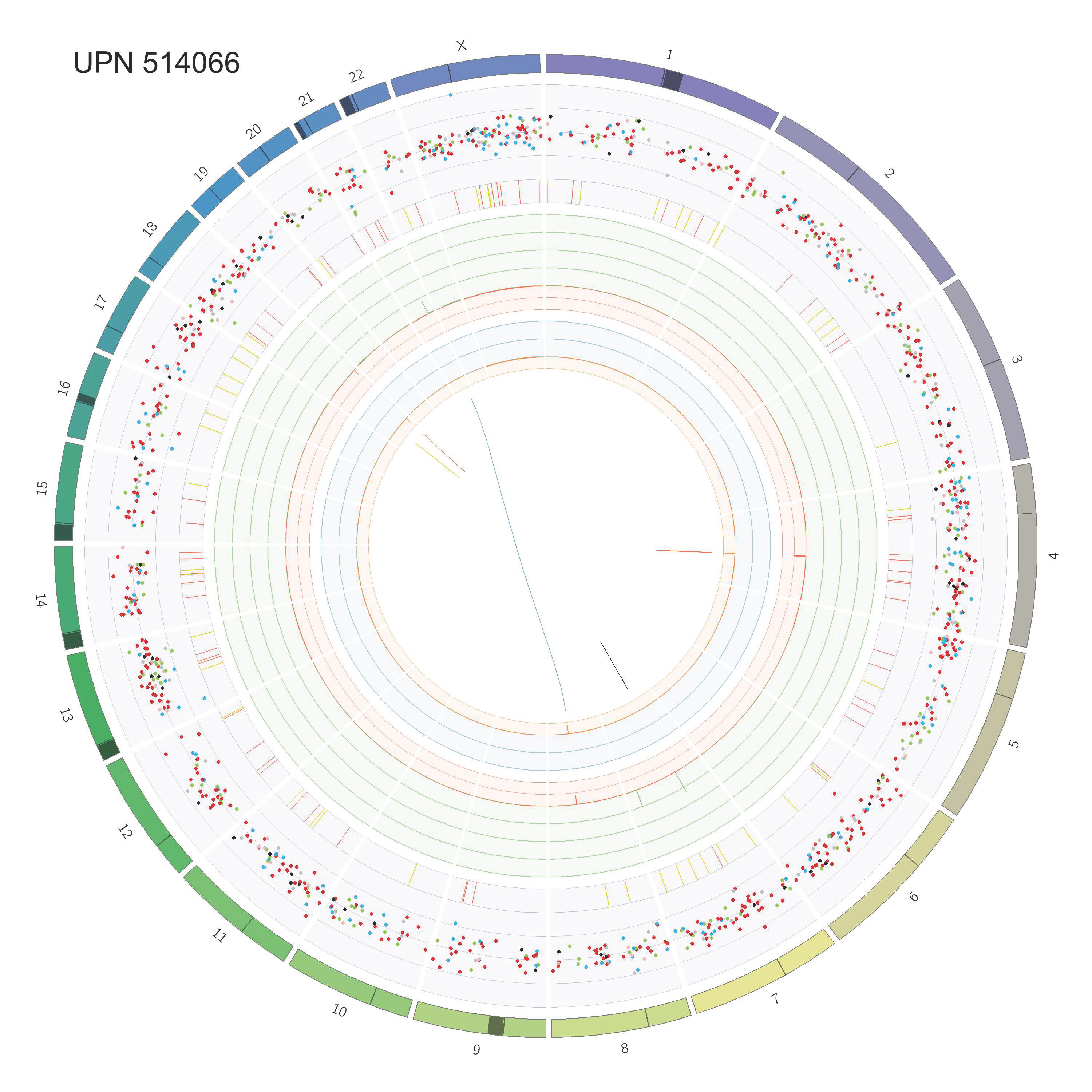

### Supp_Fig_2_circos_Page_35.jpg

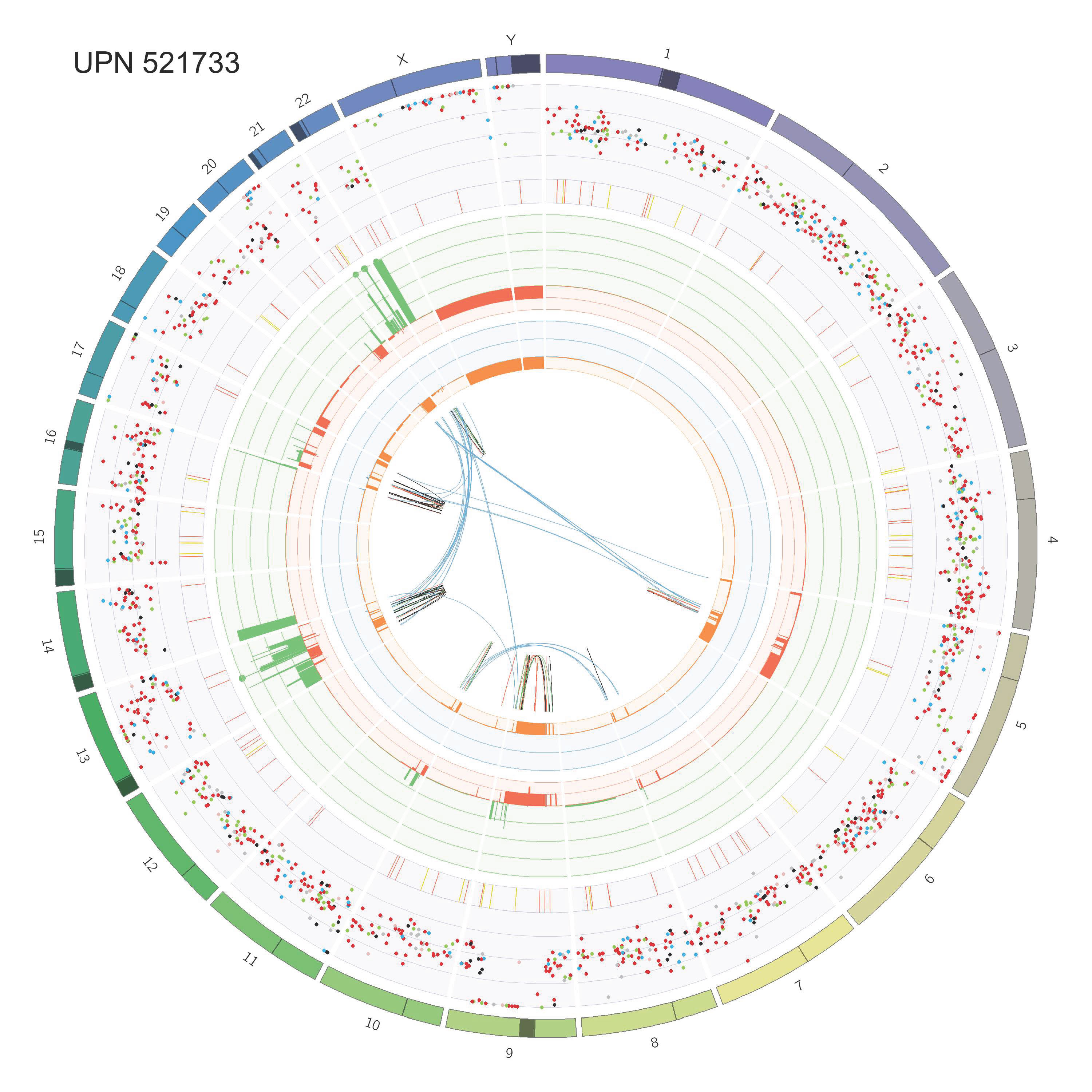

### Supp_Fig_2_circos_Page_36.jpg

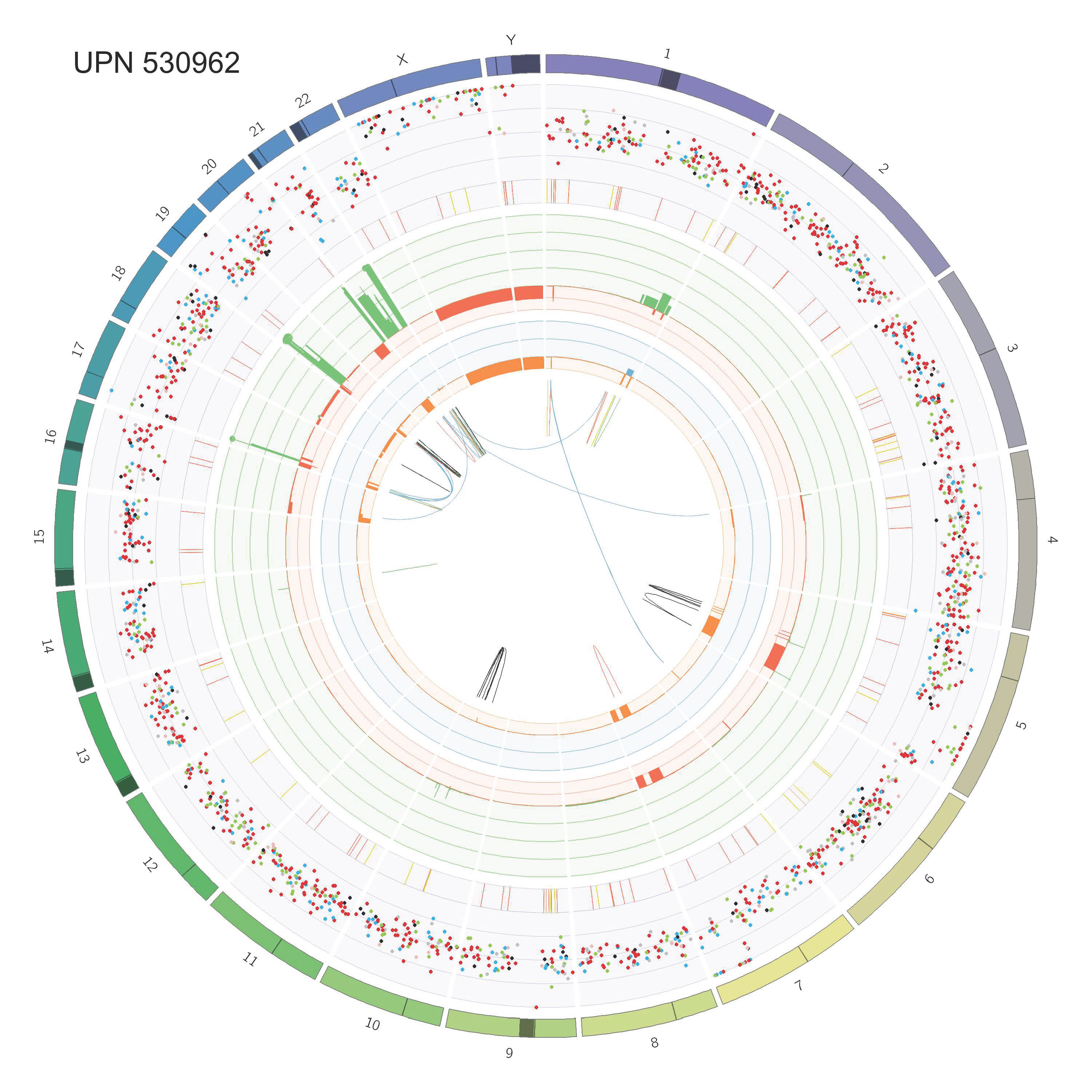

### Supp_Fig_2_circos_Page_37.jpg

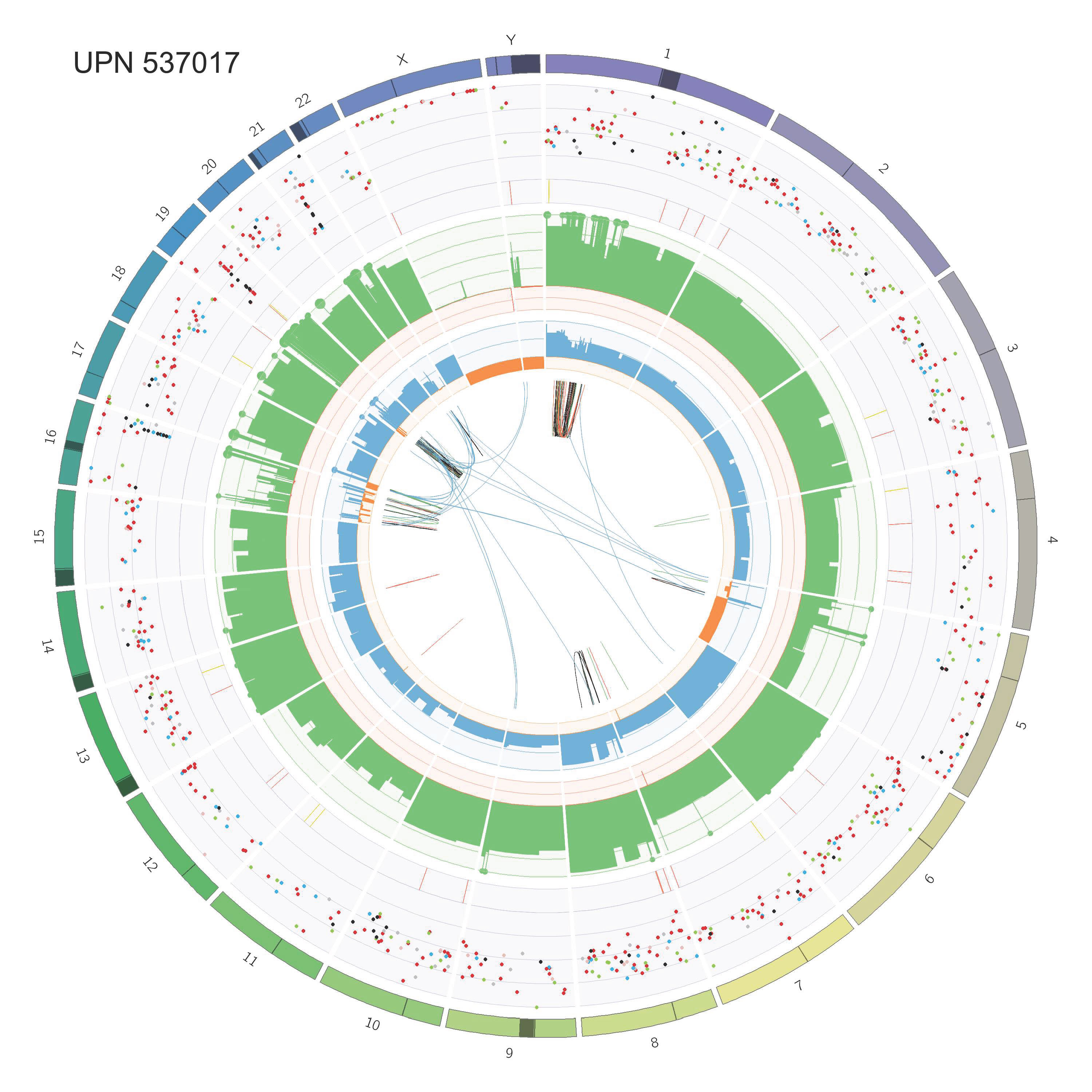

### Supp_Fig_2_circos_Page_38.jpg

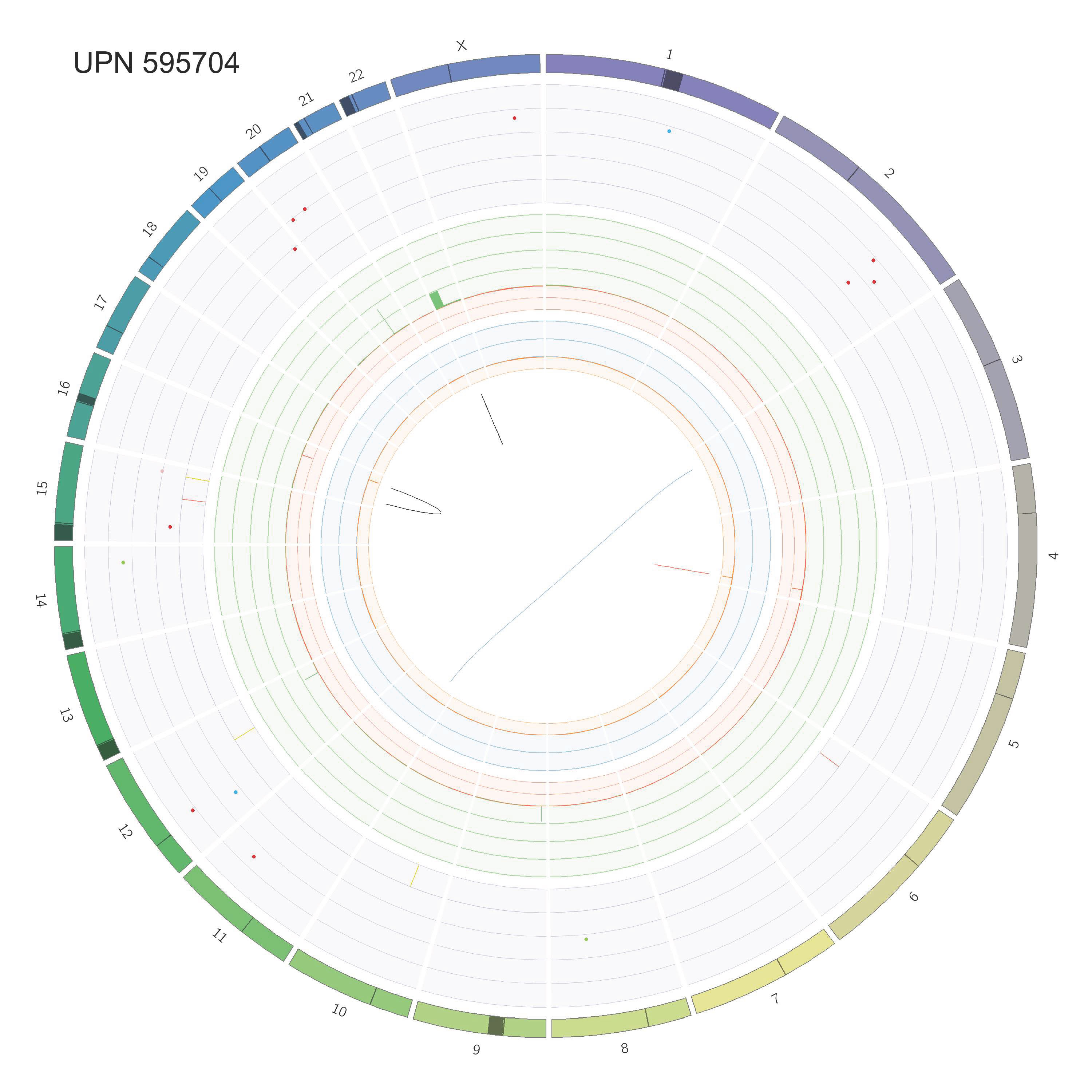

### Supp_Fig_2_circos_Page_39.jpg

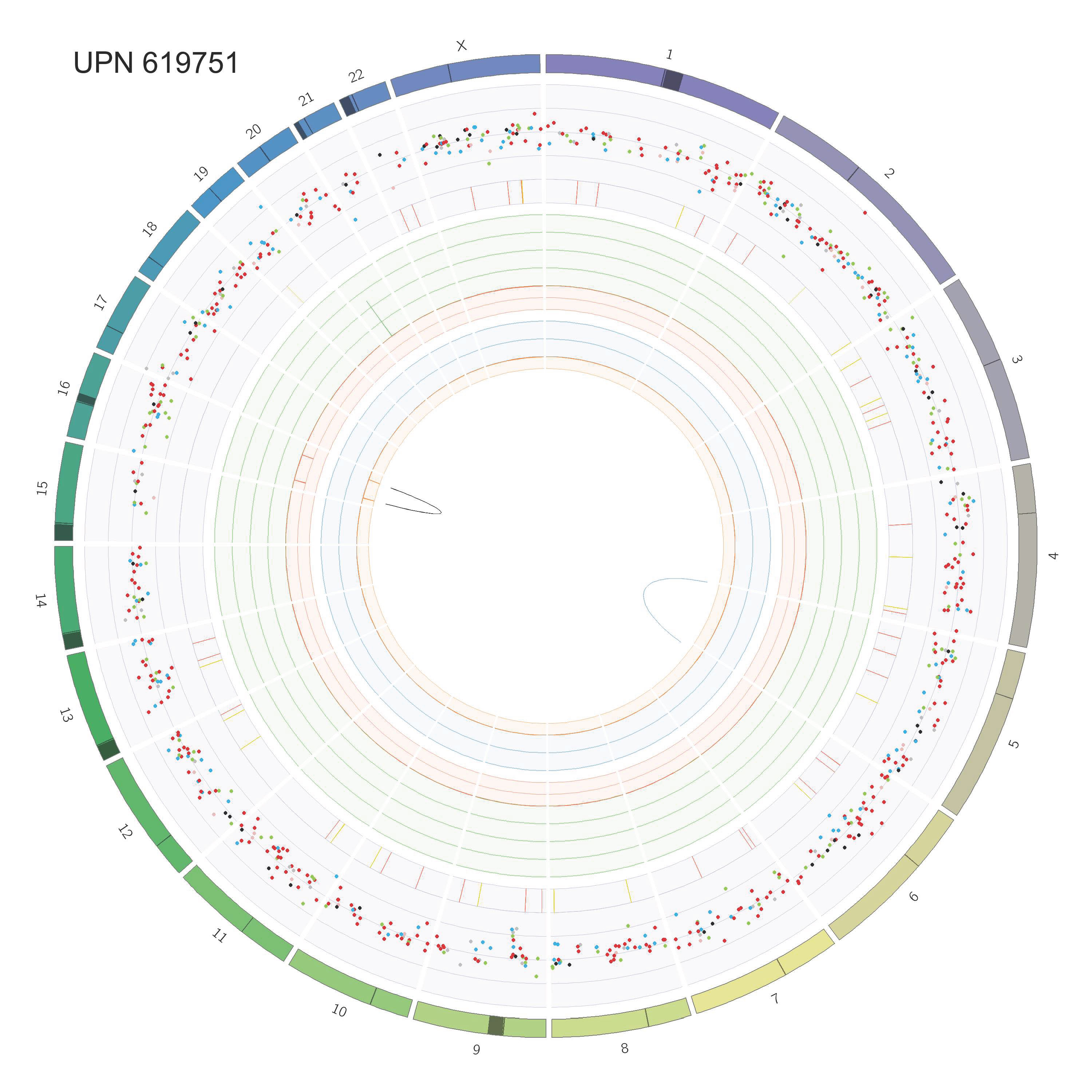

### Supp_Fig_2_circos_Page_40.jpg

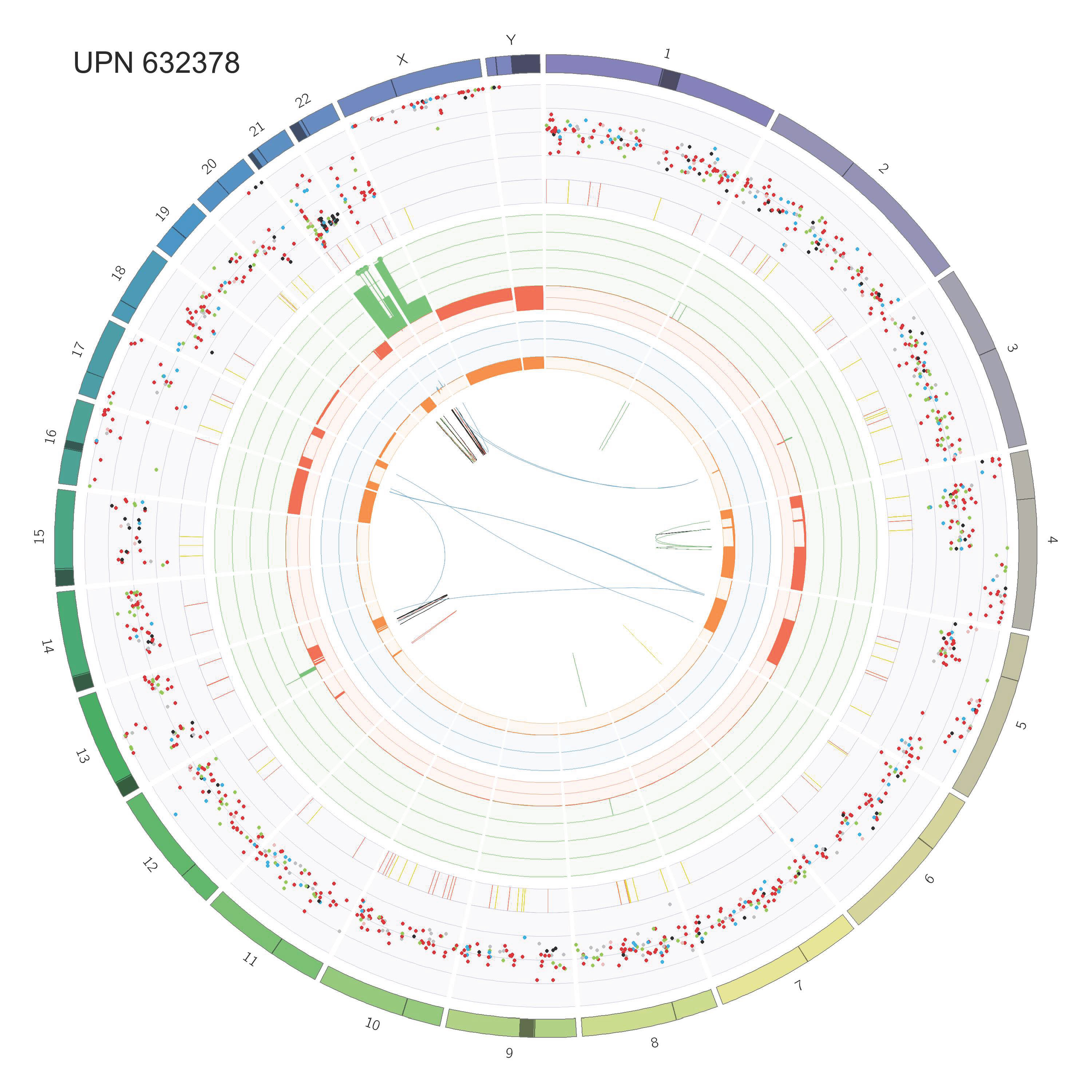

### Supp_Fig_2_circos_Page_41.jpg

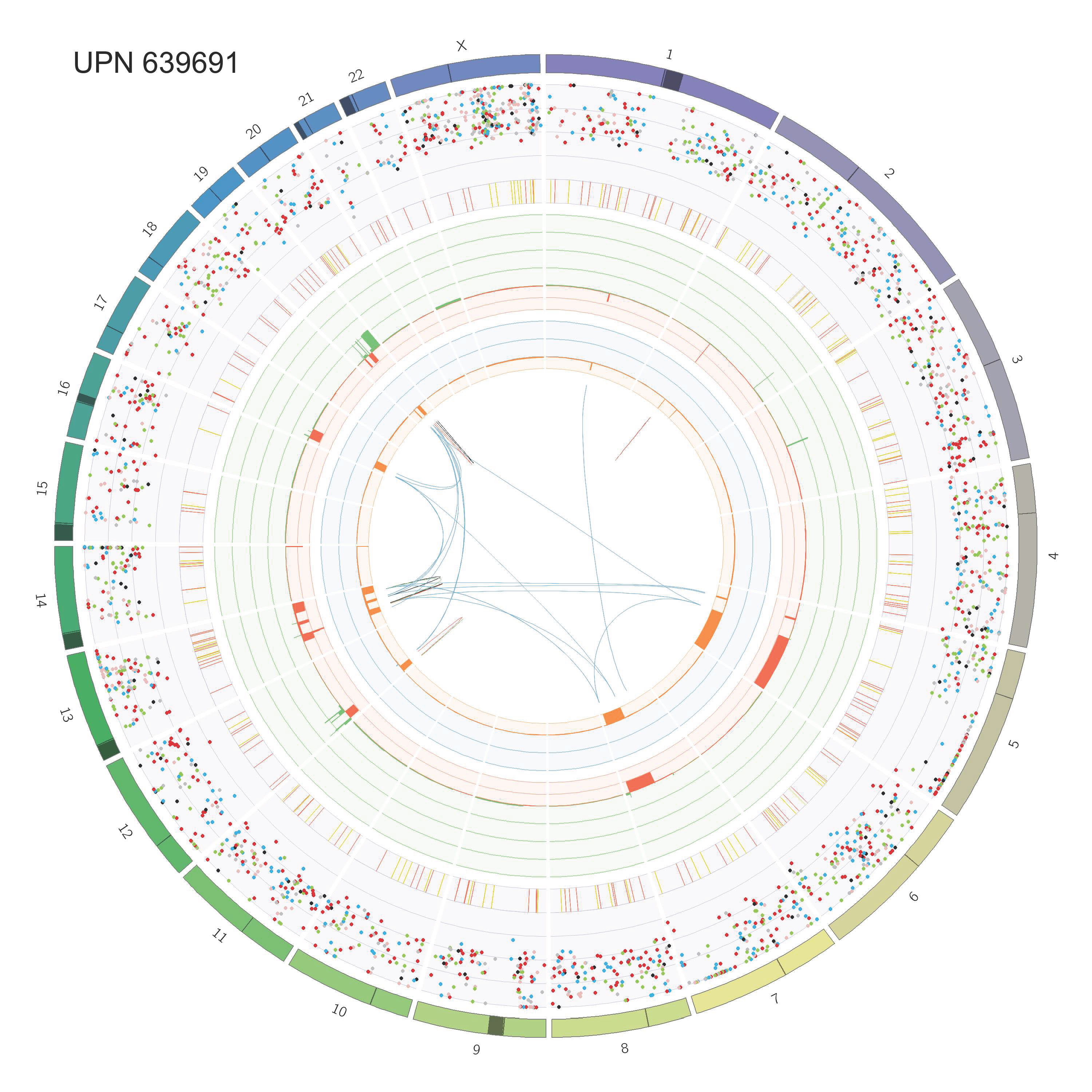

### Supp_Fig_2_circos_Page_42.jpg

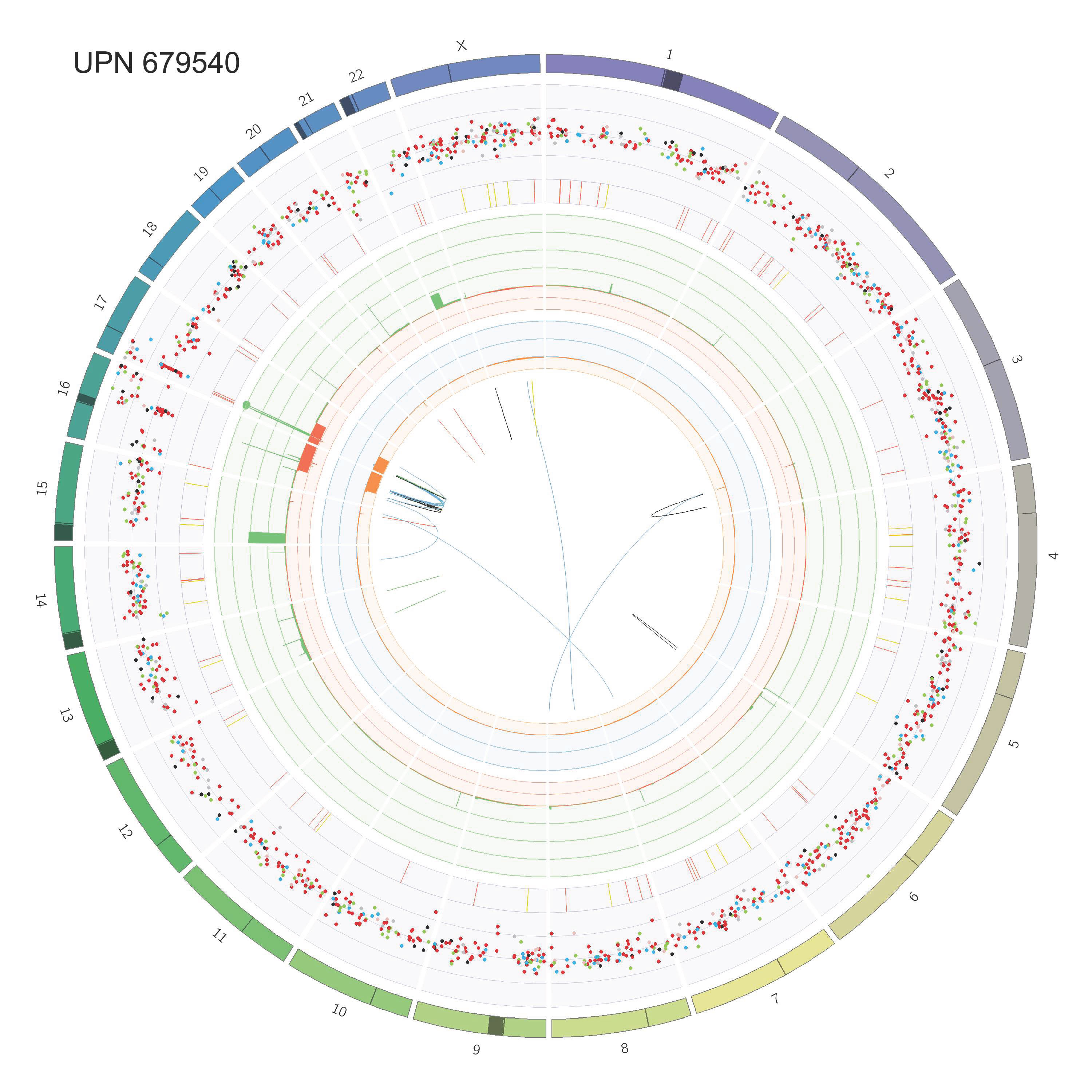

### Supp_Fig_2_circos_Page_43.jpg

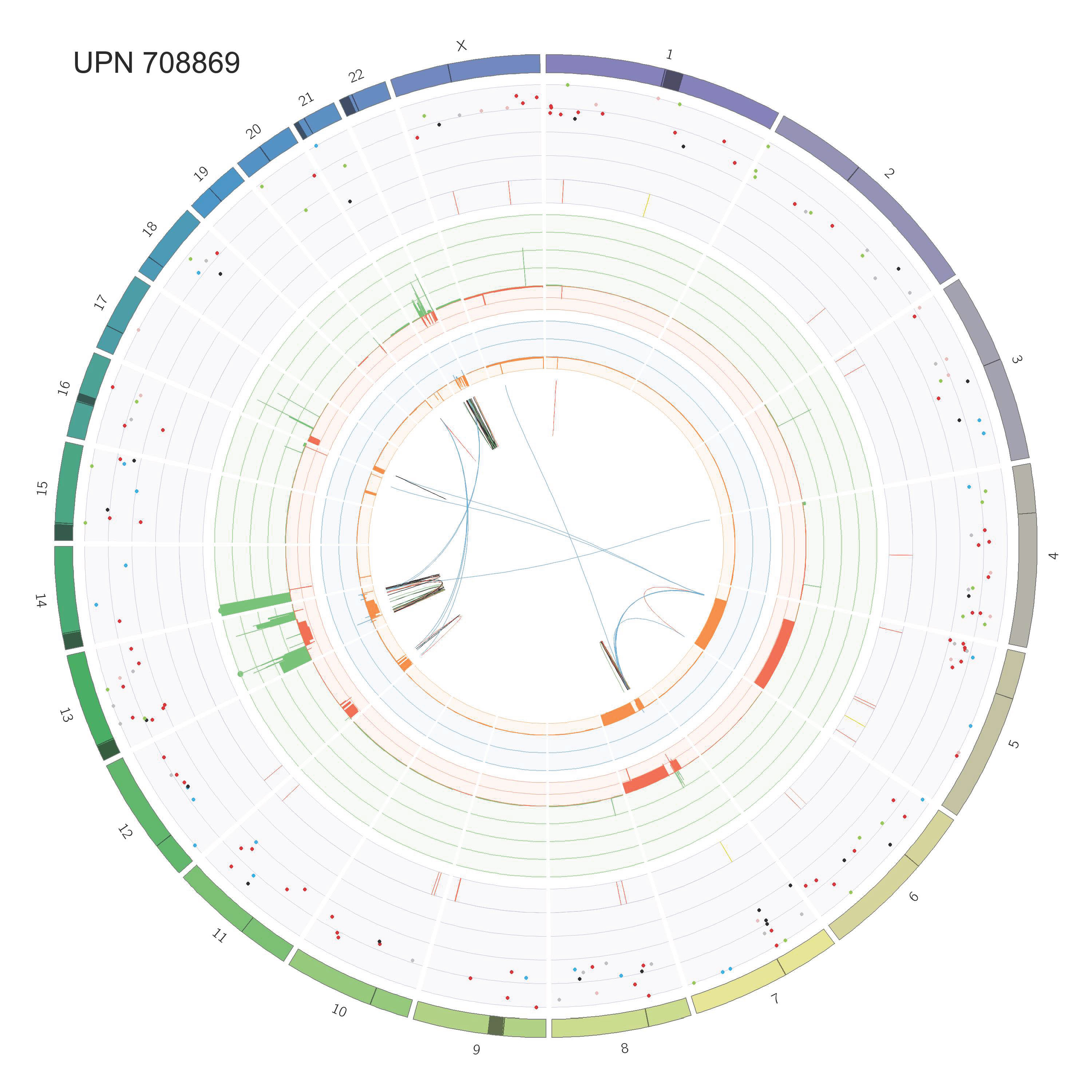

### Supp_Fig_2_circos_Page_44.jpg

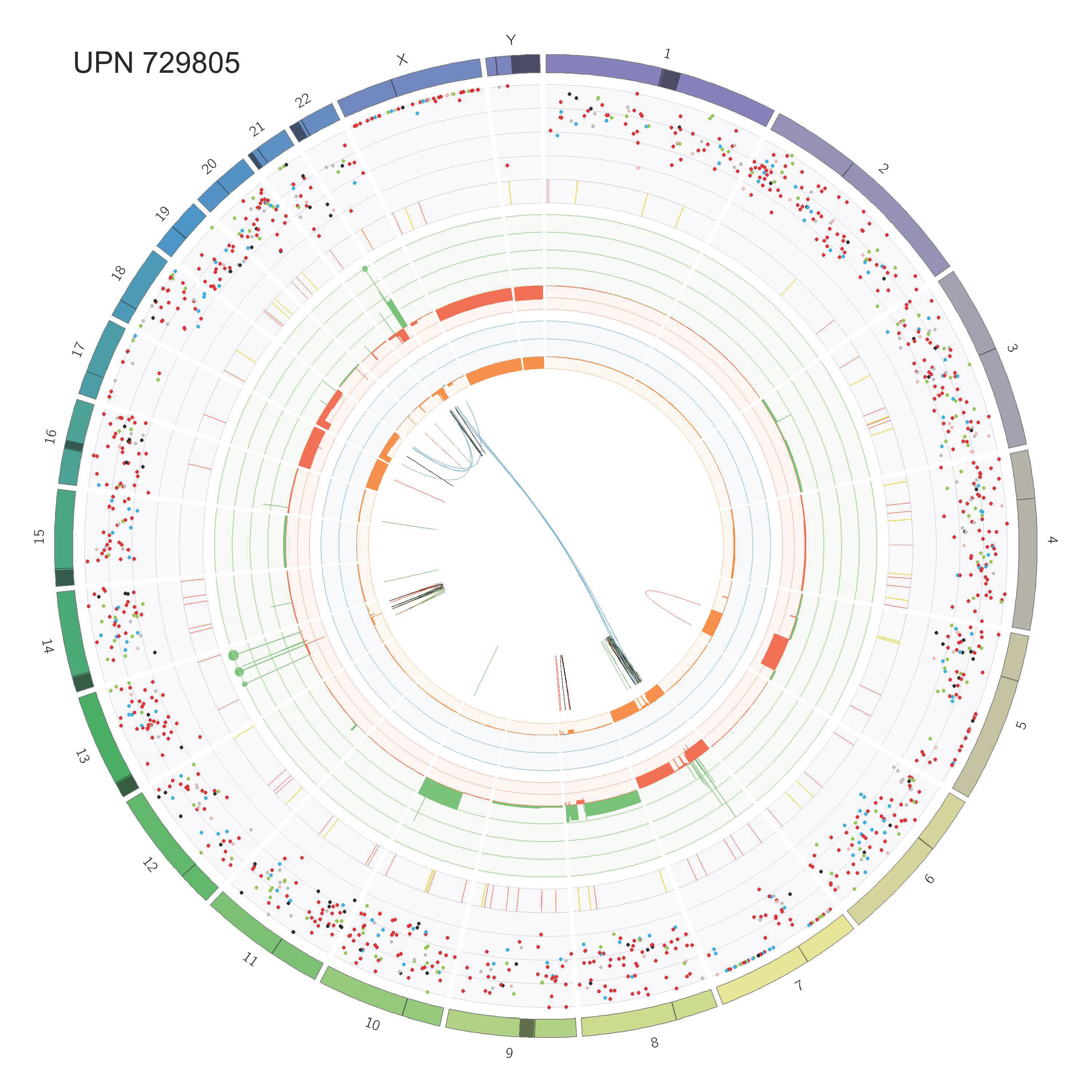

### Supp_Fig_2_circos_Page_45.jpg

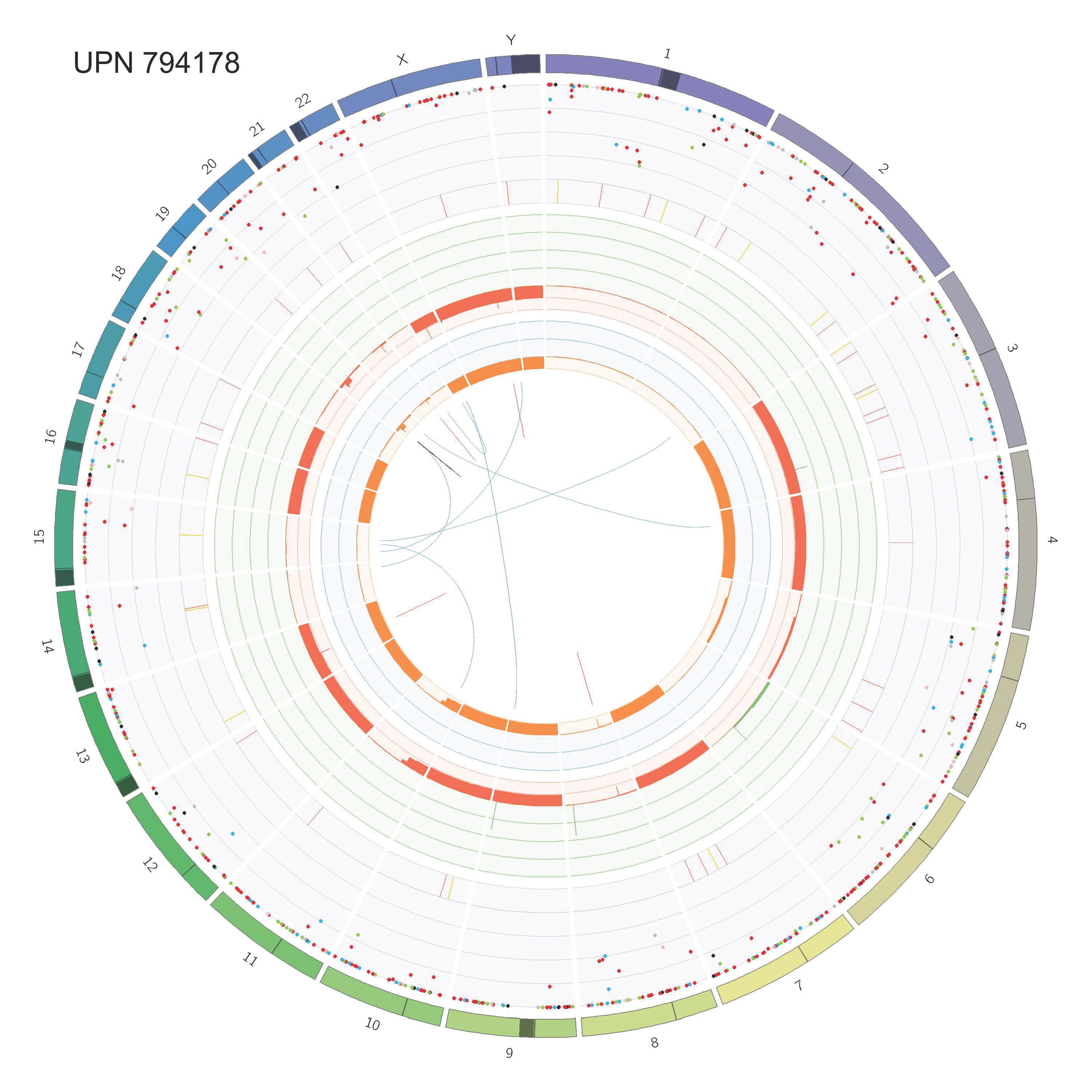

### Supp_Fig_2_circos_Page_46.jpg

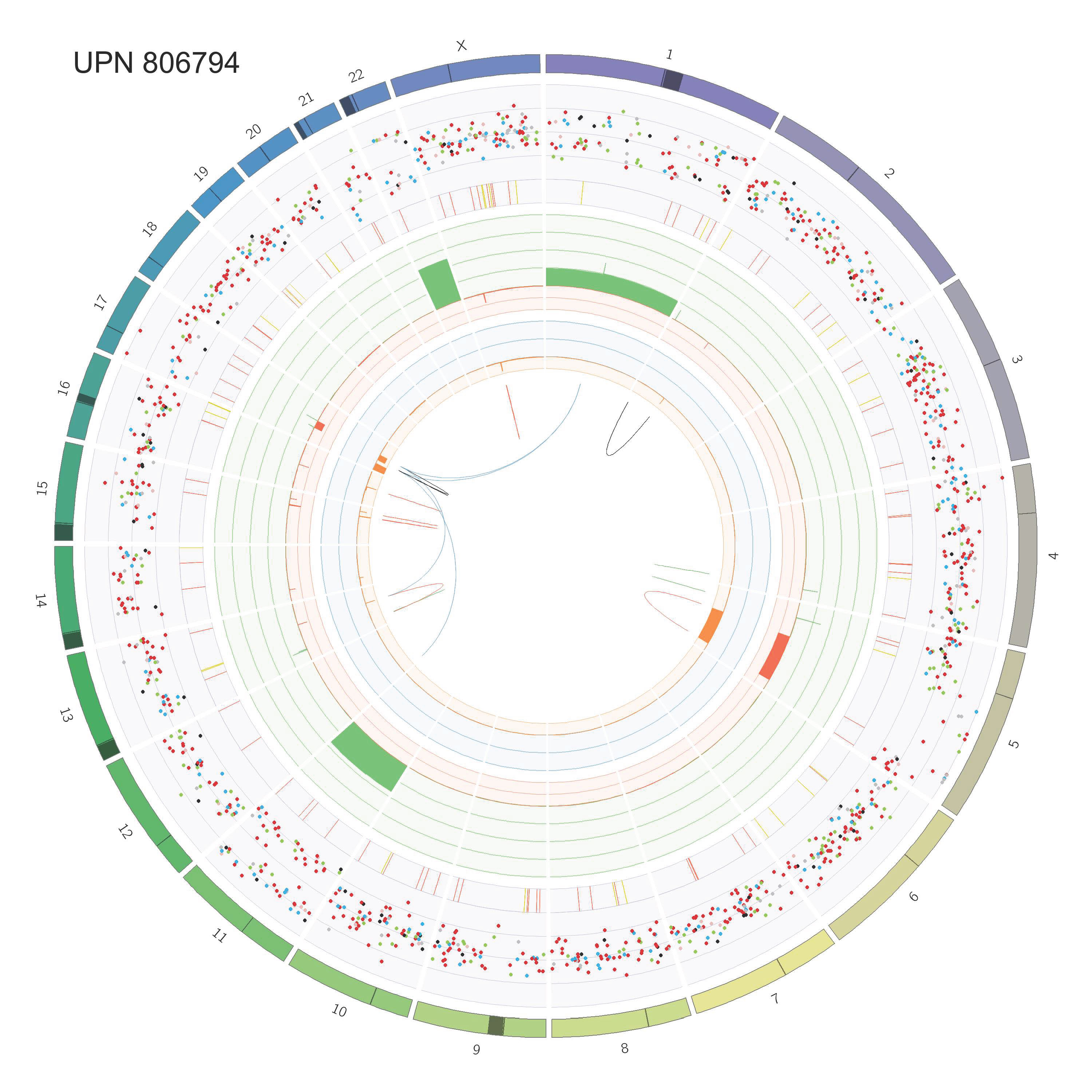

### Supp_Fig_2_circos_Page_47.jpg

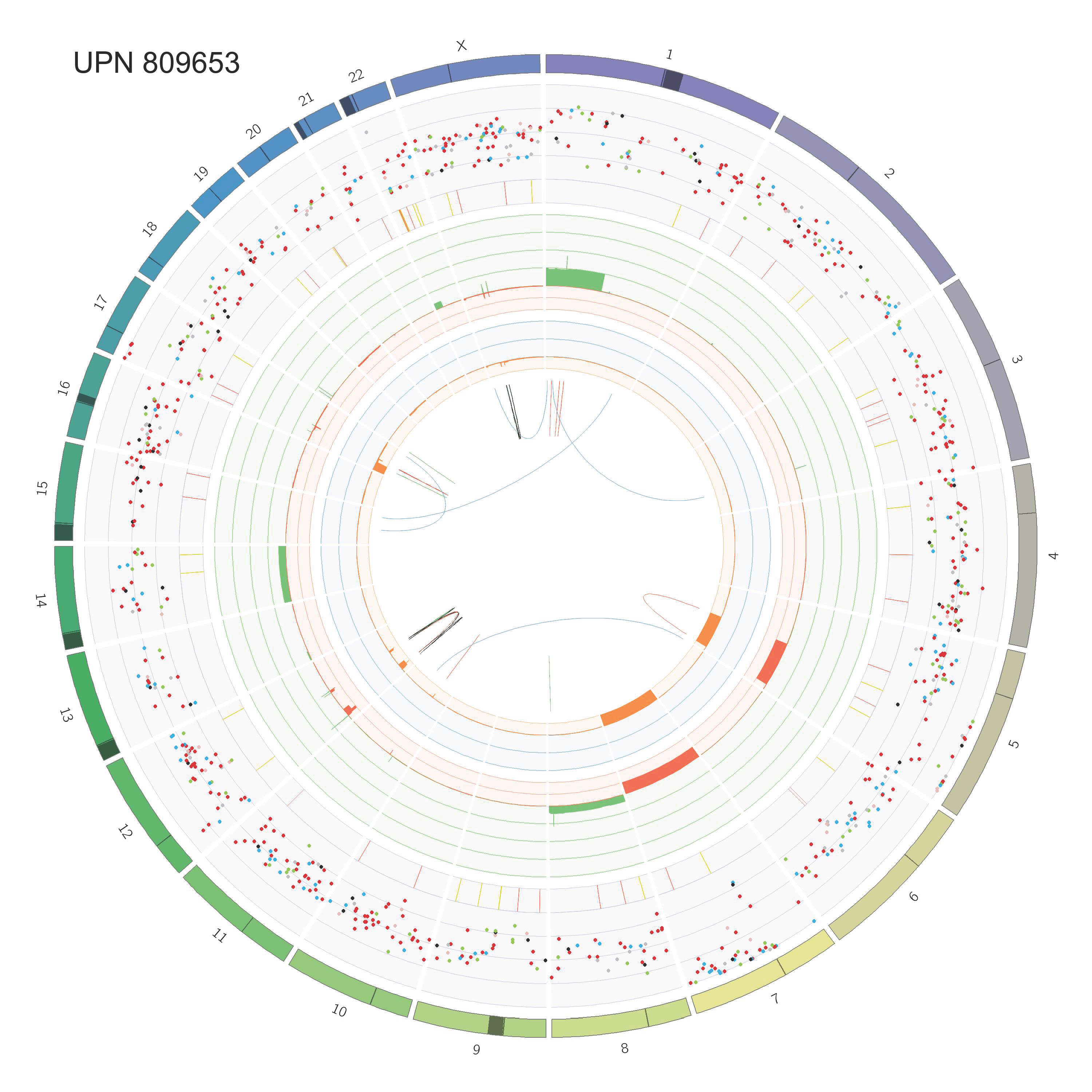

### Supp_Fig_2_circos_Page_48.jpg

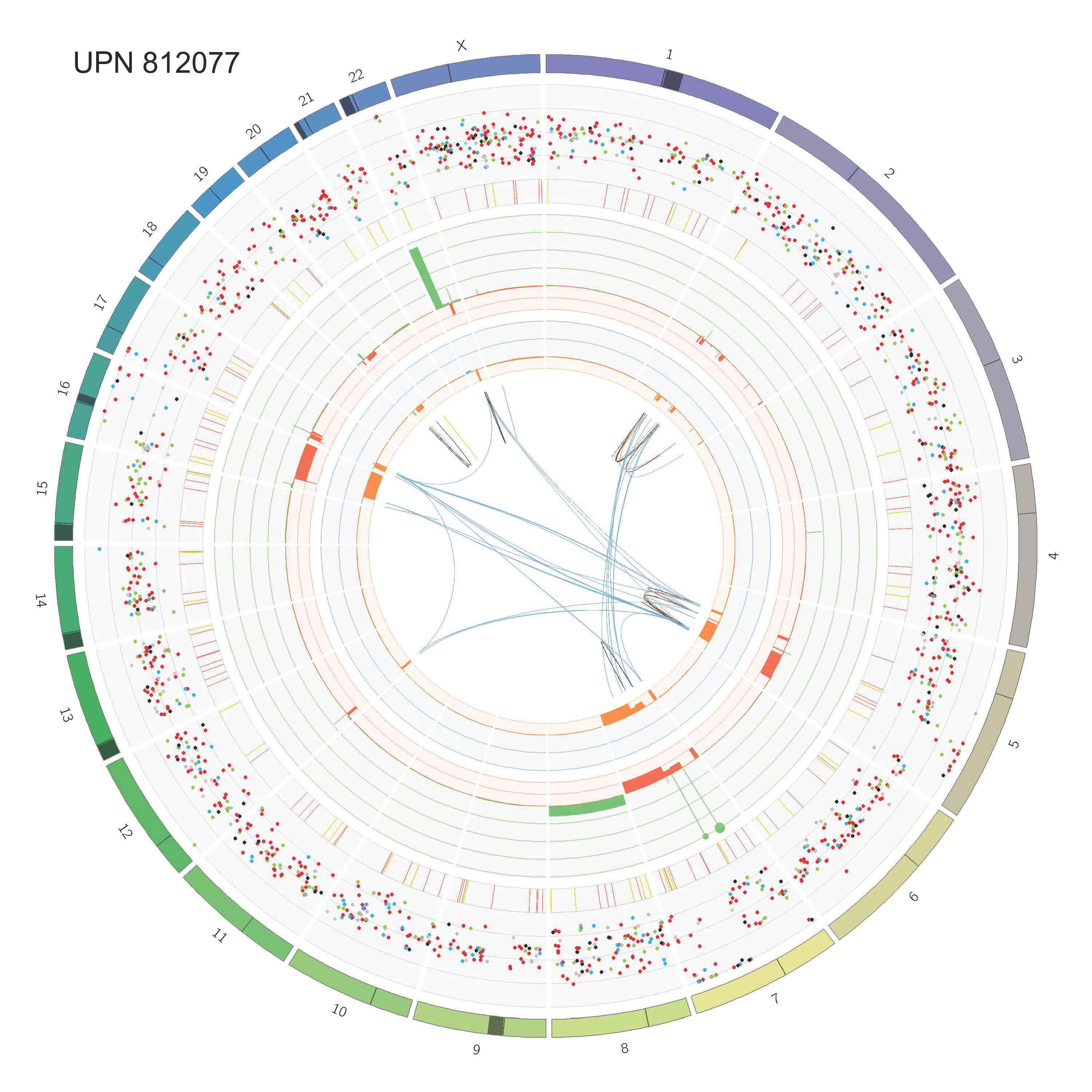

### Supp_Fig_2_circos_Page_49.jpg

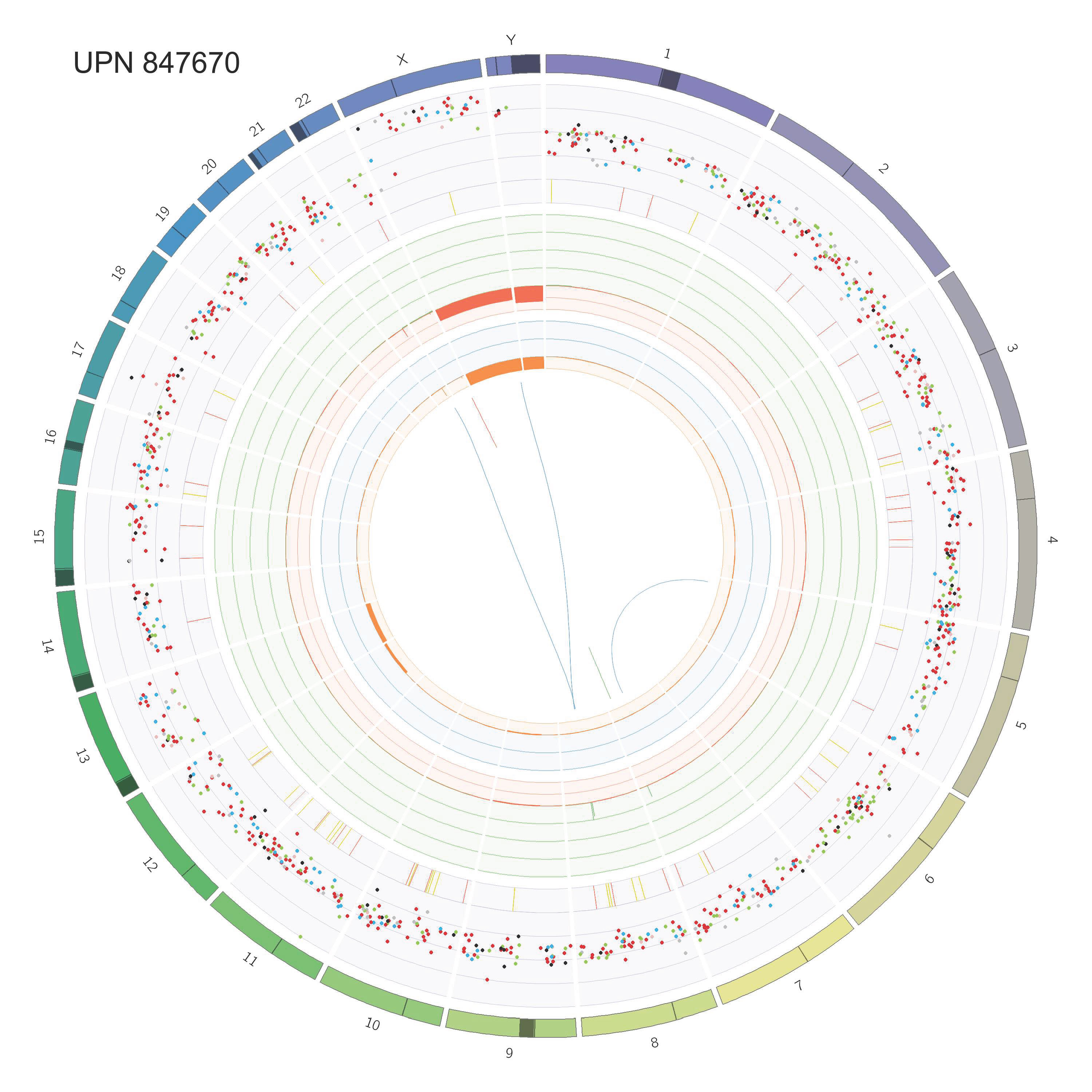

### Supp_Fig_2_circos_Page_50.jpg

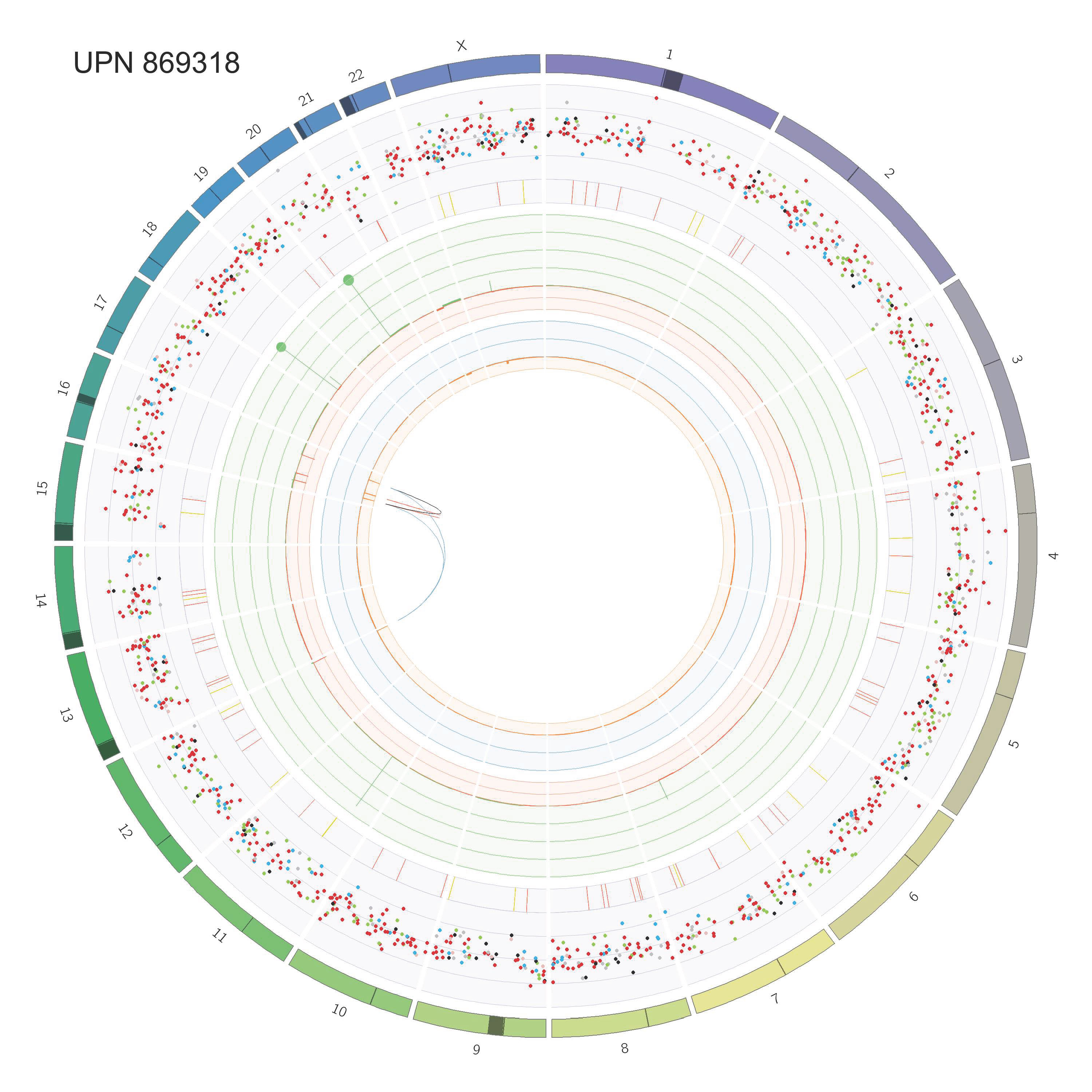

### Supp_Fig_2_circos_Page_51.jpg

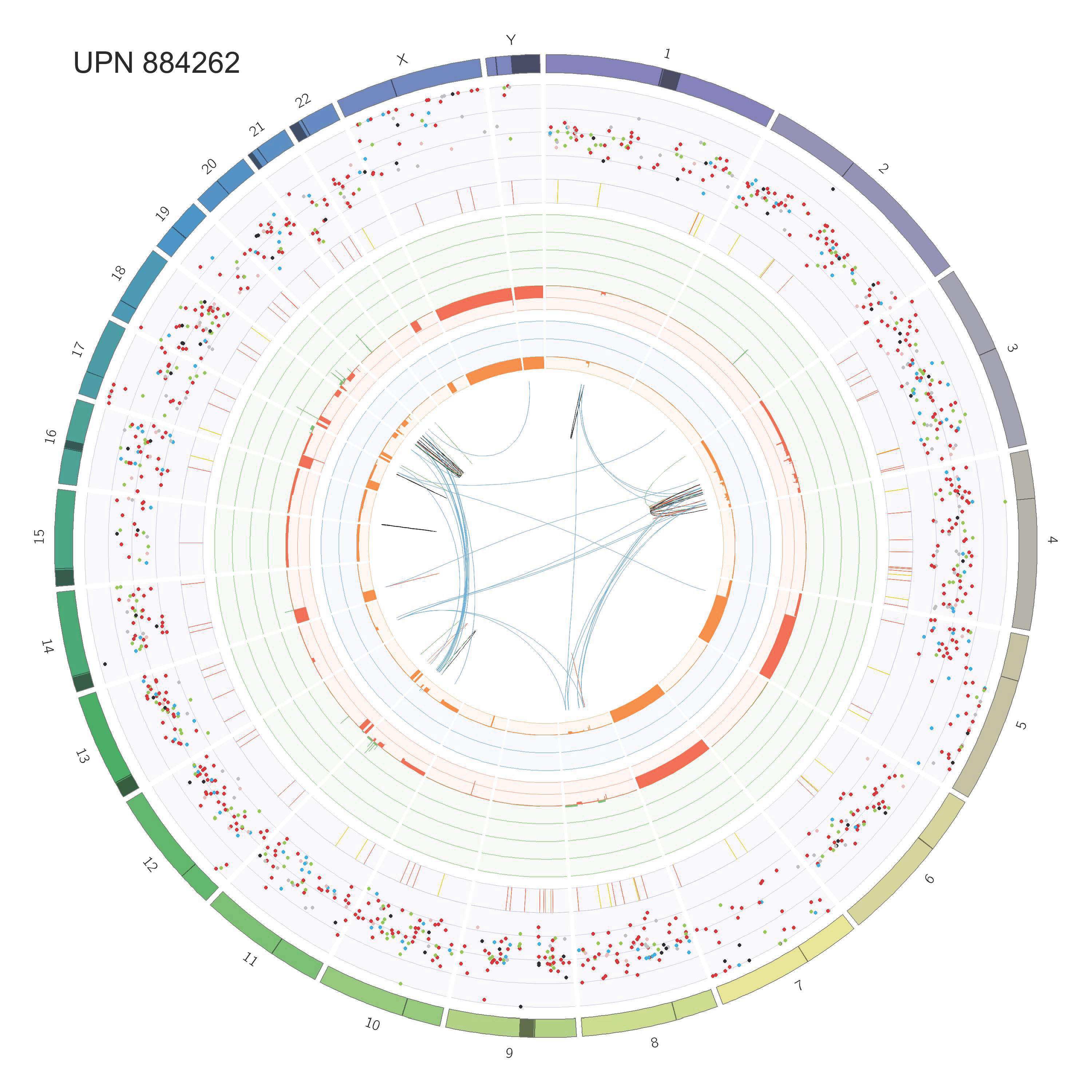

### Supp_Fig_2_circos_Page_52.jpg

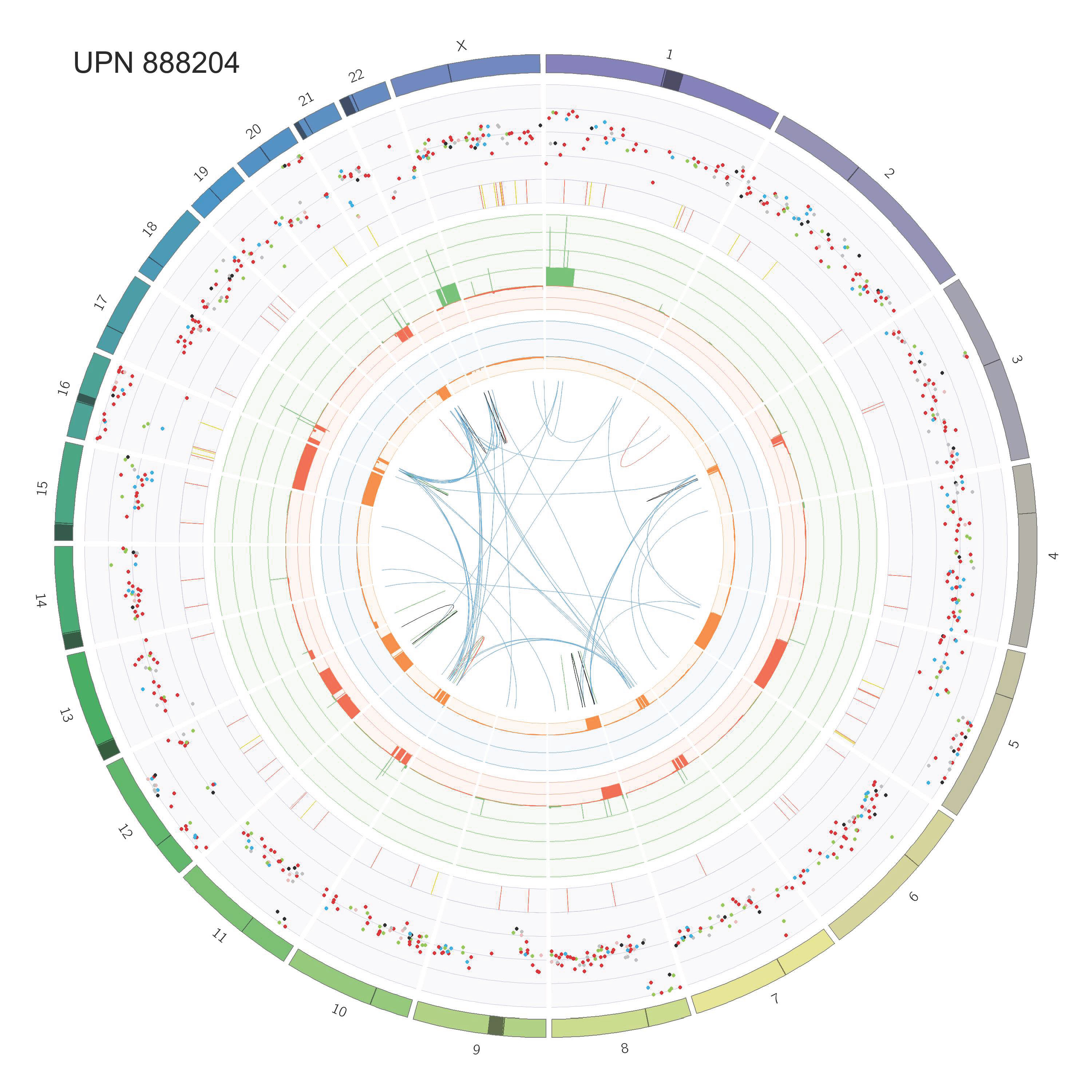

### Supp_Fig_2_circos_Page_53.jpg

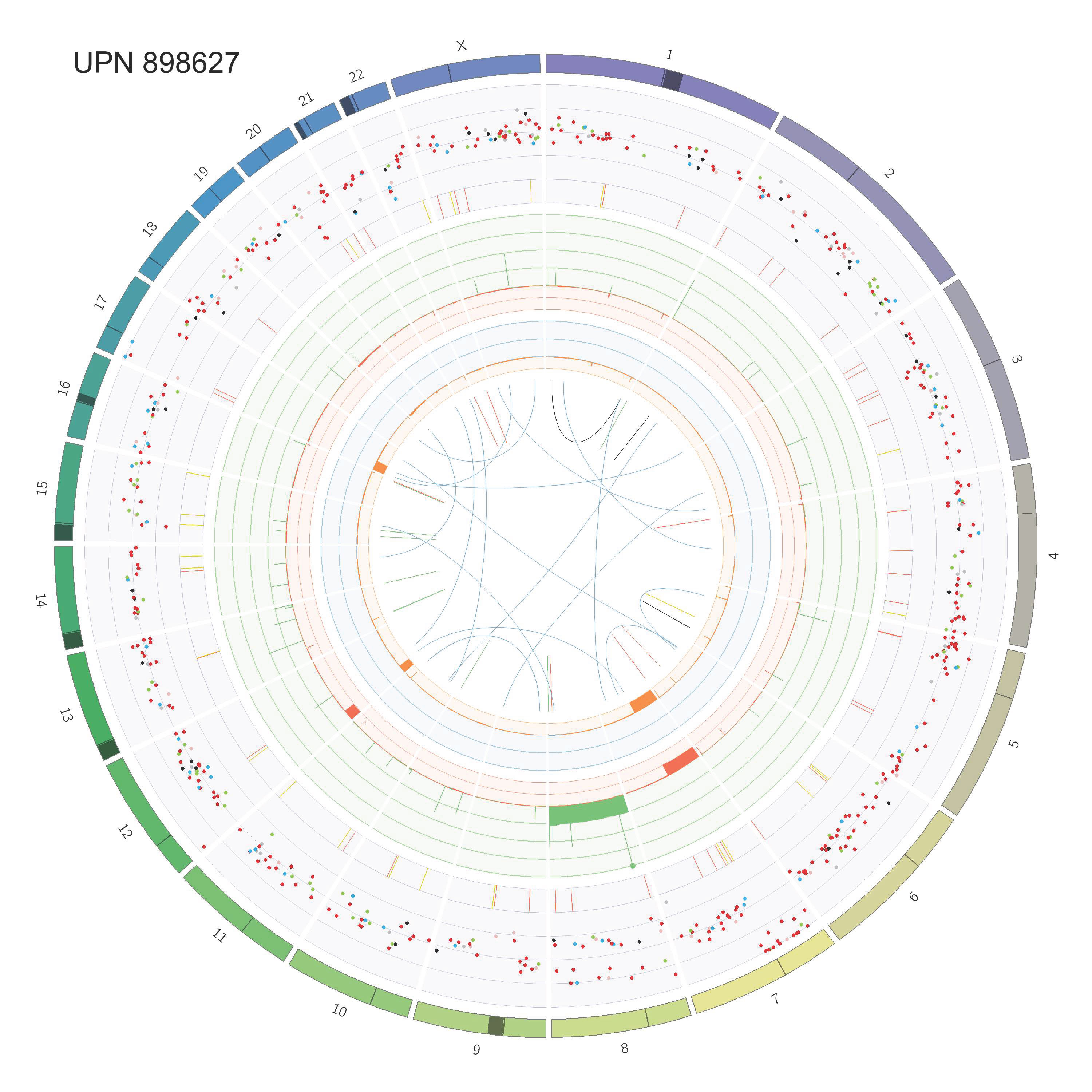

### Supp_Fig_2_circos_Page_54.jpg

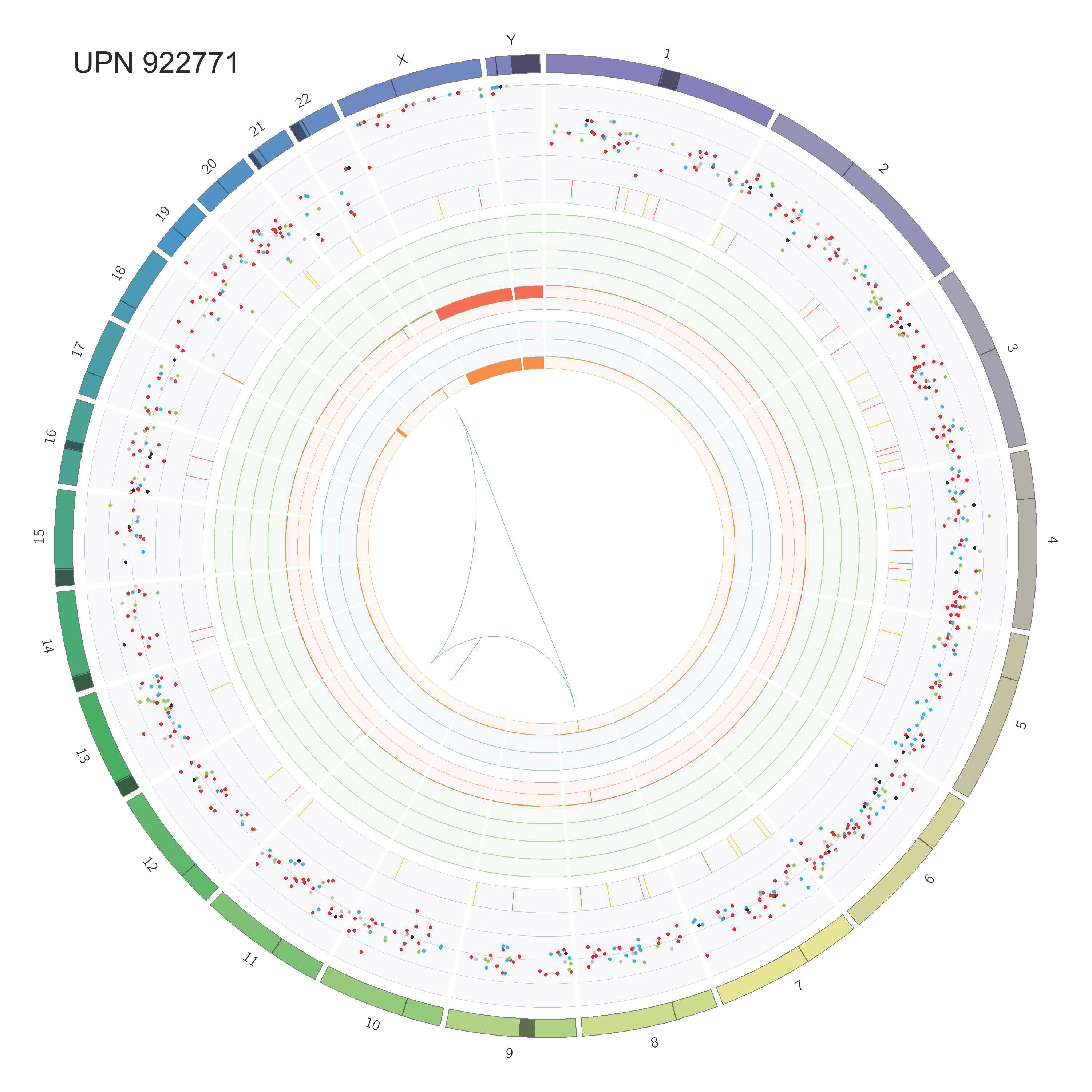

### Supp_Fig_2_circos_Page_55.jpg

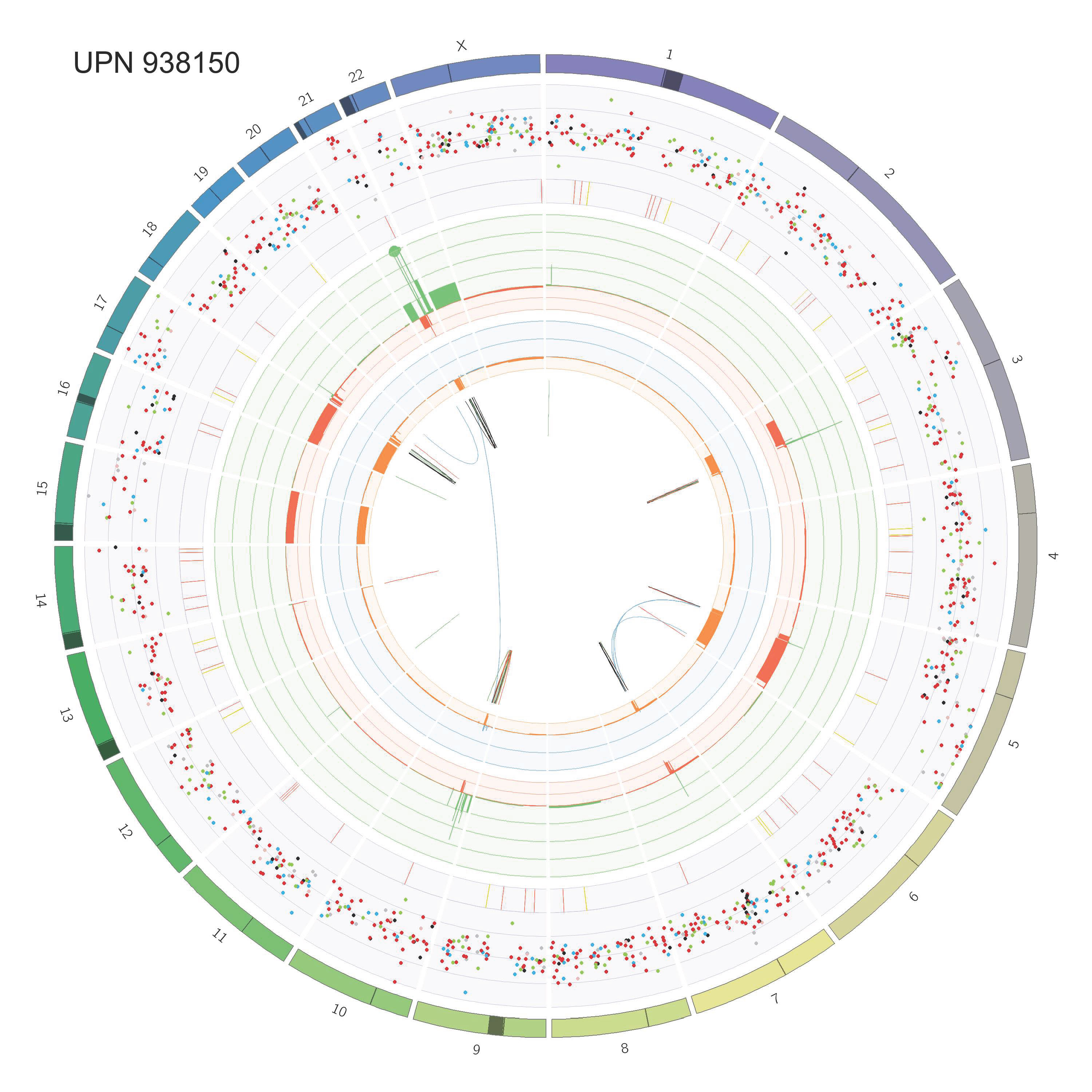

### Supp_Fig_2_circos_Page_56.jpg

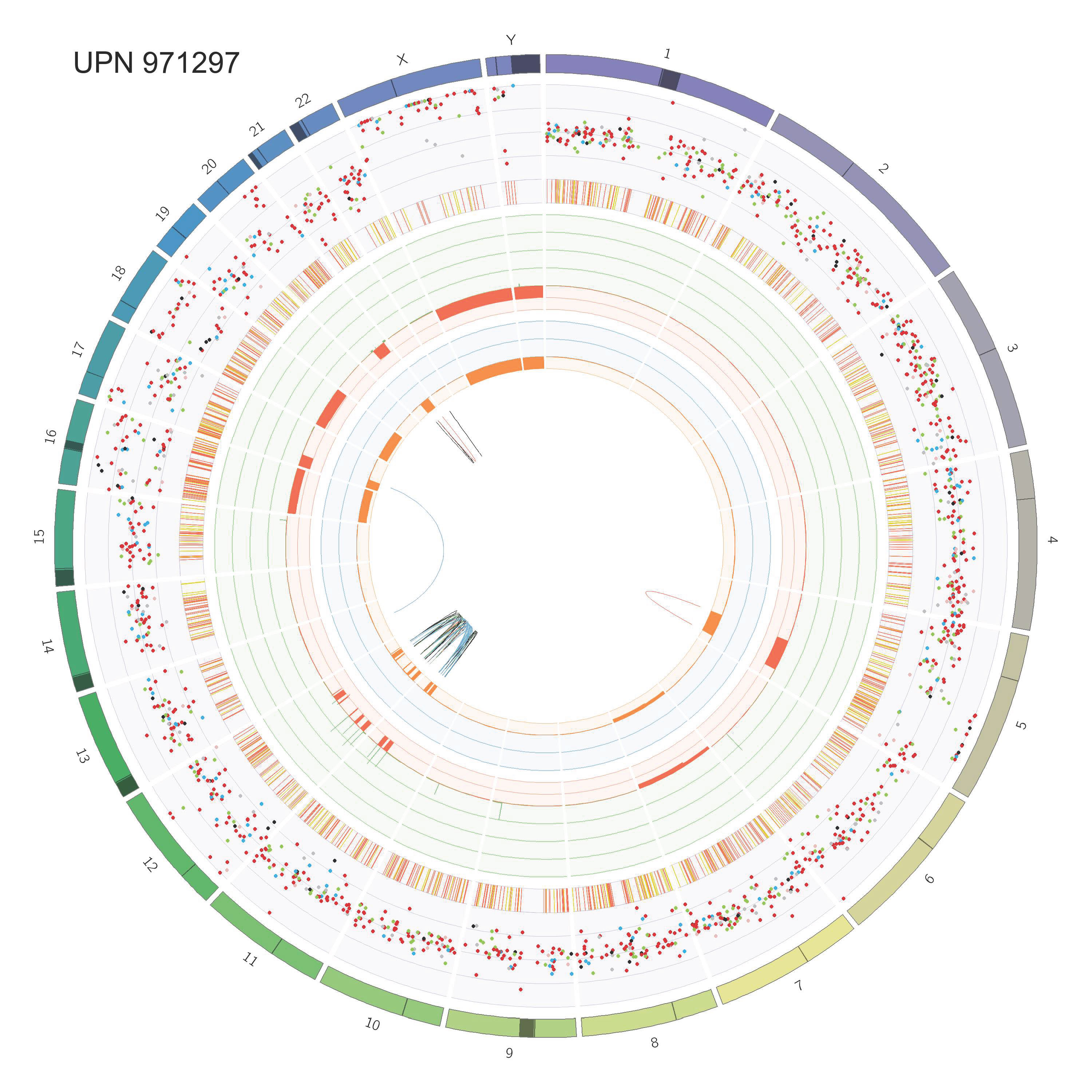

### Supp_Fig_2_circos_Page_57.jpg

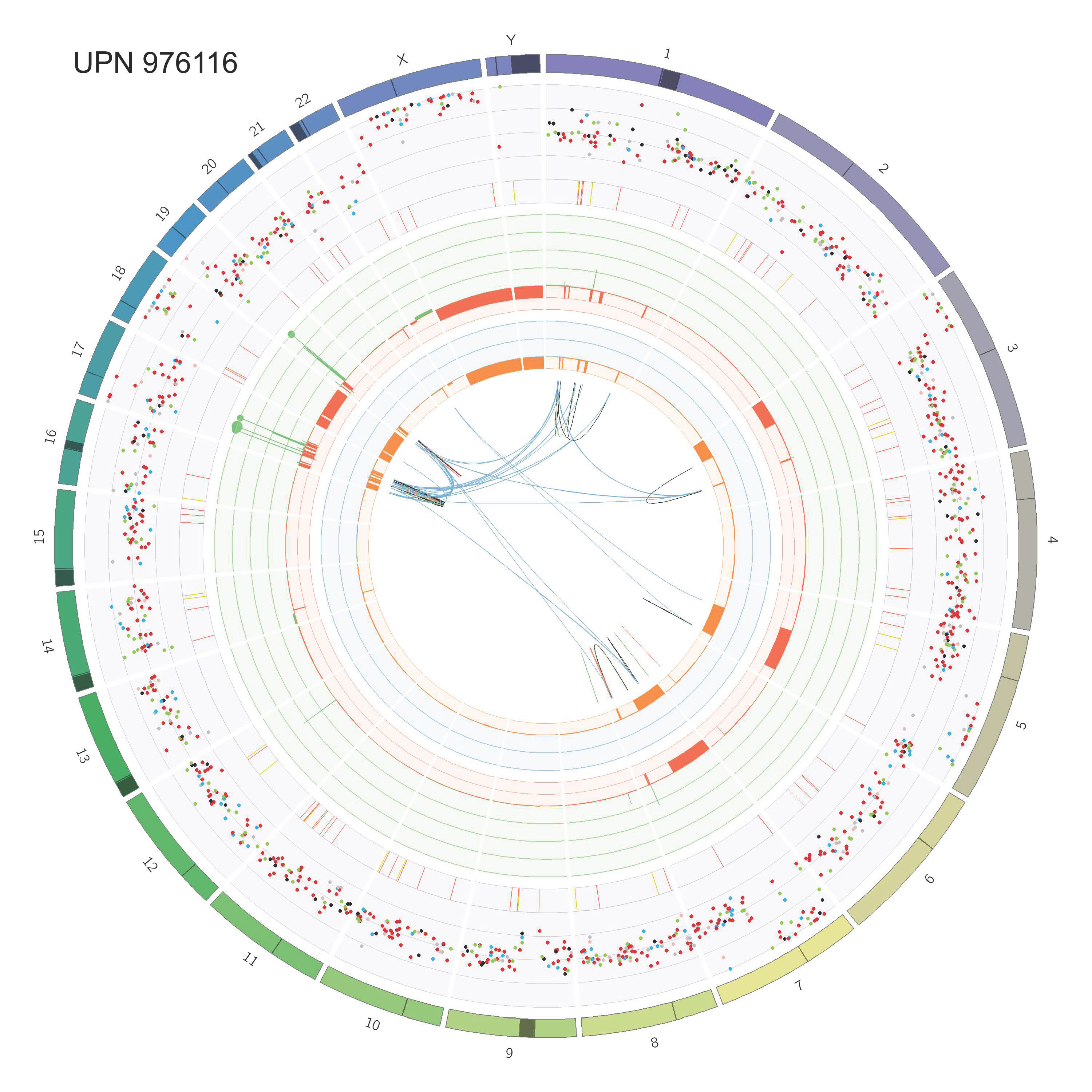

### Supp_Fig_2_circos_Page_58.jpg

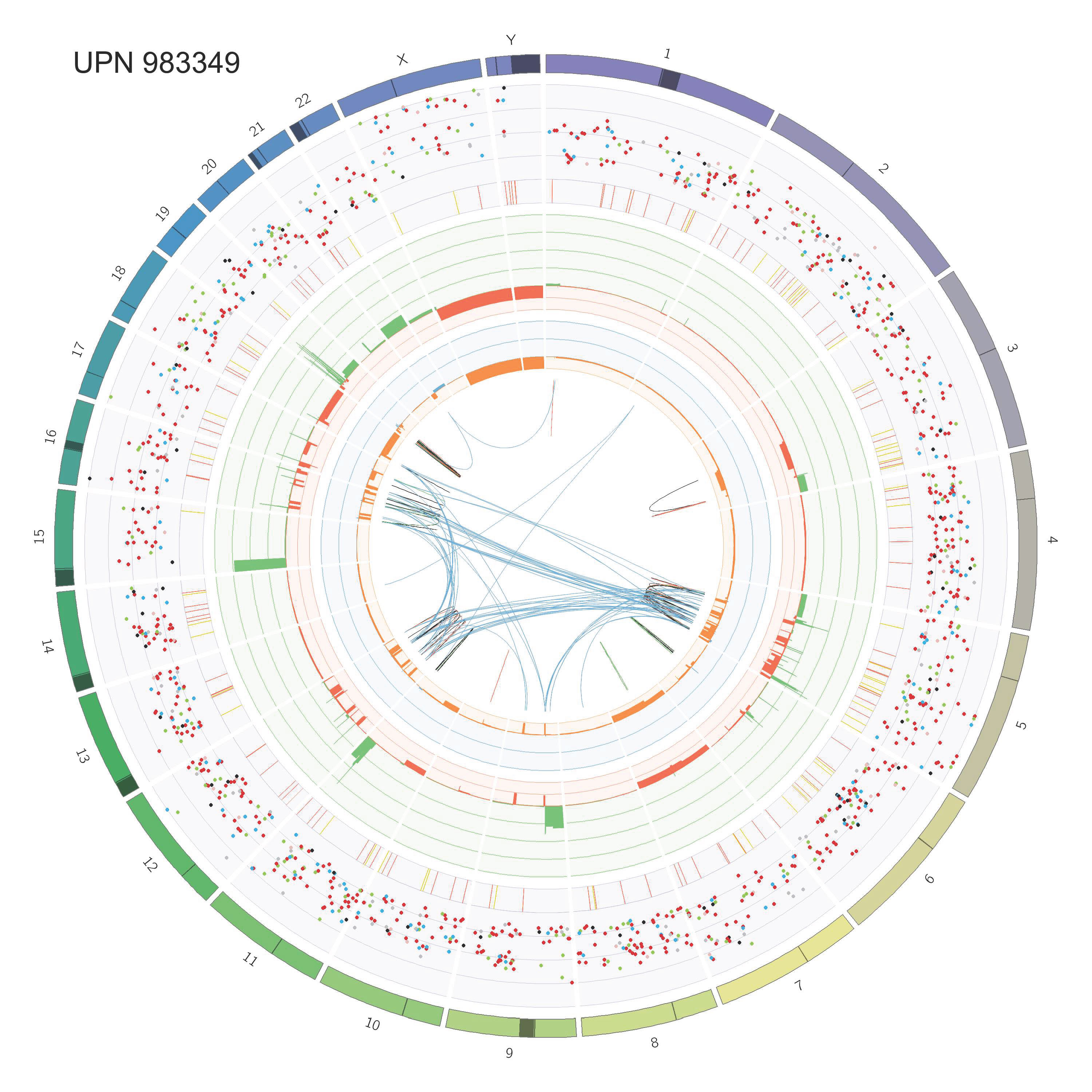

### Supp_Fig_2_circos_Page_59.jpg

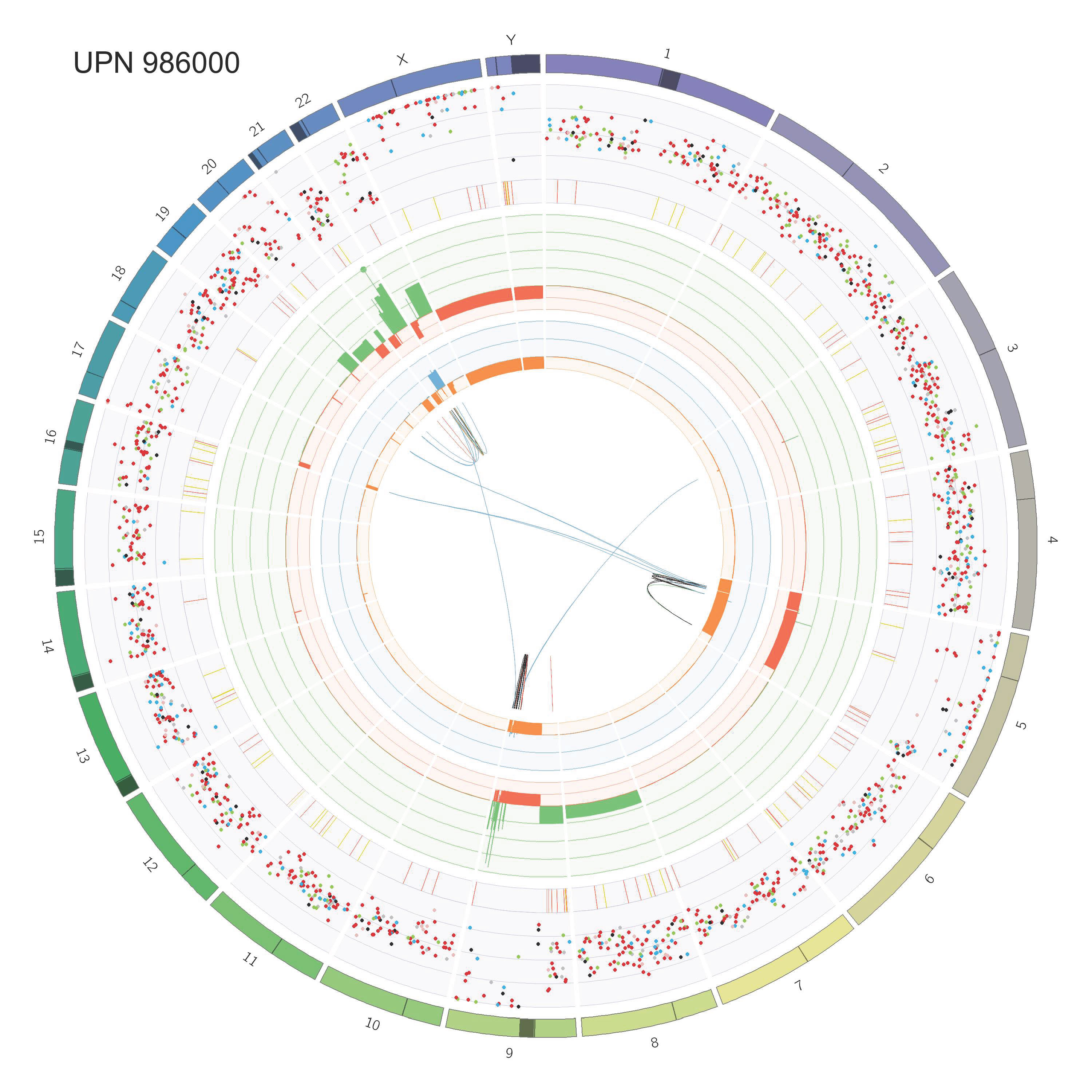

### Supp_Fig_2_circos_Page_60.jpg

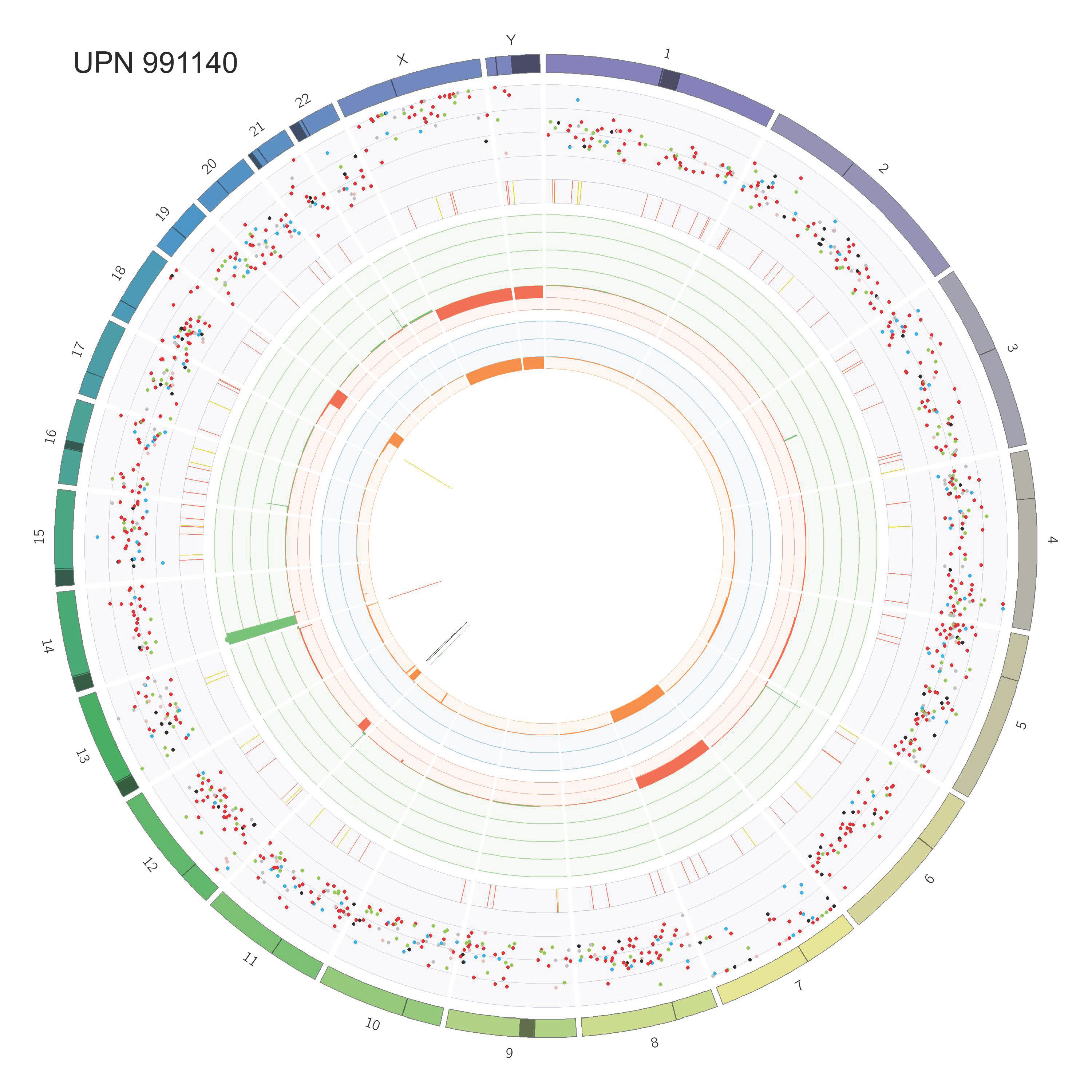

### Supplementary Figure 1

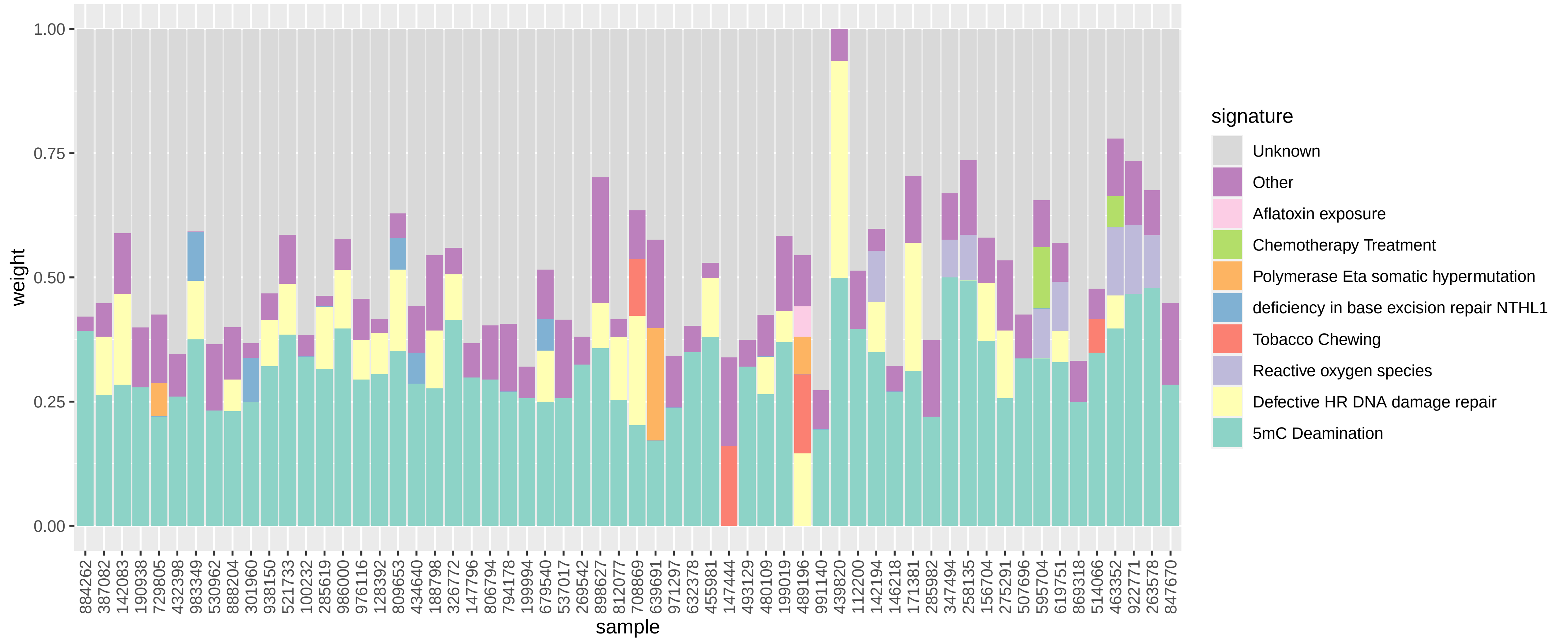

### Supplementary Figure 3

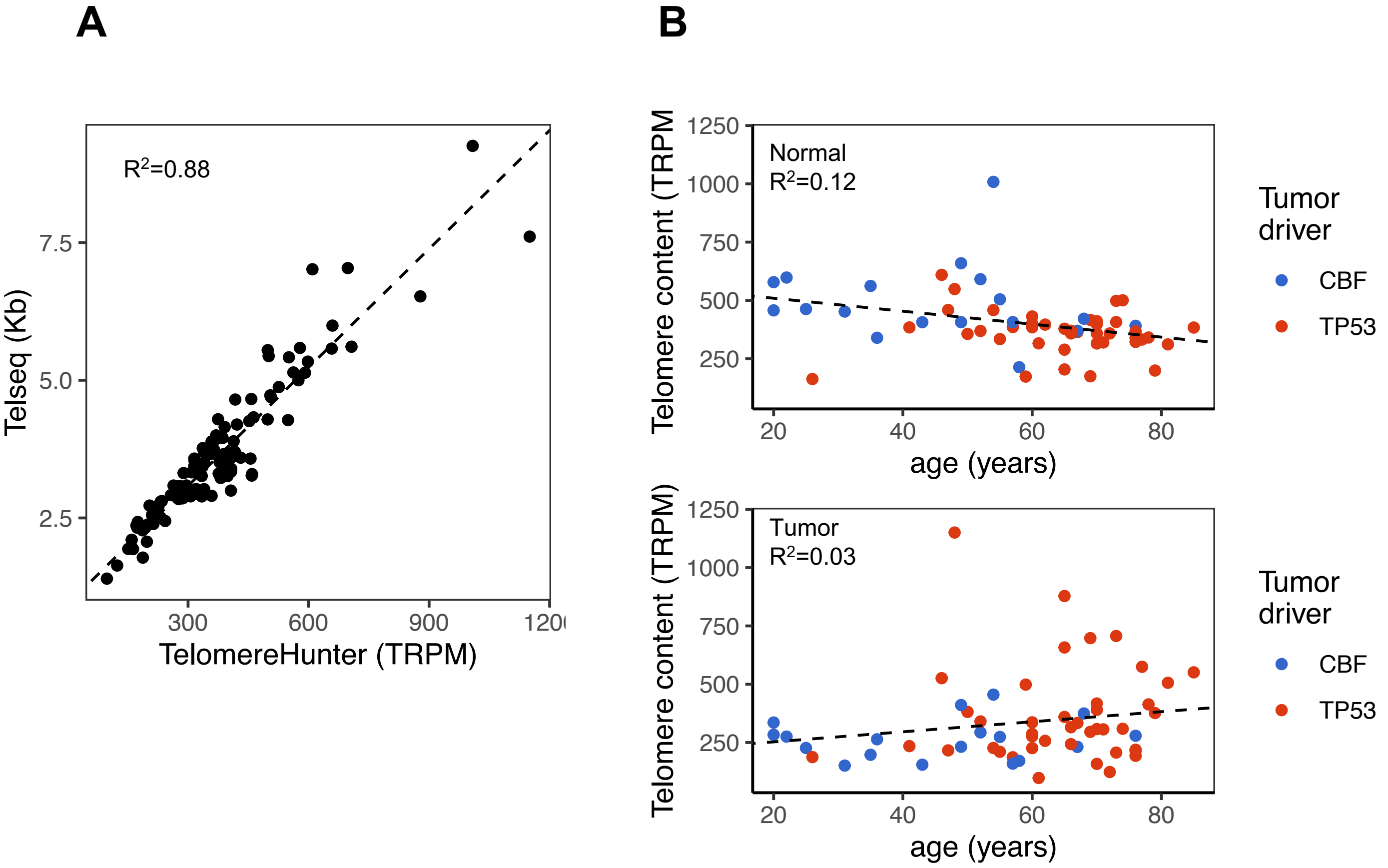

### Supplementary Figure 4

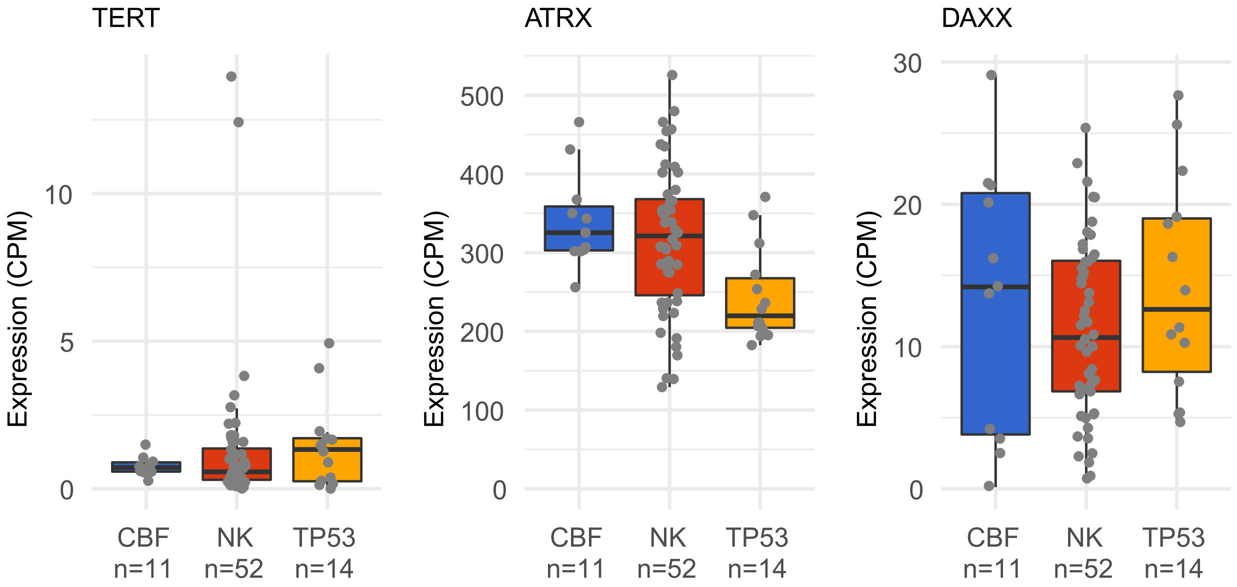
